# Allelic Variation in *HLA-DRB1* is Associated with Development of Anti-Drug Antibodies in Cancer Patients Treated with Atezolizumab that are Neutralizing *in Vitro*

**DOI:** 10.1101/2021.04.29.21256008

**Authors:** Christian Hammer, Jane Ruppel, Julie Hunkapiller, Ira Mellman, Valerie Quarmby

## Abstract

The treatment of diseases with biologic agents can result in the formation of anti-drug antibodies (ADA). The occurrence of ADA were reported for atezolizumab, an anti-PD-L1 monoclonal antibody widely used as immunotherapeutic treatment in cancer patients. Although drivers for ADA formation are unknown, a role for antigen presentation is likely, and variation in human leukocyte antigen (HLA) genes has been shown to be associated with occurrence of ADA for several biologics. Here, we performed an HLA-wide association study in 1,982 patients treated with atezolizumab across 8 clinical trials. On average, 29.8% of patients were ADA positive (N = 591, range of 13.5% -38.4%), and 14.6% of patients were positive for ADA that were neutralizing *in vitro* (NAb, N = 278, range of 6.4% - 21.9%). We found statistically significant associations between HLA class II alleles and ADA status. The top-associated alleles were *HLA-DRB1*01:01* for all ADA (p = 3.4*10^−5^, odds ratio = 1.96), and *HLA-DQA1*01:01* when considering NAb only (p = 2.8 x 10^−7^, OR = 2.31). Both alleles occur together on a common HLA haplotype, and the ADA association was explained by the NAb subset. In conclusion, our study showed that HLA class II genotype contributes to the risk of developing ADA, and specifically NAb, in patients treated with atezolizumab, but suggests that genetic factors are not sufficient as clinically meaningful predictors.

## Introduction

Immune checkpoint blockade (ICB) using therapeutic antibodies has significantly improved clinical outcome and quality of life across a range of cancer indications.^1^ However, the administration of therapeutic proteins (biologics) can lead to unwanted immune responses in the form of anti-drug antibodies (ADA),^2^ which have indeed been observed for all approved immune checkpoint inhibitors (ICI).^3,4^ Reported ADA incidences for atezolizumab, a monoclonal antibody used as immunotherapeutic treatment in cancer patients, are higher than those for other anti PD1/PD-L1 antibodies when they are used as single agent therapy.^3,4^ A subset of ADA-positive patients develop neutralizing antibodies (NAb). By definition, NAb can inhibit the function of a protein therapeutic *in vitro* regardless of their in vivo clinical relevance.^2^ The impact of atezolizumab ADA and NAb on clinical outcomes is currently under evaluation.

The role of patient-related baseline prognostic factors in mediating the risk for ADA formation, and the need to take these into account when assessing potential ADA impact is under active investigation.^5–7^ Given their role in the presentation of peptide antigens to T cells, it seems likely that inherited genetic variation in Human Leukocyte Antigen (HLA) molecules would play a role in ADA development. HLA proteins show a high degree of allelic variation, and the amino acid composition of their antigen-binding groove determines the spectrum of peptides presented.^8^ Indeed, the risk of ADA development during treatment with therapeutic proteins such as interferon beta (IFNβ) or with anti-tumor necrosis factor (anti-TNF) antibodies was previously reported to be associated with specific HLA class II alleles.^9–12^ We therefore hypothesized that the variable presentation of atezolizumab peptides via HLA molecules could contribute to ADA formation. Here, we present what is, to our knowledge, the largest genetic association study for an immunogenicity phenotype, involving a total of 1982 cancer patients treated with atezolizumab. We found statistically significant associations of HLA class II alleles with both ADA and NAb status, and fine-mapped both associations to a single amino-acid residue in the HLA-DRß1 subunit.

## Methods

### Studies and subjects

Patient data from eight atezolizumab phase III clinical trials were included in this analysis (Table 1). Clinical trial results have been previously reported, and the clinical trial protocols have been provided as supplementary materials in the original study publications.^13–21^ Patients included in this study signed an optional Research Biosample Repository (RBR) Informed Consent Form (ICF) and provided whole blood samples. By signing the optional RBR ICF, patients provided informed consent for analysis of inherited and non-inherited genetic variation from whole blood samples. Ethics Committees (EC) and Institutional Review Boards (IRB) in each country and each study site for each clinical trial approved the clinical trial protocol, the main study ICF, and the RBR ICF. The EC and IRB for each clinical trial, country, and study site are provided as Supplementary Data.

**Table 1.** Number of Patients with Available HLA Allele Data, as well as ADA and NAb Frequencies.

| Study name<br>(Study ID) | Indication | Treatment | n ADA<br>Results | n ADA+ (%) | n NAb<br>Results | n NAb+ (%) |
| --- | --- | --- | --- | --- | --- | --- |
| IMvigor211<br>(GO29436) | Urothelial<br>Cancer | Atezolizumab | 216 | 78 (36.1%) | 208 | 40 (19.2%) |
| IMmotion151<br>(WO29637) | Renal Cancer | Atezolizumab + Bev | 243 | 57 (23.5%) | 242 | 47 (19.4%) |
| IMpassion130<br>(WO29522) | TNBC | Atezolizumab + Nab-Paclitaxel | 237 | 32 (13.5%) | 233 | 15 (6.4%) |
| IMpower130<br>(GO29537) | NSCLC | Atezolizumab + Nab-Paclitaxel+CarboP | 215 | 45 (20.9%) | 208 | 16 (7.7%) |
| IMpower131<br>(GO29437) | NSCLC | All Atezolizumab treated | 360 | 132 (36.7%) | 357 | 54 (15.1%) |
|  |  | Atezolizumab + Nab-Paclitaxel + CarboP | 185 | 44 (23.8%) | 184 | 14 (8.2%) |
|  |  | Atezolizumab + Paclitaxel + CarboP | 175 | 88 (50.3%) | 173 | 40 (23.1%) |
| IMpower132<br>(GO29438) | NSCLC | All Atezolizumab treated | 138 | 49 (35.5%) | 137 | 30 (21.9%) |
|  |  | Atezolizumab + Pemetrexed + CisP | 48 | 16 (33.3%) | 47 | 12 (25.5%) |
|  |  | Atezolizumab + Pemetrexed + CarboP | 90 | 33 (36.7%) | 90 | 18 (20.0%) |
| IMpower150<br>(GO29436) | NSCLC | All Atezolizumab treated | 481 | 185 (38.5%) | 436 | 76 (17.4%) |
|  |  | Atezolizumab + Bev + Paclitaxel + CarboP | 229 | 89 (38.9%) | 207 | 36 (17.4%) |
|  |  | Atezolizumab + Paclitaxel + CarboP | 252 | 96 (38.1%) | 229 | 40 (17.5%) |
| IMpower133<br>(GO30081) | SCLC | Atezolizumab + CarboP + Etoposide | 92 | 13 (14.1%) | 91 | 0 (0%) |
| **All** |  | **All Atezolizumab treated** | **1982** | **591 (29.8%)** | **1912** | **278 (14.5%)** |

ADA=anti-drug antibodies; Bev=bevacizumab; CarboP=carboplatin; CisP=cisplatin; NAb=neutralizing antibodies; NSCLC=non-small cell lung cancer; SCLC=small cell lung cancer; TNBC=triple negative breast cancer.

### Detection and analysis of ADA and NAb status

ADA and NAb incidences were systematically assessed in atezolizumab-treated patients in all studies. Analytical methods were developed and run in accordance with industry best practices and health authority guidelines.^2,22–26^ Based on a tiered testing strategy, all ADA samples were tested in an ADA screening assay.^27^ Samples that were deemed ADA positive were subsequently analyzed in a NAb assay. The NAb assay utilized a ligand binding format, and was designed in accordance with industry best practices.^28^ Samples were pretreated prior to NAb analysis to separate ADA from drug in the sample. The resulting ADA solution was then analyzed for its ability to prevent the binding of drug to the PD-L1 target. Based on this, ADA positive samples were deemed NAb positive, NAb negative or NAb indeterminate. As with all NAb assays, ADA/drug stoichiometry does not reflect *in vivo* conditions, and consequently samples that are found to be neutralizing *in vitro* may not impact drug efficacy.^2^

### Whole Genome Sequencing and HLA genotyping

Genomic DNA was extracted from blood samples using the DNA Blood400 kit (Chemagic) and eluted in 50μL Elution Buffer (EB, Qiagen). DNA was sheared (Covaris LE220) and sequencing libraries were prepared using the TruSeq Nano DNA HT kit (Illumina Inc.). Libraries were sequenced at Human Longevity (San Diego, CA, USA). 150bp paired-end whole-genome sequencing (WGS) data was generated to an average read depth of 30× using the HiSeq platform (Illumina X10, San Diego, CA, USA) and processed using the Burrows Wheeler Aligner (BWA)/Genome Analysis Toolkit (GATK) best practices pipeline.^29–31^ Short reads were mapped to hg38/GRCh38 (GCA_000001405.15), including alternate assemblies, using an alt-aware version of BWA to generate BAM files.^32^ All sequencing data was checked for concordance with SNP fingerprint data collected before sequencing.

We used HLA-HD to infer HLA genotypes from whole-genome sequencing data, starting from BAM files generated as described above.^33^ HLA calls at 6-digit resolution were reduced to 4-digit level, reflecting the amino-acid sequence of the HLA protein.

### Statistical analyses

The association analysis focused on ADA or NAb presence versus absence phenotypes, not considering the timing of ADA or NAb occurrence. Logistic regression was used on a per-trial basis to test for association between ADA or NAb, respectively, and HLA alleles with carrier frequencies of at least 0.5% (dominant inheritance model). We included age, sex, five principal components (PC) to correct for population stratification, and trial arm (where applicable) as covariates. We used a multi-degree-of-freedom omnibus test to test for association at multi-allelic amino acid positions in the coding region of each gene. For the meta-analyses, the random-effects model method in the R package “meta” was used to calculate effect estimates, 95% confidence intervals, and p-values.^34^ Edginton’s method (sum of p), as implemented in the R package “metap”, was used for meta-analysis of omnibus test results for variable amino acid positions. The Bonferroni method was applied to correct p-values for multiple testing, correcting for the number of included HLA alleles or variable amino acid positions, respectively.

## Results

In total, our study included 1982 patients treated with atezolizumab as mono- or combination therapy and with available informed consent for genetic analyses. Out of these patients, 591 (29.8%) tested positive for ADA, and we observed a large variability in ADA incidences between treatment combinations and study arms (13.5% - 50.3%, Table 1). Samples from the ADA-positive patients were further tested for the presence of neutralizing antibodies (NAb), and we found that only a subset of these (N=278; 14.5%) were also NAb-positive (0% - 25.5%, Table 1).

In a meta-analysis across the eight studies, we identified a statistically significant association of five HLA class II alleles with the presence of ADA (Suppl. Results), with *HLA-DRB1*01:01* showing the strongest association (p = 3.4*10^−5^, OR = 1.96; Figure 1, Figure 2A). The top three alleles, *HLA-DRB1*01:01, HLA-DQB1*05:01*, and *HLA-DQA1*01:01* form a common HLA class II haplotype and are unlikely to be statistically independent. Indeed, *HLA-DQB1*05:01* and *HLA-DQA1*01:01* were no longer significantly associated in a conditional analysis, including *HLA-DRB1*01:01* as a covariate (Suppl. Results). We also inferred variable amino acid positions across all tested HLA genes. Position 96 of HLA-DRß1 showed the strongest significance (p = 4.8*10^−5^), and glutamic acid as residue at this position was associated with increased risk for ADA (OR = 1.91; Figure 1, Figure 2A). Of all alleles tested, only *HLA-DRB1*01* alleles *HLA-DRB1*01:01* and *HLA-DRB1*01:02*) carried this residue at position 96 (Table 2), and the amino acid residue did not explain the association better than the top-associated allele (Figure 1). Conditional analysis, including the top-associated position as covariate, did not yield further amino acid positions that were independently associated.

**Table 2.** Meta-analysis association results for amino acid residues at HLA-DRß1 position 96.

| HLA-DRβ1<br>(96) | ADA | | NAb | | *HLA-DRB1* alleles (1-field)<br>corresponding to amino-acid residue |
| --- | --- | --- | --- | --- | --- |
|  | P | OR (95% CI) | P | OR (95% CI) |  |
| Glutamic acid | 1.89E-04 | 1.91 (1.57-2.25) | 1.94E-07 | 2.33 (2.01-2.65) | *01 |
| Histidine | 0.56 | 1.11 (0.76-1.45) | 0.62 | 0.87 (0.33-1.42) | *03,*07,*08,*09,*11,*12,*13,*14 |
| Glutamine | 0.03 | 0.68 (0.33-1.02) | 0.10 | 0.66 (0.18-1.15) | *10,*15,*16 |
| Tyrosine | 0.56 | 1.07 (0.84-1.31) | 0.78 | 0.95 (0.55-1.34) | *04 |

ADA=anti-drug antibodies; NAb=neutralizing antibodies; OR=odds ratio; CI = confidence interval.

**Figure 1.**
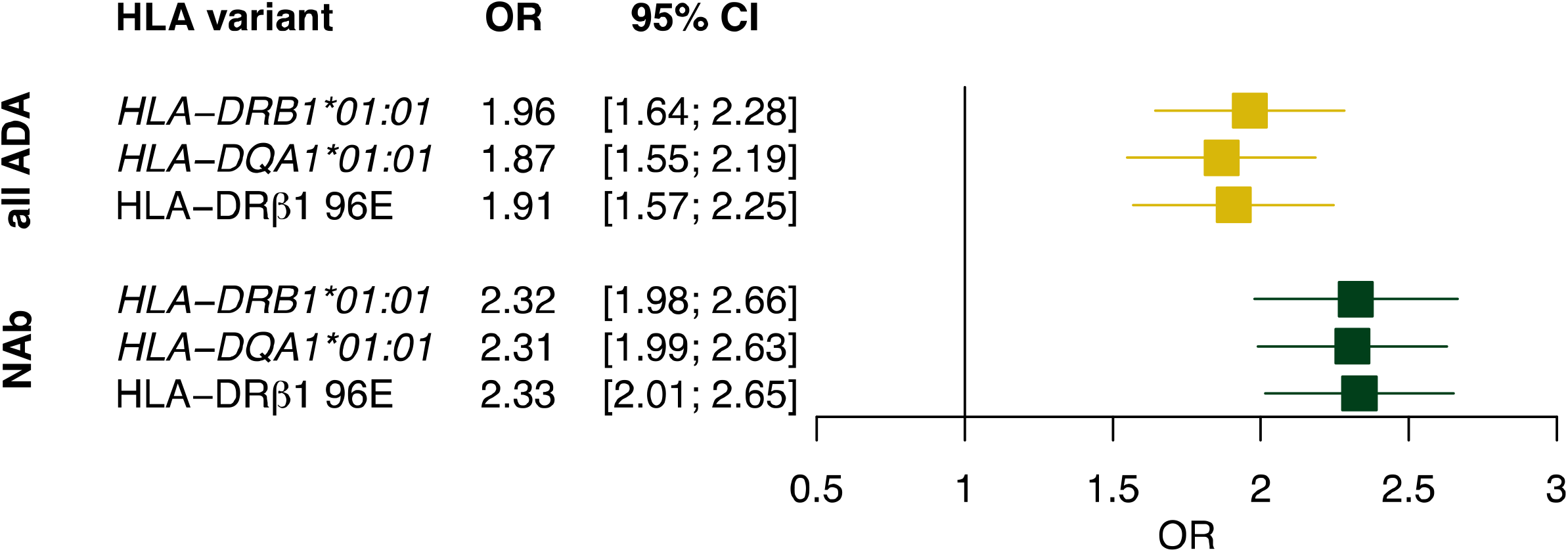
Meta-analysis summary results for the top-associated HLA alleles and amino acid residue. Forest plot showing the meta-analysis association effect estimates of the top-associated HLA alleles *HLA-DRB1*01:01* and *HLA-DQA1*01:01*, as well as amino acid residue 96E (Glutamic acid) of HLA-DRß1, with ADA and NAb status, respectively. Bars represent 95% confidence intervals. ADA = anti-drug antibodies; NAb = *in vitro* neutralizing antibodies; OR = odds ratio; CI = confidence interval.

**Figure 2.**
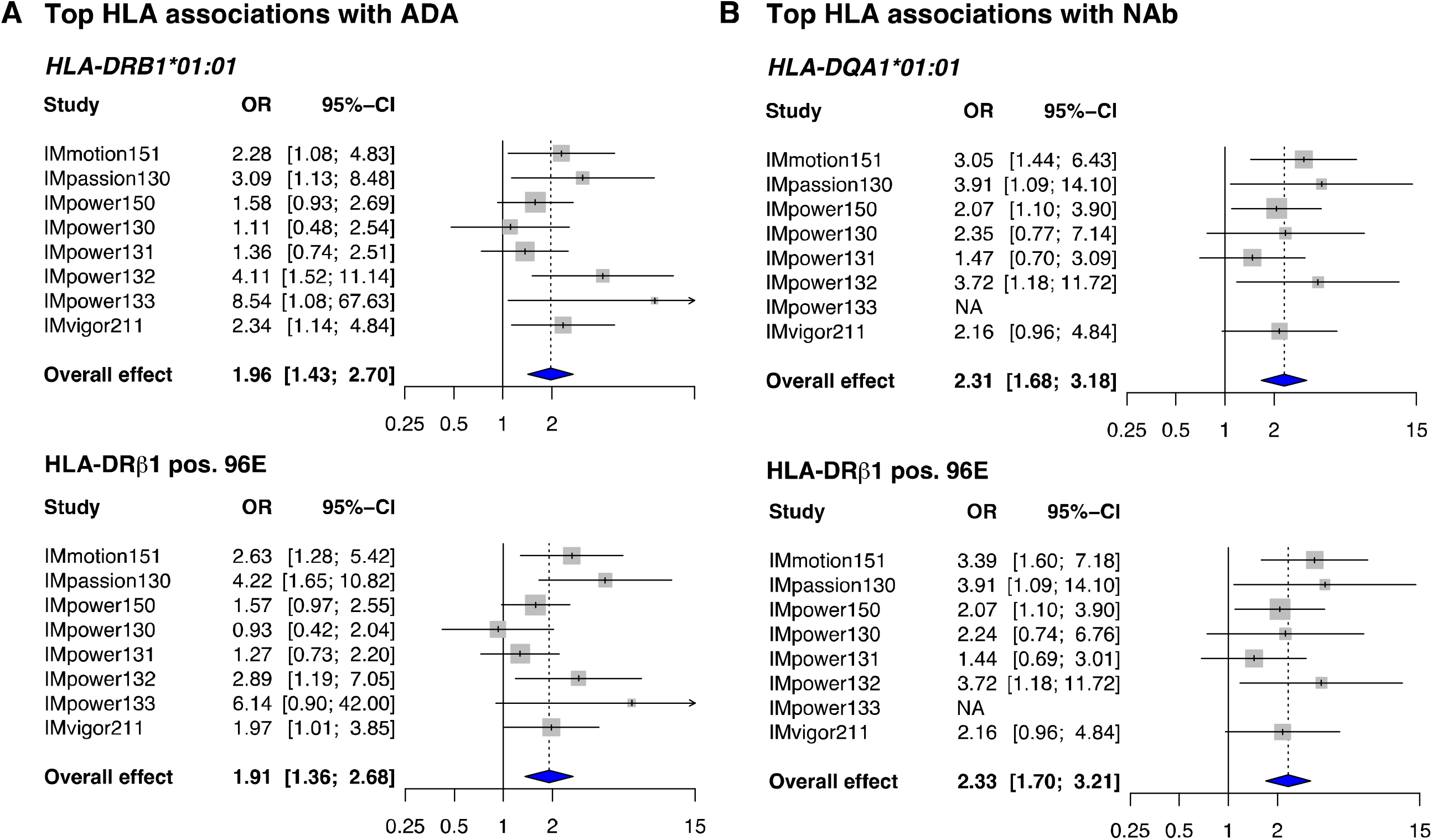
Per-study association results for the top-associated HLA alleles and amino acid residue. A) Forest plots showing the single study and meta-analysis effect estimates of the top-associated HLA allele *HLA-DRB1*01:01* and amino acid residue 96E (Glutamic acid) of HLA-DRß1 with ADA status. B) Forest plots showing the single study and meta-analysis effect estimates of the top-associated HLA allele *HLA-DQA1*01:01* and amino acid residue 96E (Glutamic acid) of HLA-DRß1 with NAb status. Squares represent relative study size; bars represent 95% confidence intervals, X-axes depict OR. ADA = anti-drug antibodies; NAb = *in vitro* neutralizing antibodies; OR = odds ratio; CI = confidence interval.

Next, we focused on patients with NAb (N = 278), excluding individuals with ADA that were not neutralizing from the analysis. The top-associated HLA allele with NAb status was *HLA-DQA1*01:01* (p = 2.8 x 10^−7^, OR = 2.31; Figure 1, Figure 2B), a member of the same common haplotype that includes *HLA-DRB1*01:01*. In fact, *HLA-DRB1*01:01* was the second strongest associated allele (p = 1.6 x 10^−6^, OR = 2.32) in the NAb analysis (Figure 1, Figure 2B). On amino acid level, position 96 of HLA-DRß1 again showed the strongest association (p = 3.2 x 10^−5^), with glutamic acid mediating increased risk for NAb development (OR = 2.33, Figure 1, Figure 2B). Conditional analysis revealed no statistically independent associations at the HLA allele or amino acid level.

Both ADA and NAb analyses yielded associations of the same group of alleles and amino acid positions, suggesting that the ADA associations are explained by the subset of patients exhibiting NAb. We therefore also investigated possible HLA associations with non-neutralizing ADA only (N = 313), and did not obtain significant results on allele or amino acid level after multiple testing correction. *HLA-DRB1*01:01* and *HLA-DQA1*01:01* yielded p values of 0.047 and 0.18, respectively, suggesting that ADA associations are predominantly driven by NAb status (Suppl. Results).

## Discussion

Although the formation of ADA is associated with many or most therapeutic antibodies, little is known about the risk factors that predispose individual patients or therapeutic antibodies to their elicitation. Moreover, ADA can exhibit variable effects, with some manifesting as antibodies that do not interfere with therapeutic efficacy, while others are neutralizing *in vivo* and therefore have greater potential to attenuate efficacy. Given that we were able to analyze a large patient cohort, our study presents clear evidence for an association of common HLA class II variation with anti-atezolizumab ADA risk. *HLA-DRB1*01:01* and *HLA-DQA1*01:01* were the top-associated alleles for ADA and NAb status, respectively. Both occur together on a common HLA class II haplotype, and each showed statistically significant association with both phenotypes. This is consistent with a single amino acid position and residue (glutamic acid at HLA-DRß1 position 96) being most strongly associated with both ADA and NAb status. Also, no significant associations were obtained when considering only patients who exhibited non-neutralizing antibodies. Thus, most of the association with ADA can be explained by the NAb subset. 47.2% of ADA-positive patients (278 of 591) carried detectable antibodies that are neutralizing *in vitro*, suggesting a certain degree of variation in antibody clonality. The possibility that HLA class II proteins including the associated alpha or beta subunits result in the presentation of a peptide that specifically contributes to NAb formation is consistent with the fact that different atezolizumab peptides are presented by different allelic variants of HLA proteins.

Across the eight studies, 81% of patients identified as White / European (Suppl. Table 1). Both identified risk alleles show the highest carrier frequencies in White / European reference populations. *HLA-DRB1*01:01* is significantly less common for example in African American and Chinese individuals.^35^ Due to the low number of non-European patients, our study was probably underpowered to establish possible additional associations of ADA status with HLA alleles that are frequent in such populations, but rare in Europeans. Future studies in more diverse populations will be required to answer this question. To exclude the possibility that our significant results were due to population stratification, we included five genetic principal components as covariates in all statistical analyses.

Of note, effect directionality was consistent across the investigated trials, providing evidence that the associations are specific to the treatment, but independent of cancer indication. In terms of effect estimates, the odds ratios for associated alleles in the range of 2 are comparable to reported HLA associations with ADA development for other biologics.^9–12^ This implies that carrying a risk genotype is neither necessary nor sufficient to predict development of ADA. It is more likely one out of many factors that together determine ADA risk, and many of these factors remain to be uncovered.^36,37^ However, the identification of HLA risk genotypes combined with *in silico* peptide binding prediction might support efforts to identify immunogenic peptides in atezolizumab or possibly in other antibodies or protein biologics. In conclusion, our results provide strong evidence that HLA allelic variation can contribute to the development of ADAs in patients treated with therapeutic antibodies. It will be important to assess whether similar associations exist in patient groups treated with other therapeutic antibodies that are known to elicit ADA.

## Supporting information

Supplementary Results

Ethics Committee approvals

## Data Availability

HLA association summary statistics are available as Supplementary Results. Qualified researchers may request access to individual patient data used in this study through Roche's data sharing platforms in accordance with the Global Policy on Sharing of Clinical Study Information:
http://www.roche.com/research_and_development/who_we_are_how_we_work/clinical_trials/our_commitment_to_data_sharing.htm
Whole genome sequencing data collected is in this study available under restricted access for biomedical research use only. Applications for access to individual level data should be made to the corresponding authors by email. Access to individual level data will require applicants to establish a data sharing agreement with Roche/Genentech to ensure compliance with patient consent and confidentiality obligations.

## Data availability

HLA association summary statistics are available as Supplementary Results. Qualified researchers may request access to individual patient data used in this study through Roche’s data sharing platforms in accordance with the Global Policy on Sharing of Clinical Study Information: http://www.roche.com/research_and_development/who_we_are_how_we_work/clinical_trials/our_commitment_to_data_sharing.htm

Whole genome sequencing data collected is in this study available under restricted access for biomedical research use only. Applications for access to individual level data should be made to the corresponding author by email. Access to individual level data will require applicants to establish a data sharing agreement with Roche/Genentech to ensure compliance with patient consent and confidentiality obligations.

**Suppl. Table 1.** Number of patients by race.

|  | American<br>Indian or<br>Alaska Native | Asian | Black or<br>African<br>American | Multiple | Unknown | White | TOTAL |
| --- | --- | --- | --- | --- | --- | --- | --- |
| IMmotion151 | 2 | 22 | 1 | 0 | 30 | 188 | 243 |
| IMpassion130 | 11 | 44 | 12 | 0 | 11 | 159 | 237 |
| IMpower150 | 0 | 49 | 9 | 1 | 13 | 409 | 481 |
| IMpower130 | 0 | 5 | 9 | 1 | 12 | 188 | 215 |
| IMpower131 | 1 | 39 | 3 | 3 | 6 | 308 | 360 |
| IMpower132 | 1 | 17 | 0 | 0 | 10 | 110 | 138 |
| IMpower133 | 0 | 15 | 0 | 0 | 1 | 76 | 92 |
| IMvigor211 | 0 | 9 | 0 | 0 | 40 | 167 | 216 |
| TOTAL | 15 | 200 | 34 | 5 | 123 | 1605 | 1982 |

