## Supplementary material for "Allelic Variation in *HLA-DRB1* is Associated with Development of Anti-Drug Antibodies in Cancer Patients Treated with Atezolizumab that are Neutralizing *in Vitro*": Ethics Committee approvals: IMmotion151 EC IRB.pdf

### LIST OF IRB/IEC - PROTOCOL WO29637

| Country | Site No. | Principal Investigator | EC/IRB | Approved Date |
|---|---|---|---|---|
| Australia | 277614 | Gurney H | Macquarie University Human Research Ethics Committee | 18 Jun 2015 |
| Australia | 277615 | Feeney K | St John of God Health Care Ethics Committee | 28 May 2015 |
| Australia | 277616 | Weickhardt A | Austin Health HREC, Research Ethics Unit | 10 Aug 2015 |
| Australia | 277617 | Parnis F | Bellberry Human Research Ethics Committee | 03 Jun 2015 |
| Australia | 277813 | Harrison M | Concord Repatriation General Hospital HREC | 09 Jul 2015 |
| Australia | 279380 | Gedye C | Austin Health; Austin Health Human Research Ethics Committee | 25 Jun 2015 |
| Australia | 280297 | Goh J | Bellberry Human Research Ethics Committee | 04 Jun 2015 |
| Bosnia and Herzegovina | 289848 | Jungic S | Ethics Committee University Clinical Centre of the Republic of Srpska | 10 Feb 2016 |
| Brazil | 290237 | Barrios.C | CEP PUCRS | 22 Dec 2015 |
| Brazil | 290243 | Pereira R | CEP ISCMPOA | 03 Aug 2016 |
| Brazil | 290244 | Franke F | Comitê de Ética em Pesquisa; Universidade Regional do Noroeste do Estado do Rio Grande do Sul UNI JUÍ | 12 Aug 2016 |
| Brazil | 290673 | Bastos D | CEP para Analise de Projetos de Pesquisa do HCFMUSP e da FMUSP;Hospital da Universidade de São Paulo | 25 Jul 2016 |
| Canada | 276934 | Hotte S | Ontario Cancer Research Ethics Board | 18 Dec 2016 |
| Canada | 276940 | Russell K | Ontario Cancer Research Ethics Board | 18 Nov 2016 |
| Canada | 276941 | Hansen A | Ontario Cancer Research Ethics Board | 6 Jan 2016 |

| Country | Site No. | Principal Investigator | EC/IRB | Approved Date |
|---|---|---|---|---|
| Canada | 276942 | Bjarnason G | Ontario Cancer Research Ethics Board | 1 Feb 2016 |
| Canada | 276944 | Zalewski P | Lakeridge Health Research Ethics Board | 20 Jul 2015 |
| Canada | 276945 | Reaume M | Ontario Cancer Research Ethics Board | 14 Jul 2015 |
| Canada | 276946 | Miller W | McGill University; Sir Mortimer B Davis Jewish General Hospital; Ethics Board | 17 Jul 2015 |
| Canada | 276949 | Wood L | Nova Scotia Health Authority Research Ethics Board | 24 Apr 2015 |
| Canada | 285313 | Lacombe L | McGill University; Sir Mortimer B Davis Jewish General Hospital; Ethics Board | 06 Nov 2015 |
| Canada | 288927 | Potvin K | Ontario Cancer Research Ethics Board | 18 Jan 2016 |
| Czech Republic | 279296 | Melichar B | Etická komise Fakultní nemocnice Olomouc | 14 Sep 2015 |
| Czech Republic | 279456 | Poprach A | Etická komise Masarykův onkologický ústav v Brně | 21 Jul 2015 |
| Czech Republic | 279457 | Buchler T | Etická komise při IKEM a TN, Thomayerova nemocnice | 08 Jul 2015 |
| Czech Republic | 292721 | Mateju M | Etická komise Všeobecné fakultní nemocnice v Praze | 19 May 2016 |
| Denmark | 278967 | Jensen N | Videnskabsetiske Komité Region Midt; Sundhedssekr. | 09 Jul 2015 |
| Denmark | 278973 | Geertsen P | Videnskabsetiske Komité Region Midt; Sundhedssekr. | 09 Jul 2015 |
| Denmark | 279352 | Donskov F | Videnskabsetiske Komité Region Midt; Sundhedssekr. | 09 Jul 2015 |
| France | 278954 | Escudier B | CPP Ile de France VII | 11 Aug 2015 |

| Country | Site No. | Principal Investigator | EC/IRB | Approved Date |
|---|---|---|---|---|
| France | 278957 | Oudard S | CPP Ile de France VII | 11 Aug 2015 |
| France | 278958 | Abadie Lacourtoisie S | CPP Ile de France VII | 11 Aug 2015 |
| France | 278961 | Culine S | CPP Ile de France VII | 11 Aug 2015 |
| France | 278962 | Amela Y | CPP Ile de France VII | 11 Aug 2015 |
| France | 278963 | Negrier S | CPP Ile de France VII | 11 Aug 2015 |
| France | 280066 | Spaeth D | CPP Ile de France VII | 11 Aug 2015 |
| France | 282441 | Joly Lobbedez F | CPP Ile de France VII | 11 Aug 2015 |
| France | 288395 | Gross Goupil M | CPP Ile de France VII | 11 Aug 2015 |
| France | 290462 | Rolland F | CPP Ile de France VII | 11 Aug 2015 |
| France | 293019 | Gravis Mescam G | CPP Ile de France VII | 11 Aug 2015 |
| Germany | 279040 | Grüllich C | Ethik-Kommission der Medizinischen Fakultät Heidelberg | 15 Aug 2015 |
| Germany | 279041 | Virchow I | Geschäftsstelle der Ethik-Kommission der Medizinischen Fakultät der Universität Duisburg-Essen | 15 Aug 2015 |
| Germany | 279043 | Bedke J | Ethik-Kommission an der Medizinischen Fakultät der Eberhard-Karls-Universität und am Universitätsklinikum Tübingen | 15 Aug 2015 |
| Germany | 279045 | Staehler M | Ethik-Kommission der medizinischen Fakultät der Ludwig-Maximilians-Universität | 15 Aug 2015 |
| Italy | 278937 | Bracarda S | Comitato Etico Area Vasta Sud Est | 18 May 2015 |
| Italy | 279094 | De Giorgi U | Comitato Etico Di Area Vasta Romagna E Irst | 11 Jun 2015 |

| Country | Site No. | Principal Investigator | EC/IRB | Approved Date |
|---|---|---|---|---|
| Italy | 279102 | Sabbatini R | Comitato Etico Provinciale Modena | 07 Jul 2015 |
| Italy | 279103 | Carteni G | Comitato Etico dell' Azienda osped. A. Cardarelli | 28 May 2015 |
| Italy | 279105 | Sternberg C | Comitato Etico A.O. S. Camillo Forlanini | 09 Sep 2015 |
| Italy | 279109 | Gianni L | Comitato Etico Irccs Ospedale San Raffaele | 04 Jun 2015 |
| Italy | 279110 | Porta C | Comitato Etico Dell'irccs San Matteo Di Pavia | 26 Oct 2015 |
| Italy | 279135 | Siena.S | Comitato Etico Milano Area C C/O A.O.Ospedale Niguarda Ca' Granda | 18 Sep 2015 |
| Japan | 287009 | Sassa N | Nagoya university Hospital IRB | 14 Sep 2015 |
| Japan | 287044 | Fukasawa S | Chiba Cancer Center Institutional Review Board | 12 Aug 2015 |
| Japan | 287087 | Nishimura K | Institutional Review Board of Osaka International Cancer Institute | 20 Aug 2015 |
| Japan | 287191 | Obara W | Iwate Medical University Institutional Review Board | 21 Aug 2015 |
| Japan | 287192 | Uemura M | Osaka University Hospital Institutional Review Board | 29 Sep 2015 |
| Japan | 287193 | Kanayama H-O | Tokushima University Hospital Institutional Review Board | 20 Aug 2015 |
| Japan | 287194 | Yatsuda J | The Institutional Review Board of Kumamoto University Hospital | 15 Sep 2015 |
| Japan | 287301 | Yonese J | The Cancer Institute Hospital of JFCR Institutional Review Board | 19 Oct 2015 |
| Japan | 287420 | Tomita Y | Niigata University Medical & Dental Hospital IRB | 26 Aug 2015 |
| Japan | 287421 | Osawa T | Hokkaido University Hospital Institutional Review Board | 25 Aug 2015 |
| Japan | 287422 | Takagi T | Tokyo Women's Medical University Hospital IRB | 14 Aug 2015 |
| Japan | 287423 | Kimura G | Nippon Medical School Hospital IRB | 10 Aug 2015 |

| Country | Site No. | Principal Investigator | EC/IRB | Approved Date |
|---|---|---|---|---|
| Japan | 287424 | Nozawa O | Kindai University Hospital Institutional Review Board | 15 Sep 2015 |
| Japan | 287444 | Kojima T | University of Tsukuba Hospital IRB | 28 Sep 2015 |
| Japan | 287476 | Oya M | Keio University Hospital IRB | 01 Oct 2015 |
| Japan | 287478 | Kondo K | Yokohama City University Hospital IRB | 30 Sep 2015 |
| Japan | 291228 | Takano T | Toranomon Hospital and Toranomon Hospital Kajigaya Institutional Review Board | 03 Mar 2016 |
| Japan | 292487 | Tamada S | Osaka City University Hospital IRB | 24 Mar 2016 |
| Japan | 292697 | Takamoto A | Okayama University Hospital Institutional Review Board | 19 Apr 2016 |
| Japan | 292698 | Tatsugami K | Kyushu University Hospital IRB | 01 Jun 2016 |
| Japan | 292699 | Iwamura M | Kitasato University Sagamihara Institutional Review Board | 21 Apr 2016 |
| Japan | 292700 | Fujii Y | Medical Hospital, Tokyo Medical and Dental University Institutional Review Board | 31 May 2016 |
| Japan | 292765 | Suzuki K | Gunma University Hospital Institutional Review Board | 28 Apr 2016 |
| Korea, Republic Of | 278877 | Keam B | Seoul National University Hospital; IRB | 04 Oct 2016 |
| Korea, Republic Of | 278879 | Rha S | SeveranceHospital YonseiUniversity; IRB | 24 Apr 2015 |
| Korea, Republic Of | 278880 | Park S | Samsung Medical Center, IRB | 28 May 2015 |
| Korea, Republic Of | 278885 | Lee J | Asan Medical Center Ethics Committee; Asan Medical Center; IRB | 24 Jun 2015 |

| Country | Site No. | Principal Investigator | EC/IRB | Approved Date |
|---|---|---|---|---|
| Korea, Republic Of | 279439 | Chung J | National Cancer Center Institutional Review Board | 01 Jun 2015 |
| Korea, Republic Of | 280149 | Kim S | Seoul National University Bundang Hospital IRB | 18 May 2015 |
| Korea, Republic Of | 280202 | Lee H | Chungnam National University Hospital; IRB | 20 Jul 2015 |
| Mexico | 281262 | Dominguez A | CEI de Clinica Bajio CLINBA; Comite de Etica en Investigacion | 11 May 2016 |
| Mexico | 282026 | Campos S | CEI de Clinica Bajio CLINBA; Comite de Etica en Investigacion | 18 May 2016 |
| Poland | 279461 | Tomczak P | Komisja Bioetyczna przy Uniwersytecie Medycznym im. K. Marcinkowskiego. | 01 Oct 2015 |
| Poland | 279463 | Zolnierek J | Komisja Bioetyczna przy Uniwersytecie Medycznym im. K. Marcinkowskiego. | 01 Oct 2015 |
| Poland | 279600 | Kukielka-Budny B | Komisja Bioetyczna przy Uniwersytecie Medycznym im. K. Marcinkowskiego. | 01 Oct 2015 |
| Poland | 292740 | Piętak K | Komisja Bioetyczna przy Uniwersytecie Medycznym im. K. Marcinkowskiego. | 16 Jun 2016 |
| Russian Federation | 279491 | Stroyakovskii D | EC of Moscow City Oncol. Hospital #62 | 14 Oct 2016 |
| Russian Federation | 279619 | Alexeev B | EC at FSI MSROI n.a. Hertsen of Rosmedtechnology | 28 Oct 2016 |
| Russian Federation | 279623 | Alyasova A | EC of FSBI Privolzhsky Federal Medical Research Centre | 16 Jul 2016 |
| Russian Federation | 279624 | Varlamov S | E.C. of Altai Oncological Center | 01 Jul 2016 |
| Singapore | 280295 | Kanesvaran R | SingHealth Centralised IRB; Review Board B | 16 Feb 2016 |

| Country | Site No. | Principal Investigator | EC/IRB | Approved Date |
|---|---|---|---|---|
| Singapore | 280296 | Wong A | SingHealth Centralised IRB; Review Board B | 01 Jun 2017 |
| Spain | 278857 | Suarez Rodriguez C | CEIC Hospital Vall D'Hebron | 07-Apr-2015 |
| Spain | 278859 | Castellano D | CEIC Hospital Universitario 12 de Octubre | 07-Apr-2015 |
| Spain | 278860 | Arranz Arija J | CEIC del Hospital Gregorio Marañón | 07-Apr-2015 |
| Spain | 278861 | Alonso Gordoa T | Hospital Ramon y Cajal ;Comité Etico de Investigación Clínica | 07-Apr-2015 |
| Spain | 278862 | Duran Martinez I | Hospital Virgen del Rocio, CEIC | 07-Apr-2015 |
| Spain | 278864 | Garcia Del Muro K | CEIC Hospital de Bellvitge | 07-Apr-2015 |
| Spain | 278865 | Gallardo Diaz E | CEIC Corporacio Sanitaria Parc Tauli | 07-Apr-2015 |
| Spain | 278867 | Mendez M | Comité Coordinador de Ética de la Investigación Biomédica de Andalucía (CEIBA) | 07-Apr-2015 |
| Spain | 279661 | Mellado Gonzalez B | Hospital Clinic I Provincial; Comité Etico de Investigacion Clinica | 07-Apr-2015 |
| Taiwan | 277866 | Chuang C | Chang Gung Med Found, Institutional Review Board | 21 May 2015 |
| Taiwan | 277867 | Lin C | Research Ethics Committee, Nat. Taiwan Univ. Hosp. | 27 May 2015 |
| Taiwan | 277868 | Yang C | The IRB, Taichung Veterans General Hospital | 27 Apr 2015 |
| Thailand | 288923 | Srimuninnimit V | Siriraj Institutional Review Board | 07 Jan 2016 |
| Thailand | 288924 | Chansriwong P | Ethical Clearance Committee on Human Rights | 15 Dec 2015 |
| Thailand | 288925 | Dechaphunkul A | Songklanagarind Ethics Committee | 10 Feb 2016 |
| Thailand | 289042 | Sriuranpong V | Institutional Review Board, Faculty of Medicine | 05 Jan 2016 |

| Country | Site No. | Principal Investigator | EC/IRB | Approved Date |
|---|---|---|---|---|
| Thailand | 289044 | Sriparakij S | Research Ethics Com. Fac Med. Chiang May University | 27 Jan 2016 |
| Turkey | 279005 | Ozguroglu M | Trakya University Medical School Ethics Commitee (for initial approval); İstanbul Uni. Cerrahpasa Medical School Ethics Commitee (current EC due legislation change) | 20 May 2015 |
| Turkey | 279023 | Cicin I | Trakya University Medical School Ethics Commitee (for initial approval); İstanbul Uni. Cerrahpasa Medical School Ethics Commitee (current EC due legislation change) | 20 May 2015 |
| Turkey | 279072 | Erman M | Trakya University Medical School Ethics Commitee (for initial approval); İstanbul Uni. Cerrahpasa Medical School Ethics Commitee (current EC due legislation change) | 20 May 2015 |
| United Kingdom | 277295 | Hawkins R | London Central Research Ethics Committee | 09 Sep 2016 |
| United Kingdom | 277296 | Griffiths R | London Central Research Ethics Committee | 09 Oct 2015 |
| United Kingdom | 277298 | Powles T | London Central Research Ethics Committee | 23 Jul 2015 |
| United Kingdom | 277300 | Fife K | London Central Research Ethics Committee | 24 Jun 2016 |
| United Kingdom | 277304 | Wagstaff J | London Central Research Ethics Committee | 27 Oct 2015 |
| United Kingdom | 277410 | Porfiri E | London Central Research Ethics Committee | 01 Dec 2015 |
| United Kingdom | 277413 | Parikh O | London Central Research Ethics Committee | 07 Apr 2016 |

| Country | Site No. | Principal Investigator | EC/IRB | Approved Date |
|---|---|---|---|---|
| United Kingdom | 278404 | Wheater M | London Central Research Ethics Committee | 21 Dec 2015 |
| United Kingdom | 280523 | Powles T | London Central Research Ethics Committee | 27 Aug 2015 |
| United Kingdom | 280525 | Protheroe A | London Central Research Ethics Committee | 24 Sep 2015 |
| United Kingdom | 281159 | Turajlic S | London Central Research Ethics Committee | 24 Sep 2015 |
| United States | 278807 | Lam E | Western Institutional Review Board | 17 Jul 2015 |
| United States | 278808 | Rini B | Cleveland Clinic Florida; Cleveland Clinic Instituaitonal Review Board | 22 Apr 2015 |
| United States | 278810 | Alter R | Western Institutional Review Board | 02 Apr 2015 |
| United States | 278811 | Stadler W | The University of Chicago IRB | 22 Apr 2015 |
| United States | 278812 | Rathmell K | Vanderbilt University Institutional Review Board | 31 Mar 2015 |
| United States | 278815 | Fong L | Western Institutional Review Board | 16 Sep 2015 |
| United States | 278816 | Motzer R | Memorial Sloan Kettering Cancer Center; Institutional Review Board | 10 Jun 2015 |
| United States | 278819 | Rezazadeh Kalebasty A | Western Institutional Review Board | 05 Jun 2015 |
| United States | 278821 | Assikis V | Copernicus Group IRB | 01 May 2015 |
| United States | 278943 | Tykodi S | Western Institutional Review Board | 07 Aug 2015 |
| United States | 282098 | Choueiri T | Dana Farber Cancer Institute Institutional Review Board (278993 is main site) | 14 Jul 2015 |
| United States | 282099 | Choueiri T | Dana Farber Cancer Institute Institutional Review Board (278993 is main site) | 14 Jul 2015 |

| Country | Site No. | Principal Investigator | EC/IRB | Approved Date |
|---|---|---|---|---|
| United States | 278993 | Choueiri T | Dana Farber Cancer Institute Institutional Review Board | 14 Jul 2015 |
| United States | 278995 | Sundararajan S | Western Institutional Review Board | 04 Jun 2015 |
| United States | 279354 | Hainsworth J | Western Institutional Review Board | 13 May 2015 |
| United States | 279682 | Fruehauf J | UC Irvine office of Research Administration | 10 Jul 2015 |
| United States | 280241 | Schnadig I | US Oncology, Inc Institutional Review Board | 13 Apr 2015 |
| United States | 280364 | Vogelzang N | US Oncology, Inc Institutional Review Board | 13 Apr 2015 |
| United States | 280425 | Page R | Western Institutional Review Board | 01 Jul 2015 |
| United States | 280533 | Kochenderfer M | US Oncology, Inc Institutional Review Board | 13 Apr 2015 |
| United States | 280602 | Shaffer D | US Oncology, Inc Institutional Review Board | 13 Apr 2015 |
| United States | 281294 | Cultrera J | Western Institutional Review Board | 22 May 2015 |
| United States | 281295 | Burke J | US Oncology, Inc Institutional Review Board | 13 Apr 2015 |
| United States | 281296 | Herms B | US Oncology, Inc Institutional Review Board | 13 May 2015 |
| United States | 281343 | Arrowsmith E | Western Institutional Review Board | 04 Jun 2015 |
| United States | 281556 | Percent I | Western Institutional Review Board | 13 May 2015 |
| United States | 281900 | Koletsky A | Copernicus Group IRB | 28 Sep 2015 |
| United States | 282279 | Hutson.T | US Oncology, Inc Institutional Review Board | 13 Apr 2015 |
