## Supplementary material for "Allelic Variation in *HLA-DRB1* is Associated with Development of Anti-Drug Antibodies in Cancer Patients Treated with Atezolizumab that are Neutralizing *in Vitro*": Ethics Committee approvals: IMpassion130 EC IRB.pdf

| Site # | Investigator | EC/IRB Name and Address | IRB/EC Approval |
|---|---|---|---|
| 274488 | Neron, Yeni Veronica | Centro de Pesquisas Oncologicas - CEPON; Comite de Etica em Pesquisa - CEPON, Rodovia Admar Gonzaga, SC 655, km 0,5, Itacorubi, 88034-000, Florianopolis, SC, BRAZIL | 10-Apr-2016 |
| 274528 | Liedke, Pedro Emanuel | Comite de Etica em Pesquisa do; HCPA, Rua Ramiro Barcelos, 2350, 90035-903, Porto Alegre - RS, RS, BRAZIL | 05-Mar-2016 |
| 274560 | Andrade, Livia | Hospital Santa Izabel - Santa Casa de Misericordia da Bahia | 30-Apr-2016 |
| 274562 | Barrios, Carlos Henrique | Comitê de Ética em Pesquisa da PUCRS, Avenida Ipiranga, 6690 – 3º andar – Sala 314, 90610-000, Porto Alegre, RS, BRAZIL | 18-Apr-2016 |
| 274604 | MELICHAR, BOHUSLAV | Eticka Komise Fakultni Nemocnice Olomouc, I.P. PAVLOVA 6, 775 20, OLOMOUC, CZECH REPUBLIC | 09-May-2016 |
| 274611 | Kubiatowski, Tomasz | Komisja Bioetyczna przy Instytucie Centrum Onkologii w Warszawie , UL. ROENTGENA 5, 02-781, WARSZAWA, POLAND | 10-Nov-2016 |
| 274612 | Nowecki, Zbigniew | Komisja Bioetyczna przy Instytucie Centrum Onkologii w Warszawie , UL. ROENTGENA 5, 02-781, WARSZAWA, POLAND | 22-Mar-2016 |
| 274615 | CHMIELOWSKA, EWA | Komisja Bioetyczna przy Instytucie Centrum Onkologii w Warszawie , UL. ROENTGENA 5, 02-781, WARSZAWA, POLAND | 22-Mar-2016 |
| 274648 | Oakman, Catherine | Peter MacCallum Cancer Centre Ethics Committee, 305 Grattan Street, 3000, Melbourne, Victoria, AUSTRALIA | 29-May-2015 |
| 274649 | Loi, Sherene | Peter MacCallum Cancer Centre Ethics Committee, 305 Grattan Street, 3000, Melbourne, Victoria, AUSTRALIA | 29-May-2015 |
| 274652 | Cuff, Kate | Peter MacCallum Cancer Centre Ethics Committee, 305 Grattan Street, 3000, Melbourne, Victoria, AUSTRALIA | 29-May-2015 |
| 274653 | McCarthy, Nicole | Bellberry Human Research Ethics Committee, Bellberry Limited, 129 Glen Osmond Road, 5063, Eastwood, South Australia, AUSTRALIA | 01-Apr-2015 |
| 274654 | Tsoi, Daphne | St John of God Health Care Ethics Committee, National Office, 175 Cambridge Street, 6904, Subiaco, WA, AUSTRALIA | 08-Apr-2015 |
| 274658 | HUANG, CHIUN-SHENG | Research Ethics Committee, Nat. Taiwan Univ. Hosp., 7 CHUNG-SHAN SOUTH ROAD , 100, TAIPEI, TAIWAN | 24-Apr-2016 |
| 274659 | Tseng, Ling - Ming | Institutional Review Board, Taipei Veterans General Hospital, No.201, Sec. 2, Shipai Rd., Beitou District, 11217, Taipei, TAIWAN | 22-Mar-2016 |
| 274661 | JUNG, KYUNG HAE | Asan Medical Center Ethics Committee, 88, Olympic-ro 43-gil, Songpa-gu, 05505, Seoul, KOREA, REPUBLIC OF | 16-Mar-2015 |
| 274662 | IM, SEOCK AH | Seoul National Univ. Ethics Comittee, 101, Daehak-ro, Jongno-gu, Seoul, Korea, 03080, Seoul, KOREA, REPUBLIC OF | 27-Mar-2015 |
| 274663 | SOHN, JOO-HYUK | SeveranceHospital- YonseiUniversity; IRB, 2F, GwangHye-gwan, 50-1, Yonsei-ro, Seodaemun-gu, 03722, Seoul, KOREA, REPUBLIC OF | 27-Mar-2015 |
| 274664 | IM, YOUNG-HYUCK | Samsung Medical Center EC, 81, Irwon-ro, Gangnam-gu, 06351, Seoul, KOREA, REPUBLIC OF | 27-Mar-2015 |
| 274685 | Gomes, Jessica | CEP da Real e Benemérita Associação Portuguesa de Beneficência/SP, Rua Maestro Cardim 769 bloco V -Térreo, Bela Vista, 01323-900, São Paulo, SP, BRAZIL | 28-Apr-2016 |
| 274707 | Kuzmin, Alexey | E. C. of SI of Healthcare Kazan Oncology Dispensary, SIBIRSKY TRAKT, 29, 420029, KAZAN, RUSSIAN FEDERATION | 09-Dec-2016 |

| | | | |
|---|---|---|---|
| 274708 | GOTOVKIN, Evgeny | Ivanovo Regional Clinical Oncology Dispensary Ethics Committee, Lyubimova str.,5, 153040, Ivanovo, RUSSIAN FEDERATION | 29-Jun-2016 |
| 274709 | Nechaeva, Marina | LEC of Arkhangelsk' regional clinical oncology dispensary, 145a pr Obvodny kanal, 163045, Arkhangelsk, RUSSIAN FEDERATION | 30-May-2016 |
| 274711 | Stroyakovskii, Daniil | EC of Moscow City Oncol. Hospital #62, p/o Stepanovskoe, Krasnogorsk district, Moscow Region, 143423, Mosow, RUSSIAN FEDERATION | 29-Jun-2016 |
| 274793 | Varela, Mirta | Comité de Ética del Centro Oncológico y de Investigaciones Buenos Aires (CECOIBA), Calle #12 Nº 4756, Berazategui, B1884BBF, Buenos Aires, ARGENTINA | 22-Jun-2016 |
| 274794 | LERZO, GUILLERMO | Comite de Etica en Investigacion Clinica (CEIC), Larrea 1381 3° A, C1117ABK, Ciudad Autonoma de Buenos Aires, ARGENTINA | 07-Jan-2016 |
| 274887 | Dirix, Luc | Institut Jules Bordet, Comité d'Ethique | 08-Jun-2015 |
| 274914 | Gombos, Andrea | Institut Jules Bordet, Comité d'Ethique | 08-Jun-2015 |
| 275013 | Beuselinck, Benoit | Institut Jules Bordet, Comité d'Ethique | 08-Jun-2015 |
| 275337 | Dent, Susan | Ontario Cancer Research Ethics Board, 661 University Avenue, Suite 510, M5G 0A3, Toronto, Ontario, CANADA | 15-Sep-2015 |
| 275339 | CHIA, STEPHEN | UBC BCCA Research Ethics Board (BCCA REB), 902-750 West Broadway, Fairmont Medical Building, V5Z 1H8, Vancouver, British Columbia, CANADA | 07-Dec-2015 |
| 275340 | PROVENCHER, LOUISE | Comite d'ethique de la recherche, Hopital Saint Sacrement du CHA, 10,rue de l'Espinay-Room A0-124, G1S 3L5, Quebec, Quebec, CANADA | 21-Jan-2016 |
| 275341 | JOY, ANIL ABRAHAM | Health Research Ethics Board of Alberta, 1500, 10140 - 103 Avenue NW, T5J-1V3, Edmonton, Alberta, CANADA | 27-Jul-2015 |
| 275342 | RAYSON, DANIEL | Nova Scotia Health Authority Research Ethics Board, QEII Health Science Centre for Clinical Res, Room 118-5790 University Avenue, B3H 1V7, Halifax, Nova Scotia, CANADA | 29-Jul-2015 |
| 275343 | Ferrario, Cristiano | Comite d'ethique de la recherche, Hopital Saint Sacrement du CHA, 10,rue de l'Espinay-Room A0-124, G1S 3L5, Quebec, Quebec, CANADA | 21-Jan-2016 |
| 275345 | Clarke, Kaethe | UBC BCCA Research Ethics Board (BCCA REB), 902-750 West Broadway, Fairmont Medical Building, V5Z 1H8, Vancouver, British Columbia, CANADA | 22-May-2016 |
| 275347 | Koneru, Rama | Lakeridge Health Research Ethics Board, 1 HOSPITAL COURT, L1G 2B9, OSHAWA, Ontario, CANADA | 30-Jul-2015 |
| 275583 | Rody, Achim | EK Lübeck, Ratzeburger Allee 160, 23538, Lübeck, GERMANY | 11-May-2015 |
| 275584 | Hoffmann, Oliver | EK Essen Ethik-Kommission der Medizinischen Fakultät der Universität Duisburg-Essen, Robert-Koch-Str. 9-11, 2. Stock, 45147, Essen, GERMANY | 11-May-2015 |
| 275585 | Mahlberg, Rolf | Ethikkommission Landesärztekammer Rheinland-Pfalz, Deutschhausplatz 3, 55116, Mainz, GERMANY | 11-May-2015 |
| 275599 | Warner, Ellen | Ontario Cancer Research Ethics Board, 661 University Avenue, Suite 510, M5G 0A3, Toronto, Ontario, CANADA | 09-Sep-2015 |
| 275649 | Wimberger, Pauline | Ethik-Kommission am Universitätsklinikum Carl-Gustav-Carus Technische Universität Dresden, Fetscherstrasse 74, 01307, Dresden, GERMANY | 11-May-2015 |
| 275650 | Ettl, Johannes | Ethikkommission Technische Universität München, Fakultät für Medizin, Ismaninger Str. 22, 81675, München, GERMANY | 11-May-2015 |
| 275653 | Grischke, Eva-Maria | EK an der Med. Fakultät d. Eberhard-Karls-Uni und am Uniklinikum Tübingen, Gartenstr. 47, 72074, Tübingen, GERMANY | 11-May-2015 |
| 275655 | Schneeweiss, Andreas | Ethikkommission der Medizinischen Fakultät Heidelberg, Alte Glockengießerei 11/1, 69115, Heidelberg, GERMANY | 11-May-2015 |

| | | | |
|---|---|---|---|
| 275656 | Weide, Rudolf | Ethikkommission Landesärztekammer Rheinland-Pfalz, Deutschhausplatz 3, 55116, Mainz, GERMANY | 11-May-2015 |
| 275717 | Fasching, Peter | Ethik-Kommission der Medizinischen Fakultät der Friedrich-Alexander-Universität Erlangen-Nürnberg, Krankenhausstr. 12, 91054, Erlangen, GERMANY | 11-May-2015 |
| 275837 | WARDLEY, ANDREW | NRES Committee London - City and East; Bristol Research Ethics Committee Centre, Whitefriars, Level 3, Block B, Lewins Mead, Bristol, BS1 2NT, UNITED KINGDOM | 08-Oct-2015 |
| 275917 | Harries, Mark | NRES Committee London - City and East; Bristol Research Ethics Committee Centre, Whitefriars, Level 3, Block B, Lewins Mead, Bristol, BS1 2NT, UNITED KINGDOM | 07-Aug-2015 |
| 275918 | CAMERON, DAVID | NRES Committee London - City and East; Bristol Research Ethics Committee Centre | 08-Jul-2015 |
| 276391 | Telli, Melinda | Stanford Research Compliance Office, 3000 El Camino Real, Five Palo Alto Square, 4th Floor, Palo Alto, CA, 94306, UNITED STATES | 10-Jun-2015 |
| 276394 | Pohlmann, Paula | MedStar Health Research Institute-Georgetown Univ. Oncology IRB, 3900 Reservoir Rd NW, MedDent SW104, Washington, DC, 20057, UNITED STATES | 22-Apr-2015 |
| 276395 | Abramson, Vandana | Vanderbilt University Institutional Review Board, 1313 21st Ave. South, 504 Oxford House, Nashville, TN, 37232-4315, UNITED STATES | 03-Aug-2015 |
| 276396 | Adams, Sylvia | NYU Langone Medical Center IRB, 1 Park Avenue, 6th Floor, New York, NY, 10016, UNITED STATES | 12-Aug-2015 |
| 276398 | Rugo, Hope S. | UCSF Committee on Human Research, 3333 California St., Suite 315, SAN FRANCISCO, CA, 94118, UNITED STATES | 11-Dec-2015 |
| 276403 | Ward, Patrick | US Oncology IRB, 10101 Woodloch Forest Drive, The Woodlands, TX, 77380, UNITED STATES | 18-Sep-2015 |
| 276407 | Hofstatter, Erin | Yale University Human Research Protection Program, 55 College Street, New Haven, CT, 06510, UNITED STATES | 14-Sep-2015 |
| 276411 | Litton, Jennifer | UT MD Anderson Cancer Centre; UT MDACC IRB #1, 7007 Bertner Ave., Unit 1637, HOUSTON, TX, 77030, UNITED STATES | 08-Jul-2015 |
| 276420 | DE LAURENTIIS, MICHELINO | Comitato Etico IRCCS Pascale<br>, VIA M. SEMMOLA 1, 80131, NAPOLI, Campania, ITALY | 17-Feb-2016 |
| 276421 | Colleoni, Marco Angelo | Comitato Etico Istituto Europeo Oncologico, Via Ripamonti 435, 20141 Milano, Italy | 23-Mar-2016 |
| 276891 | PATEL, TARAL | QUORUM REVIEW IRB, 1501 Fourth Ave., Suite 800, Seattle, WA, 98101, UNITED STATES | 05-May-2015 |
| 276914 | Suga, Jennifer | Kaiser Permanente Northern California IRB, 1800 Harrison Street, 16th Floor, Oakland, CA, 94612, UNITED STATES | 16-Jun-2015 |
| 276956 | TANNER, MINNA | Pirkanmaan sairaanhoitopiirin eettinen toimikunta<br>PL 2000, Tampere 33521, Finland | 03-May-2016 |
| 277114 | Moezi, Mehdi | QUORUM REVIEW IRB, 1501 Fourth Ave., Suite 800, Seattle, WA, 98101, UNITED STATES | 05-May-2015 |
| 277528 | Ibrahim, Emad | QUORUM REVIEW IRB, 1501 Fourth Ave., Suite 800, Seattle, WA, 98101, UNITED STATES | 05-May-2015 |
| 277747 | Puhalla, Shannon | Quorum, 1501 Fourth Avenue, Suite 800, SEATTLE, WA, 98101, UNITED STATES | 16-Dec-2015 |
| 277748 | Oyola, Raul H. | QUORUM REVIEW IRB, 1501 Fourth Ave., Suite 800, Seattle, WA, 98101, UNITED STATES | 20-May-2015 |
| 278058 | Smith, Wade | QUORUM REVIEW IRB, 1501 Fourth Ave., Suite 800, Seattle, WA, 98101, UNITED STATES | 05-May-2015 |
| 278059 | Conlin, Alison | Providence Health System IRB, 5251 NE Glisan Street, Portland, OR, 97213, UNITED STATES | 09-Jul-2015 |
| 278136 | Nagpal, Sunil | West Michigan Cancer Center IRB, 200 North Park Street, Kalamazoo, MI, 49007, UNITED STATES | 24-Apr-2015 |

| | | | |
|---|---|---|---|
| 278200 | Emens, Leisha | Johns Hopkins Medicine Institutional Review Board, 1620 McElderry Street, Reed Hall B-130, Baltimore, MD, 21205-1911, UNITED STATES | 01-Jul-2015 |
| 278255 | Weise, Amy | Wayne State University Human Investigation committee, 87 East Canfield, Second Floor, Detroit, MI, 48201, UNITED STATES | 17-Aug-2015 |
| 278296 | Dubey, Sidharth | NRES Committee London - City and East; Bristol Research Ethics Committee Centre, Whitefriars, Level 3, Block B, Lewins Mead, Bristol, BS1 2NT, UNITED KINGDOM | 22-Jul-2015 |
| 278330 | POLIKOFF, JONATHAN | Kaiser Permanente Southern California, 393 East Walnut Street, 2nd Floor, Pasadena, CA, 91188, UNITED STATES | 19-May-2015 |
| 278580 | TAN-CHIU, ELIZABETH | QUORUM REVIEW IRB, 1501 Fourth Ave., Suite 800, Seattle, WA, 98101, UNITED STATES | 05-May-2015 |
| 278610 | Wilkinson, Mary | QUORUM REVIEW IRB, 1501 Fourth Ave., Suite 800, Seattle, WA, 98101, UNITED STATES | 05-May-2015 |
| 278643 | RISEBERG, DAVID | Mercy Medical Center IRB, 345 St. Paul Place, 7th Floor, Baltimore, MD, 21202, UNITED STATES | 07-Apr-2015 |
| 278647 | Frank, Richard | Norwalk Hospital IRB, 34 Maple Street, Norwalk, CT, 06856, UNITED STATES | 11-May-2015 |
| 278725 | Schmid, Peter | NRES Committee London - City and East; Bristol Research Ethics Committee Centre, Whitefriars, Level 3, Block B, Lewins Mead, Bristol, BS1 2NT, UNITED KINGDOM | 16-Jun-2015 |
| 278911 | Forstbauer, Helmut | EK der Ärztekammer Nordrhein, Tersteegenstr. 9, 40474, Düsseldorf, GERMANY | 11-May-2015 |
| 278912 | Peters, Uwe | Ethik-Kommission Berlin Landesamt für Gesundheit und Soziales, Fehrbelliner Platz 1, 10707, Berlin, GERMANY | 11-May-2015 |
| 278913 | Müller, Volkmar | Ethik-Kommission der Ärztekammer Hamburg, Humboldtstr. 67a, 22083, Hamburg, GERMANY | 11-May-2015 |
| 278915 | Liersch, Rüdiger | Ethik-Kommission der Ärztekammer Westfalen-Lippe und der Medizinischen Fakultät der WWU Münster, Gartenstr. 210 - 214, 48147, Münster, GERMANY | 11-May-2015 |
| 278938 | Schumacher, Claudia | EK der Ärztekammer Nordrhein, Tersteegenstr. 9, 40474, Düsseldorf, GERMANY | 11-May-2015 |
| 279333 | Hegewisch-Becker, Susanna | Ethik-Kommission der Ärztekammer Hamburg, Humboldtstr. 67a, 22083, Hamburg, GERMANY | 11-May-2015 |
| 279472 | Kono, Scott | Kaiser Permanente Colorado Institutional Review Board, 10065 East Harvard Avenue, Suite 300, Denver, CO, 80231, UNITED STATES | 16-Jun-2015 |
| 279724 | Rakowski, Thomas | WIRB, 1019 39th Avenue SE, Suite 120, Puyallup, WA, 98374-2115, UNITED STATES | 22-Jun-2015 |
| 279725 | Young, Tammy H. | Quorum, 1501 Fourth Avenue, Suite 800, SEATTLE, WA, 98101, UNITED STATES | 05-Apr-2016 |
| 279726 | Vidal, Greg | WIRB, 1019 39th Avenue SE, Suite 120, Puyallup, WA, 98374-2115, UNITED STATES | 01-Mar-2016 |
| 280430 | Hamilton, Erika | WIRB, 1019 39th Avenue SE, Suite 120, Puyallup, WA, 98374-2115, UNITED STATES | 22-Jul-2015 |
| 280441 | Sanchez-Rivera, Ines | US Oncology, Inc Institutional Review Board, 10101 Woodloch Forest, The Woodlands, TX, 77380, UNITED STATES | 21-May-2015 |
| 280553 | Wright, Gail Lynn | WIRB, 1019 39th Avenue SE, Suite 120, Puyallup, WA, 98374-2115, UNITED STATES | 03-Sep-2015 |
| 280554 | Hart, Lowell | WIRB, 1019 39th Avenue SE, Suite 120, Puyallup, WA, 98374-2115, UNITED STATES | 14-Aug-2015 |
| 280676 | Lo, K. M. Steve | QUORUM REVIEW IRB, 1501 Fourth Ave., Suite 800, Seattle, WA, 98101, UNITED STATES | 16-Jun-2015 |
| 280677 | EDENFIELD, WILLIAM | WIRB, 1019 39th Avenue SE, Suite 120, Puyallup, WA, 98374-2115, UNITED STATES | 21-May-2015 |
| 280957 | Krill-Jackson, Elisa | Mount Sinai Medical Center IRB, 4300 Alton Road, Miami Beach, FL, 33140, UNITED STATES | 21-Sep-2015 |

| | | | |
|---|---|---|---|
| 280958 | Sparano, Joseph | Biomedical Research Alliance of New York, LLC, 1981 Marcus Avenue, Suite 210, Lake Success, NY, 11042, UNITED STATES | 14-Jul-2015 |
| 286952 | WILCKEN, NICHOLAS | Peter MacCallum Cancer Centre Ethics Committee, ST ANDREWS PLACE, 3002, EAST MELBOURNE, Victoria, AUSTRALIA | 01-Sep-2015 |
| 289579 | Shao, Ryan | Wellmont Health System Institutional Review Board, 105 W. Stone Drive, Suite 6A, Kingsport, TN, 37660, UNITED STATES | 08-Dec-2015 |
| 289580 | Forero, Andres | WIRB, 1019 39th Avenue SE, Suite 120, Puyallup, WA, 98374-2115, UNITED STATES | 14-Jun-2016 |
| 289743 | STEGER, GUENTHER | Ethikkommission der Universität Wien /AKH, Borschkegasse 8b/E 06, 1090, Wien, AUSTRIA | 13-May-2016 |
| 289744 | Stoeger, Herbert | Ethikkommission der Universität Wien /AKH, Borschkegasse 8b/E 06, 1090, Wien, AUSTRIA | 13-May-2016 |
| 289745 | PETZER, ANDREAS | Ethikkommission der Universität Wien /AKH, Borschkegasse 8b/E 06, 1090, Wien, AUSTRIA | 13-May-2016 |
| 289771 | Canon, Jean-Luc | Institut Jules Bordet, Comité d'Ethique | 02-Mar-2016 |
| 289772 | Borms, Marleen | Institut Jules Bordet, Comité d'Ethique | 02-Mar-2016 |
| 289773 | Duhoux, Francois | Institut Jules Bordet, Comité d'Ethique | 02-Mar-2016 |
| 289794 | Turna, Hande | Hacettepe University Ethics Committee; Ethics Committtee | 31-Mar-2016 |
| 289795 | Paydas, Semra | Hacettepe University Ethics Committee; Ethics Committtee | 31-Mar-2016 |
| 289799 | Thomssen, Christoph | Ethikkommission der Martin-Luther-Universität Halle-Wittenberg, Magdeburger Str. 16, 06112, Halle (Saale), GERMANY | 10-Dec-2015 |
| 289801 | USLU, RUCHAN | Hacettepe University Ethics Committee; Ethics Committtee | 31-Mar-2016 |
| 289802 | Aksoy, Sercan | Hacettepe University Ethics Committee; Ethics Committtee | 31-Mar-2016 |
| 289804 | Murtezani, Zafir | Clinical Hospital Center Bezanijska kosa; Ethics Committee Clinical Hospital Center Bezanijska kosa, Bezanijska kosa bb, 11000, Belgrade, SERBIA | 07-Apr-2016 |
| 289932 | Leung, Roland | HKU/HA HKW IRB, Room 901, Adminstration Block, 102 Pok Fu Lam Road, Queen Mary Hospital, Hong Kong, HONG KONG | 14-Apr-2016 |
| 290056 | Papazisis, Konstantinos | National Ethics Committee | 25-Apr-2016 |
| 290057 | KALOFONOS, HARALABOS | National Ethics Committee | 25-Apr-2016 |
| 290059 | Mavroudis, Dimitris | National Ethics Committee | 25-Apr-2016 |
| 290060 | Katsaounis, Panagiotis | National Ethics Committee | 25-Apr-2016 |
| 290061 | Ardavanis, Alexandros | National Ethics Committee | 25-Apr-2016 |
| 290063 | Jernling, Martin | Regionala Etikprovningsnamnden i Uppsala | 04-Apr-2016 |
| 290067 | Kersten, Christian | REK Vest; Universitetet i Bergen, Det Medisinske Fakultet | 22-Feb-2016 |
| 290068 | Gilje, Bjornar | REK Vest; Universitetet i Bergen, Det Medisinske Fakultet | 22-Feb-2016 |
| 290069 | Molvaer, Sindre | REK Vest; Universitetet i Bergen, Det Medisinske Fakultet | 22-Feb-2016 |
| 290103 | Webster, Marc | Health Research Ethics Board of Alberta, 1500, 10140 - 103 Avenue NW, T5J-1V3, Edmonton, Alberta, CANADA | 16-Feb-2016 |
| 290104 | Robinson, Andrew | Ontario Cancer Research Ethics Board, 661 University Avenue, Suite 510, M5G 0A3, Toronto, Ontario, CANADA | 11-Mar-2016 |
| 290128 | Chang, Jenny | The Methodist Hospital Research Institute IRB, 6565 Fannin Street, MGJ4-010, Houston, TX, 77030, UNITED STATES | 09-Aug-2016 |
| 290186 | (Sirisinha) Dejthevaporn, THITIYA | Ethical Clearance Committee on Human Rights, 270 RamaVI Road. Faculty of Medicine, Ramathibodi Hospital, Phayathai Rajathevi Bangkok 10400, 10400, Bangkok, THAILAND | 15-Mar-2016 |

| | | | |
|---|---|---|---|
| 290187 | Soparattanapaisarn, Nopadol | Ethics Committee, Faculty of Medicine, Siriraj Hospital, Mahidol University, 10700, Bangkok, THAILAND | 07-Apr-2016 |
| 290188 | Dechaphunkul, Arunee | Office of Human Research Ethics Committee (HREC; Faculty of Medicine, Prince of Songkla University,, 15 Karnjanavanit Road, Hat Yai, 90110, Songkla,THAILAND, 90110, Hat Yai, THAILAND | 09-Jun-2016 |
| 290200 | WANG, HWEI-CHUNG | Research Ethics Committee China Medical University & Hospital, 2 Yude Road, 40447, Taichung, TAIWAN | 22-Apr-2016 |
| 290201 | HOU, MING-FENG | Kaohsiung Medical University Hospital, IRB, No.100, Tzyou 1st Rd., 807, Kaohsiung, TAIWAN | 12-May-2016 |
| 290294 | ROCHLITZ, CHRISTOPH | Ethikkommission Nordwest- und Zentralschweiz (EKNZ) | 13-May-2016 |
| 290295 | Weder, Patrik | Ethikkommission Ostschweiz (EKOS) | 13-May-2016 |
| 290296 | Petrausch, Ulf | Kantonale Ethikkommission Zürich (KEK), Kantonale Ethikkommission, Stampfenbachstrasse 121, 8090, Zürich, SWITZERLAND | 19-May-2016 |
| 290310 | BESLIJA, SEMIR | Ethic committee of Clinical Center University of Sarajevo | 04-May-2016 |
| 290316 | Ponomareva, Olha | Ethics Committee of Kyiv City Oncological Hospital, Verkhovynna street, 69, 03115, Kyiv, UKRAINE | 24-Jun-2016 |
| 290317 | BONDARENKO, IGOR | Ethics Committee of Dnipropetrovsk City Multilat. Clinical Hospital №4, Blyzhnya street, 31, 49102, Dnipropetrovsk, UKRAINE | 08-Jul-2016 |
| 290318 | Shparyk, Yaroslav | Local Ethics Committee of Lviv State Reg. Oncol. Med. Diagn. Centre, Gasheka street, 2A, 79031, Lviv, UKRAINE | 15-Jun-2016 |
| 290321 | Sinielnikov, Ivan | Ethics Committee of Treatment and Preventive Institution "Volyn Regional Oncology Dispensary", 1, Timiriazeva str., 43018, Lutsk, VOLHYNIAN GOVERNORATE, UKRAINE | 22-Aug-2016 |
| 290324 | Moiseenko, Vladimir | S-Pb clinical scientific practical center of specialized kinds of medical care (oncological); Center, Pesochniy, Leningradskaya str, 68a, lit A, Saint-Petersburg, RUSSIAN FEDERATION | 03-Jun-2016 |
| 290325 | SMOLIN, ALEXEY | Ethics Committee of the Main Military Clinical Hospital n.a. N.N.Burdenko, 3 Gospitalnaya square, 105229, Moscow, RUSSIAN FEDERATION | 22-Jun-2016 |
| 290336 | Bilici, Ahmet | Hacettepe University Ethics Committee; Ethics Committtee | 31-Mar-2016 |
| 290347 | Franke, Fabio Andre | Comitê de Ética em Pesquisa; Universidade Regional do Noroeste do Estado do Rio Grande do Sul UNIJUÍ, Rua do Comércio, 3000, Subsolo do Prédio da Biblioteca – Sala 06, 98700-000, Ijuí, RS, BRAZIL | 18-Apr-2016 |
| 290348 | Hegg, Roberto | Comite de Etica em Pesquisa do Centro de Referencia da Saude da Mulher, Avenida Brigadeiro Luis Antonio, 683, Segundo Andar - Bela Vista, 01317-000, São Paulo, SP, BRAZIL | 04-Apr-2016 |
| 290349 | Santos Borges, Giuliano | Comite de Etica do Hospital de Clinicas de Porto Alegre | 20-Apr-2016 |
| 290357 | Silva, Eduardo Henrique | Comite de Etica em Pesquisa Seres Humanos da Universidade Federal do Ceara | 07-Apr-2016 |
| 290371 | Kaen, Diego Lucas | Comite de Etica en Investigacion Clinica (CEIC), Larrea 1381 3° A, C1117ABK, Ciudad Autonoma de Buenos Aires, ARGENTINA | 07-Jan-2016 |
| 290401 | Castillo, Omar | Instituto Conmemorativo Gorgas de Estudios de la Salud Comité de Bioética de la investigación | 10-Aug-2016 |
| 290402 | CASTRO-SALGUERO, HUGO | Comité de Ética Independiente Zugueme, 3a Calle 11-36, Zona 15, 01015, Guatemala, GUATEMALA | 24-Feb-2016 |
| 290403 | Corrales Rodriguez, Luis | Comite Etico Cientifico Universidad de Ciencias Medicas, Sabana Norte, del ICE 200 metros noreste, edificio esquinero, primer piso, San José, COSTA RICA | 14-Jul-2016 |
| 290405 | VILLALOBOS VALENCIA, RICARDO | Comité de Ética en Investigación Sanatorio Alcocer Pozo S.A de C.V. | 09-May-2016 |

| | | | |
|---|---|---|---|
| 290413 | Streb, Joanna | Komisja Bioetyczna przy Instytucie Centrum Onkologii w Warszawie , UL. ROENTGENA 5, 02-781, WARSZAWA, POLAND | 21-Jun-2016 |
| 290415 | LESNIEWSKI-KMAK, KRZYSZTOF | Komisja Bioetyczna przy Instytucie Centrum Onkologii w Warszawie , UL. ROENTGENA 5, 02-781, WARSZAWA, POLAND | 22-Mar-2016 |
| 290416 | PRAUSOVA, JANA | Eticka Komise pro Multicentricka klinicka hodnoceni; Fakultni Nemocnice v Motole, V Uvalu 84, 150 06, Praha 5 - Motol, CZECH REPUBLIC | 30-Mar-2016 |
| 290417 | PADRIK, PEETER | Tallinn Medical Research Ethics Committee, National Institute for Health Development, Hiiu 42, 11619, Tallinn, ESTONIA | 14-Apr-2016 |
| 290418 | Valvere, Vahur | Tallinn Medical Research Ethics Committee, National Institute for Health Development, Hiiu 42, 11619, Tallinn, ESTONIA | 17-Mar-2016 |
| 290420 | Rubovszky, Gabor | Medical Research Council, Ethics Committee for Clinical Pharmacology, Arany J. u. 6-8., 1051, Budapest, HUNGARY | 03-May-2016 |
| 290421 | Kahan, Zsuzsanna | Medical Research Council, Ethics Committee for Clinical Pharmacology, Arany J. u. 6-8., 1051, Budapest, HUNGARY | 03-May-2016 |
| 290422 | Landherr, Laszlo | Medical Research Council, Ethics Committee for Clinical Pharmacology, Arany J. u. 6-8., 1051, Budapest, HUNGARY | 03-May-2016 |
| 290423 | Bitina, Marianna | Ethics Committee for Clinical Trials of Medicinal Products, Aizkraukles Street 21-113, LV-1006, RIga, LATVIA | 15-Apr-2016 |
| 290425 | Hegmane, Alinta | Ethics Committee for Clinical Trials of Medicinal Products, Aizkraukles Street 21-113, LV-1006, RIga, LATVIA | 15-Apr-2016 |
| 290427 | Borstnar, Simona | Republic of Slovenia National Medical Ethics Committee, Stefanova 5, 1000, Ljubljana, SLOVENIA | 05-May-2016 |
| 290431 | MORALES-VASQUES, FLAVIA | Comite de Etica en Investigacion de Mexico Centre for Clinical Research SA de CV, Amores No. 709, Col. del Valle, 03100, Distrito Federal, MEXICO | 15-Mar-2016 |
| 290495 | GANJU, VINOD | Bellberry Human Research Ethics Committee, Bellberry Limited, 129 Glen Osmond Road, 5063, Eastwood, South Australia, AUSTRALIA | 29-Feb-2016 |
| 290501 | Duchnowska, Renata | Komisja Bioetyczna przy Instytucie Centrum Onkologii w Warszawie , UL. ROENTGENA 5, 02-781, WARSZAWA, POLAND | 21-Jun-2016 |
| 290504 | Negru, Mircea Serban | Comisia Nationala de Bioetica a Medicamentului si a Dispozitivelor Medicale- Sos , Sos. Stefan cel Mare nr. 19-21, Pavilion K, sector 2, 020125, Bucuresti, ROMANIA | 21-Jul-2016 |
| 290511 | Avila Zamora, Oscar Nicolas | Instituto Estatal de Cancerologia de Colima; Comite de Etica en Investigacion | 01-Jul-2016 |
| 290553 | Juarez Ramiro, Alejandro | Comite de Etica en Investigacion de Mexico Centre for Clinical Research SA de CV, Amores No. 709, Col. del Valle, 03100, Distrito Federal, MEXICO | 15-Mar-2016 |
| 290563 | Wong, Andrea | SingHealth Centralised IRB; Review Board B, Singapore Health Services Pte Ltd, 168 Jalan Bukit Merah, #06-08 Tower 3 Connection One, 150168, Singapore, SINGAPORE | 22-Mar-2016 |
| 290565 | Dent, Rebecca | SingHealth Centralised IRB; Review Board B, Singapore Health Services Pte Ltd, 168 Jalan Bukit Merah, #06-08 Tower 3 Connection One, 150168, Singapore, SINGAPORE | 22-Mar-2016 |
| 290589 | Franco, Sandra | Comite De Etica De Investigacion Clinica | 10-Aug-2016 |
| 290615 | Yanez, Eduardo | Comité de Ética Cientifico del Servicio de Salud Araucania Sul, Vicuña Mackenna, 51, 4781086, Temuco, CHILE | 19-Apr-2016 |
| 290708 | Ciruelos Gil, Eva | CEIC Parc de Salut Mar; IMIM- Hospital del Mar, C/ Dr. Aiguader, 88, Planta 1, 08003, Barcelona, BARCELONA, SPAIN | 08-Mar-2016 |

| | | | |
|---|---|---|---|
| 290709 | Albanell Mestres, Joan | CEIC Parc de Salut Mar; IMIM- Hospital del Mar, C/ Dr. Aiguader, 88, Planta 1, 08003, Barcelona, BARCELONA, SPAIN | 08-Mar-2016 |
| 290710 | Redondo Sanchez, Andres | CEIC Parc de Salut Mar; IMIM- Hospital del Mar, C/ Dr. Aiguader, 88, Planta 1, 08003, Barcelona, BARCELONA, SPAIN | 08-Mar-2016 |
| 290711 | de la Haba Rodriguez, Juan | CEIC Parc de Salut Mar; IMIM- Hospital del Mar, C/ Dr. Aiguader, 88, Planta 1, 08003, Barcelona, BARCELONA, SPAIN | 08-Mar-2016 |
| 290713 | Saura Manich, Cristina | CEIC Parc de Salut Mar; IMIM- Hospital del Mar, C/ Dr. Aiguader, 88, Planta 1, 08003, Barcelona, BARCELONA, SPAIN | 08-Mar-2016 |
| 290714 | Stradella, Agostina | CEIC Parc de Salut Mar; IMIM- Hospital del Mar, C/ Dr. Aiguader, 88, Planta 1, 08003, Barcelona, BARCELONA, SPAIN | 08-Mar-2016 |
| 290715 | Jimenez, Begoña | CEIC Parc de Salut Mar; IMIM- Hospital del Mar, C/ Dr. Aiguader, 88, Planta 1, 08003, Barcelona, BARCELONA, SPAIN | 08-Mar-2016 |
| 290716 | Ruiz Simo, Amparo | CEIC Parc de Salut Mar; IMIM- Hospital del Mar, C/ Dr. Aiguader, 88, Planta 1, 08003, Barcelona, BARCELONA, SPAIN | 08-Mar-2016 |
| 290722 | Turner, Nicholas | NRES Committee London - City and East; Bristol Research Ethics Committee Centre | 26-May-2016 |
| 290723 | Board, Ruth | NRES Committee London - City and East; Bristol Research Ethics Committee Centre | 06-Jul-2016 |
| 290728 | Torregroza, Marco | Comite De Etica Medica E Investigacion | 24-Nov-2016 |
| 290798 | PETRUZELKA, LUBOS | Etická komise pro multicentrická klinická hodnocen, Na Bojisti 1, 128 08, Praha 2, CZECH REPUBLIC | 15-Dec-2016 |
| 291208 | Jiménez, Geiner | Comite Etico Cientifico Universidad de Ciencias Medicas, Sabana Norte, del ICE 200 metros noreste, edificio esquinero, primer piso, San José, COSTA RICA | 12-Sep-2016 |
| 291293 | Tredan, Olivier | CPP Sud Est IV, Centre Léon Bérard, 28 rue Laennec, 69373 Lyon, France | 05-Apr-2016 |
| 291294 | Mansi, Laura | CPP Sud Est IV, Centre Léon Bérard, 28 rue Laennec, 69373 Lyon, France | 05-Apr-2016 |
| 291296 | Jacot, William | CPP Sud Est IV, Centre Léon Bérard, 28 rue Laennec, 69373 Lyon, France | 05-Apr-2016 |
| 291297 | Loirat Legrain, Delphine | CPP Sud Est IV, Centre Léon Bérard, 28 rue Laennec, 69373 Lyon, France | 05-Apr-2016 |
| 291298 | Arnaud, Antoine | CPP Sud Est IV, Centre Léon Bérard, 28 rue Laennec, 69373 Lyon, France | 05-Apr-2016 |
| 291299 | MOUSSEAU, MIREILLE | CPP Sud Est IV, Centre Léon Bérard, 28 rue Laennec, 69373 Lyon, France | 05-Apr-2016 |
| 291303 | Frenel, Jean Sebastien | CPP Sud Est IV, Centre Léon Bérard, 28 rue Laennec, 69373 Lyon, France | 05-Apr-2016 |
| 291306 | Mailliez, Audrey | CPP Sud Est IV, Centre Léon Bérard, 28 rue Laennec, 69373 Lyon, France | 05-Apr-2016 |
| 291307 | PETIT, THIERRY | CPP Sud Est IV, Centre Léon Bérard, 28 rue Laennec, 69373 Lyon, France | 05-Apr-2016 |
| 291327 | Citron, Marc | QUORUM REVIEW IRB, 1501 Fourth Ave., Suite 800, Seattle, WA, 98101, UNITED STATES | 15-Feb-2016 |
| 291397 | SPANO, JEAN-PHILIPPE | CPP Sud Est IV, Centre Léon Bérard, 28 rue Laennec, 69373 Lyon, France | 05-Apr-2016 |
| 291398 | LORTHOLARY, ALAIN | CPP Sud Est IV, Centre Léon Bérard, 28 rue Laennec, 69373 Lyon, France | 05-Apr-2016 |
| 291405 | Nakajima, Hikaru | St. Luke's Hospital and Health Network IRB, 801 Ostrum Street, Bethlehem, PA, 18015, UNITED STATES | 01-Mar-2016 |
| 291406 | Ma, Cynthia | Washington University Medical Center, Human Studies Committee, 660 S. Euclid Avenue, Campus Box 8089, St. Louis, MO, 63110, UNITED STATES | 25-Aug-2016 |
| 291407 | Thumma, Saritha | Quorum, 1501 Fourth Avenue, Suite 800, SEATTLE, WA, 98101, UNITED STATES | 17-May-2016 |

| | | | |
|---|---|---|---|
| 291409 | Rao, Ruta | Rush University Medical Center; Rush University Medical Center Institutional Review Board, 1653 West Congress Parkway, Jelke Building Room 1591, Chicago, IL, 60612, UNITED STATES | 17-Jun-2016 |
| 291410 | Mashru, Sandeep | Kaiser Permanente Northwest Region IRB, 3800 N. Interstate Ave., Portland, OR, 97227, UNITED STATES | 20-Apr-2016 |
| 291412 | Wallmark, John | Quorum, 1501 Fourth Avenue, Suite 800, SEATTLE, WA, 98101, UNITED STATES | 13-Apr-2016 |
| 291427 | Beck, Thad | Quorum, 1501 Fourth Avenue, Suite 800, SEATTLE, WA, 98101, UNITED STATES | 10-Mar-2016 |
| 291460 | Turner, Nicholas | NRES Committee London - City and East; Bristol Research Ethics Committee Centre | 26-May-2016 |
| 291878 | Lee, Keun Seok | National Cancer Center Institutional Review Board, 323 Ilsan-ro, Ilsandong-gu, 410-769, Goyang-si, KOREA, REPUBLIC OF | 07-Apr-2016 |
| 291912 | BEITH, JANE | Sydney Local Health District Ethics Review Committee (RPAH Zone), 100 Carillon Avenue, Suite 210A, RPAH Medical Centre, 2042, Newtown, New South Wales, AUSTRALIA | 06-Sep-2016 |
| 291952 | Sanchez, Cesar | Comite de Etica de la Pontificia Uni Catolica de Chile, Marcoleta 391, 8330024, Santiago, CHILE | 18-Aug-2016 |
| 292167 | Graff, Stephanie | WIRB, 1019 39th Avenue SE, Puyallup, WA, 98374, UNITED STATES | 16-Mar-2016 |
| 292247 | Weckstein, Douglas | Quorum, 1501 Fourth Avenue, Suite 800, SEATTLE, WA, 98101, UNITED STATES | 31-Mar-2016 |
| 292358 | Emde, Till-Oliver | Ethik-Kommission der Ärztekammer Westfalen-Lippe und der Medizinischen Fakultät der WWU Münster, Gartenstr. 210 - 214, 48147, Münster, GERMANY | 04-May-2016 |
| 292359 | HAGEN, VOLKER | Ethik-Kommission der Ärztekammer Westfalen-Lippe und der Medizinischen Fakultät der WWU Münster, Gartenstr. 210 - 214, 48147, Münster, GERMANY | 04-May-2016 |
| 292361 | Lüdtke-Heckenkamp, Kerstin | Ethikkommission der Ärztekammer Niedersachsen, Berliner Allee 20, 30175, Hannover, GERMANY | 18-May-2016 |
| 292628 | Villarreal Garza, Cynthia Mayte | CEI del Instituto Tecnologico y de Estudios Superiores de Monterrey; ITESM, Av. Morones PrietoNo. 3000 Pte. Col. Los Doctores, 64710, MONTERREY, NUEVO LEON, MEXICO | 20-Jun-2016 |
| 293239 | Wilson, Caroline | NRES Committee London - City and East; Bristol Research Ethics Committee Centre | 15-Aug-2016 |
| 294075 | Okada, Morihito | Hiroshima University hospital Institutional Review Board, 1-2-3, Kasumi, Minami-ku, Hiroshima-shi, 734-8551, Hiroshima, JAPAN | 04-Jul-2016 |
| 294358 | Takahashi, Masato | National Hospital Organization Hokkaido Cancer Center Institutional Review Board, 2-3-54, Kikusui 4jyo, Shiroishi-ku, Sapporo-shi, 003-0804, Hokkaido, JAPAN | 14-Jul-2016 |
| 294370 | Ishida, Takanori | Tohoku University Hospital Institutional Review Board, 1-1 Seiryo-machi, Aoba-ku, Sendai-shi, 980-8574, Miyagi, JAPAN | 25-Jul-2016 |
| 294412 | SAJI, SHIGEHIRA | Fukushima Medical University Hospital Institutional Review Board, 1 Hikarigaoka, Fukushima-shi, 960-1295, Fukushima, JAPAN | 10-Aug-2016 |
| 294413 | Yanagita, Yasuhiro | Gunma Prefectural Cancer Center Institutional Review Board, 617-1 Takahayashinishi-cho, Ota-shi, 373-8550, Gunma, JAPAN | 26-Jul-2016 |
| 294414 | Inoue, Kenichi | Saitama Cancer Center Institutional Review Board, 780 Komuro Inamachi, Kitaadachi-gun, 362-0806, Saitama, JAPAN | 26-Jul-2016 |
| 294415 | SAEKI, TOSHIAKI | Saitama Medical University International Medical Center Institutional Review Board, 1397-1 Yamae, Hidaka-shi, 350-1298, Saitama, JAPAN | 27-Jul-2016 |
| 294416 | Tamura, Kenji | National Cancer Center Institutional Review Board, 5-1-1 Tsukiji Chuo-Ku, 104-0045, Tokyo, JAPAN | 27-Jul-2016 |

| | | | |
|---|---|---|---|
| 294418 | ITO, YOSHINORI | The Cancer Institute Hospital of JFCR Institutional Review Board, 3-8-31 Ariake Koto-Ku, 135-8550, Tokyo, JAPAN | 03-Aug-2016 |
| 294419 | Aruga, Tomoyuki | Tokyo Metropolitan Komagome Hospital Ethics Committee, 3-18-22 HONKOMAGOME, BUNKYO-KU, 113-8677, TOKYO, JAPAN | 13-Jul-2016 |
| 294424 | Niikura, Naoki | Tokai University Hospital Institutional Review Board, 143 Shimokasuya, Isehara-shi, 259-1193, Kanagawa, JAPAN | 28-Jul-2016 |
| 294426 | Tsugawa, Koichiro | IRB of a group of St. Marianna Univ. School of Medicine Hospitals, 2-16-1 Sugao,Miyamae-ku, Kawasaki-shi, 216-8511, Kanagawa, JAPAN | 22-Jul-2016 |
| 294427 | Kaneko, Koji | Niigata Cancer Center Hospital Institutional Review Board, 2-15-3 Kawagishi-cho, Chuo-ku, Niigata-shi, 951-8566, Niigata, JAPAN | 14-Jul-2016 |
| 294430 | WATANABE, JUNICHIRO | Shizuoka Cancer Center Ethical Review Board for Clinical Studies, 1007 Shimonagakubo; Nagaizumi-cho,Suntou-gun, 411-8777, Shizuoka, JAPAN | 07-Sep-2016 |
| 294431 | Sawaki, Masataka | Aichi Cancer Center Hospital Institutional Review Board, 1-1 Kanokoden, Chikusa-ku, Nakoya-shi, 464-8681, Aichi, JAPAN | 28-Jul-2016 |
| 294432 | Mizuno, Toshiro | Mie University Hospital Institutional Review Board, 2-174 Edobashi, Tsu-shi, 514-8507, Mie, JAPAN | 20-Jul-2016 |
| 294433 | Suzuki, Eiji | Kyoto University Hospital Institutional Review Board, 54 Kawaharacho, Shogoin, Sakyo-ku, 606-8507, Kyoto, JAPAN | 27-Jul-2016 |
| 294435 | Nakayama, Takahiro | Institutional Review Board of Osaka International Cancer Institute, 3-1-69, Otemae, Chuo-ku, Osaka-shi, 541-8567, Osaka, JAPAN | 21-Jul-2016 |
| 294446 | Iwasa, Tsutomu | Kindai University Hospital Institutional Review Board, 377-2 Ohnohigashi, Osaka-Sayama-shi, 589-8511, Osaka, JAPAN | 26-Jul-2016 |
| 294447 | MASUDA, NORIKAZU | National Hospital Organization Osaka National Hospital Institutional Review Board, 2-1-14 Hoenzaka, Chuo-ku, Osaka-shi, 540-0006, Osaka, JAPAN | 19-Jul-2016 |
| 294448 | Miyoshi, Yasuo | Hyogo College of Medicine Institutional Review Board, 1-1 Mukogawa-cho, Nishinomiya-shi, 663- 8501, Hyogo, JAPAN | 19-Jul-2016 |
| 294449 | Ohtani, Shoichiro | Hiroshima City Hospital Institutional Review Board, 7-33 Motomachi, Naka-ku, HIroshima-shi, 730-8518, Hiroshima, JAPAN | 27-Jul-2016 |
| 294451 | Sagara, Yasuaki | Sagara Hospital Institutional Review Board, 3-31 Matsubara-cho, Kagoshima-shi, 892-0833, Kagoshima, JAPAN | 09-Aug-2016 |
| 294452 | Kamada, Yoshihiko | Okinawa Medical Association Institutional Review Board, 218-9 Arakawa Haebaru-cho Shimajiri-gun, 901-1105, Okinawa, JAPAN | 29-Jun-2016 |
| 294456 | MARGOLIN, SARA | Regionala Etikprovningsnamnden i Uppsala, P.O. Box 1964, Drottninggatan 4, 751 49, Uppsala, SWEDEN | 08-Sep-2016 |
| 294470 | IWASE, HIROTAKA | The Institutional Review Board of Kumamoto University Hospital, 1-1-1 Honjo, Chuo-ku, Kumamoto-shi, 860-8556, Kumamoto, JAPAN | 27-Jul-2016 |
| 294471 | Yamauchi, Teruo | St. Luke's International Hospital Institutional Review Board, 9-1 Akashi-cho, Chuo-ku, 104-8560, Tokyo, JAPAN | 14-Jul-2016 |
| 294599 | Doihara, Hiroyoshi | IRB of Okayama University Hospital, 2-5-1 Shikata-cho, Kita-ku, Okayama-shi, 700−8558, Okayama, JAPAN | 16-Aug-2016 |
| 295937 | Waters, Simon | NRES Committee London - City and East; Bristol Research Ethics Committee Centre | 13-Dec-2016 |
| 296702 | Grafe, Andrea | Ethik-Kommission der Landesärztekammer Thüringen, Im Semmicht 33, 07751, Jena, GERMANY | 15-Nov-2016 |
| 297231 | Freitas Junior, Ruffo | Comitê de Ética em Pesquisa Associação de Combate ao Câncer em GO | 30-Jan-2017 |
| 298892 | LICHINITSER, MIKHAIL | Blokhin Russian Cancer Research Center Ethics Committee , Kashirskoye shosse,24, Moscow, RUSSIAN FEDERATION | 27-Dec-2016 |
