## Supplementary material for "Allelic Variation in *HLA-DRB1* is Associated with Development of Anti-Drug Antibodies in Cancer Patients Treated with Atezolizumab that are Neutralizing *in Vitro*": Ethics Committee approvals: IMpower130 EC IRB.pdf

| Country | EC/IRB | Address |
|---|---|---|
| Belgium | Cliniques Universitaires Saint-Luc - Comité | Promenade de l'Alma 51 bte B1.43.03 Bruxelles 1200 Belgium |
| Canada | Comite d'ethique de la recherche de l' Hopital Maisonneuve-Rosemont | 10 Rue De L'Espinay Quebec Quebec G1L 3L5 Canada |
| Canada | McGill University Health Center Montreal Hospital | 1032 W. Sheridan Road Research Services Granada Center, Suite 400 Chicago Illinois 60660 United States |
| Canada | Royal Victoria Regional Health Centre Research Ethics Board | Royal Perth Hospital, Wellington Street Perth Washington 6001 Australia |
| Canada | UBC BCCA Research Ethics Board | 750 West Broadway Fairmont Medical Building Suite 902 Vancouver British Columbia V5Z 1H5 Canada |
| Canada | William Osler Health system Research Ethics Board | 2100 Bovaird Drive East Brampton Ontario L6W 3J7 Canada |
| France | CPP Est III | 1020 Almira Street 3rd Floor, Room 321 Saginaw Michigan 48602 United States |
| Germany | Ethikkommission der Landesärztekammer Baden-Württemberg | Bachstrasse 18 Gebaeude 1 Jena 7740 Germany |
| Germany | Ethik-Kommission der Sächsischen Landesärztekammer | Gartenstrasse 210 - 214 Münster Nordrhein-Westfalen 48147 Germany |
| Hong Kong | Joint Chinese University of Hong Kong - New Territories East Cluster Clinical Research Ethics | 30 - 32 Ngan Shing Street, Prince of Wales Hospital 8th Floor, Lui Che Woo Clinical Science Building, Hong Kong Hong Kong |
| Israel | Assaf Harofe Medical Center EC | Beer Yaakov 70300 Zerifin 70300 Israel |
| Israel | Galilee Medical Center EC | Western Galilee Hospital POB 21 Nahariya 22100 Israel |
| Israel | Hadassah University Hospital Local EC | Wilgenstraat 2 Campus Wilgenstraat Roeselare West-Vlaanderen 8800 Belgium |
| Israel | Kaplan Medical Center Local EC | Kaplan Medical Center P.o Box 1 Rehovot 76100 Israel |
| Israel | Meir EC | National Institute Of Health, C/o Institute For Health Management, Kuala Lumpur Bangsar WilayahPersekutuan KualaLumpur 59000 Malaysia |
| Israel | Rabin Medical Center Ethics Committee | 1501 Fourth Avenue Suite 800 Seattle Washington 98101 United States |
| Israel | Rabin Medical Center Local EC | 39 Jabutinsky street Petah Tikva 49100 Israel |
| Israel | Rambam Medical Center Ethics Committee | 8 Haaliya Hashniya Street Bat Galim Haifa 31096 Israel |
| Israel | Soroka University Medical Center Local EC | Post Office Box 151 Beer Sheva 84101 Israel |
| Israel | Tel Aviv Sourasky EC | 6 Weitzman Street Tel Aviv Tel-Aviv 64239 Israel |
| Israel | The Chaim Sheba Medical Center EC | Chaim Sheba Medical Center Tel Hashomer Ramat Gan 52621 Israel |
| Italy | Comitato Etico Campania Nord | Via Giovanbattista Pergolesi 33 Monza Lombardia 20900 Italy |
| Italy | Comitato Etico delle Aziende Sanitarie dell'Umbria | Largo Francesco Vito, 1 Roma Lazio 168 Italy |

| | | |
|---|---|---|
| **Italy** | **Comitato Etico delle Province di Chieti e Pescara** | Largo Francesco Vito, 1 Roma Lazio 168 Italy |
| **Italy** | **Comitato Etico IRCCS Istituto Nazionale per lo Studio e la Cura dei Tumori Fondazione G Pascale** | Via Dell'eremo 9/11 Lecco Lombardia 23900 Italy |
| **Italy** | **Comitato Etico Regionale delle Marche** | Via Olgettina, 60 Milano Lombardia 20132 Italy |
| **Italy** | **Comitato Etico Seconda Università degli Studi di Napoli Az. Osp. Univ. S.U.N. - A.O.R.N. "Ospedali** | Via Olgettina, 60 Milano Lombardia 20132 Italy |
| **Spain** | **CEIC Consorcio Hospital General Universitario de Valencia** | Avenida Tres Cruces, 2 Consorcio Hospital General Universitario de Valencia Pabellon B-3 - 4ª planta Valencia Valencia 46014 Spain |
| **Spain** | **CEIC de Aragon (CEICA)** | Calle Micer Masco, 31 Valencia Valencia 46010 Spain |
| **Spain** | **CEIC de Galicia (CAEI)** | Calle Micer Masco, 31 Valencia Valencia 46010 Spain |
| **Spain** | **CEIC Hospital de la Santa Creu i Sant Pau** | Avenida Vicente Blasco Ibánez, 17 Valencia Valencia 46010 Spain |
| **Spain** | **CEIC Hospital Santa Creu i Sant Pau** | Avenida Sant Antoni Maria Claret, 167 Servicio de Famacología Clínica Pabellón HC Barcelona Barcelona 8025 Spain |
| **Spain** | **CEIC Hospital Universitario de Canarias** | Avda. de Córdoba s/n Instituto de Investigación Hospital 12 de Octubre (i+12) Bloque D - Planta 6ª Area de Gestión de Proyectos - Unidad Administrativa CEIC Madrid 28041 Spain |
| **United States** | **Appalachian Regional Healthcare IRB** | 58 Canal Circular Road Apollo Gleneagles Hospitals Kolkata Kolkata West Bengal 700054 India |
| **United States** | **Banner MD Anderson Cancer Center IRB** | 2940 E. Banner Gateway Dr. Suite 375 Gilbert California 85234 United States |
| **United States** | **Biomedical Research Alliance of New York LLC Institutional Review Board** | University Of North Carolina, Cb # 7097 Medical School Building 52 Chapel Hill North Carolina 27599 United States |
| **United States** | **Birmingham Veterans Administration Medical Center IRB** | 207 Olds Hall East Lansing Michigan 48824 United States |
| **United States** | **Copernicus Group Independent Review Board** | 5000 CentreGreen Way Suite 200 Cary North Carolina 27513 United States |
| **United States** | **Copernicus IRB** | 1 Triangle Drive Suite 100 Po Box 110605 Research Triangle Park North Carolina 27709 United States |
| **United States** | **Duke University Health System Institutional Review Board** | Hock Plaza Durham North Carolina 27705 United States |
| **United States** | **Englewood Hospital and Medical Center** | 350 Engle Street Englewood New Jersey 7631 United States |
| **United States** | **Kaiser Permanente Northern California Institutional Review Board** | 1800 Harrison Street 16th Floor Oakland California 94162 United States |

| | | |
|---|---|---|
| **United States** | **Kaiser Permanente of Colorado Institutional Review Board** | 3800 North Interstate Avenue Portland Oregon 97227 United States |
| **United States** | **Lahey Clinic, Inc. Institutional Review Board** | 3-23-1, Shiobara, Minami-ku Fukuoka-shi 815-8588 Japan |
| **United States** | **Lancaster General Hospital IRB** | 3-23-1, Shiobara, Minami-ku Fukuoka-shi 815-8588 Japan |
| **United States** | **Loyola University Institutional Review Board** | 1032 W. Sheridan Road Research Services Granada Center, Suite 400 Chicago Illinois 60660 United States |
| **United States** | **Mayo Clinic Institutional Review Board** | 1032 W. Sheridan Road Research Services Granada Center, Suite 400 Chicago Illinois 60660 United States |
| **United States** | **MD Anderson Institutional Review Board** | 1400 Pressler Street Unit 1452 Houston Texas 77030-4009 United States |
| **United States** | **New England Institutional Review Board** | 85 Wells Avenue Suite 107 Newton Massachussets 2459 United States |
| **United States** | **North Mississippi Health Services** | 830 South Gloster Street Tupelo Mississippi 38801 United States |
| **United States** | **NYU School of Medicine Institutional Review Board** | 68, Hangeulbiseok-ro,Nowon-gu Seoul 1830 Korea, Republic of |
| **United States** | **Ochsner Clinic Foundation Institutional Review** | 68, Hangeulbiseok-ro,Nowon-gu Seoul 1830 Korea, Republic of |
| **United States** | **Pinnacle Health Hospitals Institutional Review** | 1968 Peachtree Road Northwest Atlanta Georgia 30309 United States |
| **United States** | **Rhode Island Hospital Institutional Review Board** | 1 Hoppin Street Office of Research Administration, Coral West Suite 1300 Committee on the Protection of Human Subjects Providence Rhode Island 2903 United States |
| **United States** | **Saint Barnabas Medical Center Institutional Review Board** | 1055 North Curtis Road Boise Idaho 83706 United States |
| **United States** | **Siouxland Institutional Review Board** | 230 Nebraska Street Sioux City Iowa 51101 United States |
| **United States** | **Springfield Committee for Research Involving Human Subjects (SCRIHS)** | 801 North Rutledge Street Springfield Illinois 62702 United States |
| **United States** | **The Christ Hospital IRB** | 3535 Market Street Suite 1200 Philadelphia Pennsylvania 19104 United States |
| **United States** | **University of Arkansas IRB** | Lembah Pantai Kuala Lumpur 59100 Malaysia |
| **United States** | **University of Chicago Hospitals Institutional Review Board** | 5751 South Woodlawn Avenue McGiffert Hall Chicago Illinois 60637 United States |
| **United States** | **University Of Iowa Human Subjects Office IRB** | 1 Illini Drive Peoria Illinois 61656 United States |
| **United States** | **University of Louisville IRB** | 501 East Broadway Medcenter One Suite 200 Louisville Kentucky 40202 United States |
| **United States** | **University Of Miami** | 55 Lake Ave., North Worcester Massachussets 1655 United States |
| **United States** | **University of Nevada Reno Research Integrity Office** | 987830 Nebraska Medical Center Omaha Nebraska 68198-7830 United States |
| **United States** | **W.G. 'Bill' Hefner VA Medical Center** | 1601 Brenner Avenue Salisbury North Carolina 28144 United States |

| | | |
|---|---|---|
| **United States** | **Walter Reed National Military Medical Center IRB** | 1601 Brenner Avenue Salisbury North Carolina 28144 United States |
| **United States** | **Western Institutional Review Board** | 1019 39th Avenue Southeast Suite 120 Puyallup Washington 98374 United States |
| **United States** | **Western Institutional Review Board (WIRB)** | 1019 39th Avenue Southeast Suite 120 Puyallup Washington 98374 United States |
| **United States** | **WIRB Copernicus Group** | 222 Station Plaza North Suite 521 Mineola New York 11501 United States |
