## Supplementary material for "Allelic Variation in *HLA-DRB1* is Associated with Development of Anti-Drug Antibodies in Cancer Patients Treated with Atezolizumab that are Neutralizing *in Vitro*": Ethics Committee approvals: IMpower131 EC IRB.pdf

| Site # | Country | Investigator | EC/IRB Name and Address | IRB/EC Approval |
|---|---|---|---|---|
| 279803 | Argentina | Jarchum, Gustavo | Comité Institutcional de Etica de Investigación en Salud Sanatorio Allende, Hipolito Yrigoyen 384, X5000JHQ, Córdoba, Córdoba, Argentina | 11/12/15 |
| 279804 | Argentina | Magri, Ignacio | Comité Institucional de Ética de Investigación en Salud del Instituto Médico Río Cuarto, Hipólito Yrigoyen 1020, 5800, Rio Cuarto, Córdoba, Argentina | 10/6/15 |
| 282502 | Argentina | Pastor, Andrea | Comite De Etica Del Hospital Provincial Del Centenario, Urquiza 3101, S2002KDS, Rosario, Santa Fe, Argentina | 11/25/15 |
| 279805 | Argentina | Kotliar, Mauricio | Comité de Ética Independiente - Fundación Sanatorio, Francisco Acuña de Figueroa 1240, C1180AAX, Buenos Aires, Ciudad Autónoma de BuenosAires, Argentina | 4/28/15 |
| 279801 | Argentina | Cigno, Edgardo | Comite De Etica Del Sanatorio Britanico Sa, Paraguay 40, S2000CVB, Rosario, Santa Fe, Argentina | 1/26/16 |
| 279800 | Argentina | Martin, Claudio | Comité de Etica en Investigación del Instituto Alexander Fleming, Cramer 1180, C1426ANZ, Buenos Aires, Ciudad Autónoma de BuenosAires, Argentina | 9/16/15 |
| 279799 | Argentina | Lerzo, Guillermo | Comité Independiente de Etica en investigación clínica "Dr. Carlos A. Barclay, Larrea 1381, 3° A, 1117, Buenos Aires, Ciudad Autónoma de BuenosAires, Argentina | 2/12/15 |
| 279798 | Argentina | Kowalyszyn, Rubén | Comité Independiente de Etica en investigación clínica "Dr. Carlos A. Barclay, Larrea 1381, 3° A, 1117, Buenos Aires, Ciudad Autónoma de BuenosAires, Argentina | 3/9/15 |
| 279796 | Argentina | Kahl, Susana | Comité de ética en investigación Fundación Oncosalud, Siria 16, 2700, Perga, Buenos Aires, Argentina | 4/16/15 |
| 279795 | Argentina | Varela, Mirta | Comité de Ética Centro de Oncología e Investigación Buenos Aires, Calle 12 #4756, B1880BBF, Berazategui, Buenos Aires, Argentina | 2/1/16 |
| 279794 | Argentina | Kaen, Diego | Comité Independiente de Etica en investigación clínica "Dr. Carlos A. Barclay, Larrea 1381, 3° A, 1117, Buenos Aires, Ciudad Autónoma de BuenosAires, Argentina | 2/19/15 |
| 279793 | Argentina | Picon, Pablo | Comité de Ética Independiente Patagónico, San Martin 391, L6300DVM, Santa Rosa, La Pampa, Argentina | 4/9/15 |
| 278899 | Australia | Singhal, Nimit | Hunter New England Research Ethics and Governance Unit, Locked Bag 1, 2305, New Lambton, New South Wales, Australia | 3/26/15 |
| 278900 | Australia | Gauden, Stan | Tasmania Health and Medical Human Research Ethics Committee, 301 Sandy Bay Road, 7001, Hobart, Tasmania, Australia | 3/30/15 |

| | | | | |
|---|---|---|---|---|
| 278905 | Australia | Crombie, Catherine | Hunter New England Research Ethics and Governance Unit, Locked Bag 1, 2305, New Lambton, New South Wales, Australia | 3/26/15 |
| 278907 | Australia | Blinman, Prunella | Hunter New England Research Ethics and Governance Unit, Locked Bag 1, 2305, New Lambton, New South Wales, Australia | 3/26/15 |
| 278908 | Australia | Potasz, Nicole | Hunter New England Research Ethics and Governance Unit, Locked Bag 1, 2305, New Lambton, New South Wales, Australia | 3/26/15 |
| 280017 | Australia | Lewis, Craig | Hunter New England Research Ethics and Governance Unit, Locked Bag 1, 2305, New Lambton, New South Wales, Australia | 3/26/15 |
| 280019 | Australia | Gill, Sanjeev | Hunter New England Research Ethics and Governance Unit, Locked Bag 1, 2305, New Lambton, New South Wales, Australia | 3/26/15 |
| 281140 | Australia | Eliadis, Paul | Bellberry Human Research Ethics Committee, 123 Glen Osmond Road, 5063, Eastwood, South Australia, Australia | 4/24/15 |
| 280018 | Australia | Richardson, Gary | Cabrini Human Research Ethics Committee, 183 Wattletree Road, 3144, Malvern, Vic, Australia | 7/29/15 |
| 280016 | Australia | Kosmider, Suzanne | Hunter New England Research Ethics and Governance Unit, Locked Bag 1, 2305, New Lambton, New South Wales, Australia | 3/26/15 |
| 278906 | Australia | Millward, Michael | Sir Charles Gairdner Hospital HREC, Hospital Avenue, 6009, Nedlands, Western Australia, Australia | 7/28/15 |
| 278904 | Australia | John, Thomas | Hunter New England Research Ethics and Governance Unit, Locked Bag 1, 2305, New Lambton, New South Wales, Australia | 12/15/15 |
| 278903 | Australia | Joshi, Abhishek | Hunter New England Research Ethics and Governance Unit, Locked Bag 1, 2305, New Lambton, New South Wales, Australia | 3/26/15 |
| 278902 | Australia | Nordman, Ina | Hunter New England Research Ethics and Governance Unit, Locked Bag 1, 2305, New Lambton, New South Wales, Australia | 7/7/15 |
| 278901 | Australia | Boyer, Michael | Sydney Local Health District Human Research Ethics Committee - CRGH, Hospital Road, 2139, Concord, New South Wales, Australia | 6/18/15 |
| 278898 | Australia | O’Byrne, Kenneth | Hunter New England Research Ethics and Governance Unit, Locked Bag 1, 2305, New Lambton, New South Wales, Australia | 3/26/15 |
| 278897 | Australia | Hughes, Brett | Hunter New England Research Ethics and Governance Unit, Locked Bag 1, 2305, New Lambton, New South Wales, Australia | 3/26/15 |

| | | | | |
|---|---|---|---|---|
| 280014 | Austria | Greil, Richard | Ethikkommission für das Bundesland Salzburg, Sebastian-Stief-Gasse 2, 5010, Salzburg, Salzburg, Austria | 10/21/15 |
| 279880 | Austria | Eckmayr, Josef | Ethikkommission des Landes Oberösterreich, Wagner-Jauregg Weg 15, 4020, Linz, , Austria | 10/21/15 |
| 280642 | Belgium | Dirix, Luc | AZ Sint Augustinus, Oosterveldlaan 24, 2610, Wilrijk, Antwerpen, Belgium | 6/16/15 |
| 279755 | Belgium | Collard, Philippe | Comité d’Ethique hospitalo-facultaire Cliniques universitaires Saint-Luc, Promenade de l’Alma 51 bte B1.43.03, 1200, Bruxelles, Brussels, Belgium | 6/16/15 |
| 280979 | Belgium | Jerusalem, Guy | Comité d’Ethique hospitalo-facultaire Cliniques universitaires Saint-Luc, Promenade de l’Alma 51 bte B1.43.03, 1200, Bruxelles, Brussels, Belgium | 6/16/15 |
| 280980 | Belgium | Goeminne, Jean Charles | Comité d’Ethique hospitalo-facultaire Cliniques universitaires Saint-Luc, Promenade de l’Alma 51 bte B1.43.03, 1200, Bruxelles, Brussels, Belgium | 6/16/15 |
| 279754 | Belgium | Alexander, Patrick | Ethisch Comité Werken Glorieux VZW, Glorieuxlaan 55, 9600, Ronse, Oost-Vlaanderen, Belgium | 6/16/15 |
| 283381 | Brazil | Schwartsmann, Gilberto | Comitê de Ética em Pesquisa do Hospital de Clínicas de Porto Alegre, Rua Ramiro Barcelos 2350, 90035-903, Porto Alegre, Rio Grande do Sul, , Brazil | 4/6/16 |
| 283380 | Brazil | Costamilan, Rita de Cassia | Comitê de Ética em Pesquisa da Universidade de Caxias do Sul, Rua Francisco Getúlio Vargas 1130, 95070-560, Caxias do Sul, Rio Grande do Sul, Brazil | 3/31/16 |
| 281359 | Brazil | Brust, Leandro | Comite de Etica em Pesquisa em Seres Humanos do Centro Universitario UNIVATES/RS, Rua Avelino Tallini 171, Bairro Universitário, 95900-000, Lajeado, Rio Grande do Sul, Brazil | 3/1/16 |
| 281357 | Brazil | Abbade Dettino, Aldo | Comitê de Ética em Pesquisa da Fundação Antônio Prudente - AC Camargo Câncer Center, Rua Professor Antonio Prudente 211, 01509-900, São Paulo, São Paulo, Brazil | 6/6/16 |
| 281356 | Brazil | Bruno, Luiz | Comitê de Ética em Pesquisa - Hospital Mãe de Deus, R. José de Alencar 286, 90880-480, Porto Alegre, Rio Grande do Sul, Brazil | 5/6/16 |
| 280820 | Brazil | Campos, Clodoaldo | Comitê de Ética em Pesquisa em Seres Humanos da Irmandade da Santa Casa de Londrina, Rua Espírito Santo, 523, 86010-510, Londrina, Paraná, Brazil | 5/5/16 |
| 280819 | Brazil | Matias, Danielli | Comitê de Ética em Pesquisa em Seres Humanos da Liga Norte Riograndense Contra o Câncer, Rua Dr. Mário Negócio, 2267, 59040-000, Natal, Rio Grande do Norte, Brazil | 2/2/16 |
| 280816 | Brazil | Faccio, Adilson | Comitê de Ética em Pesquisa em Seres Humanos da Universidade de Ribeirão Preto (UNAERP), Avenida Costabile Romano, 2201, Ribeirania, 14096380, Ribeirão Preto, São Paulo, Brazil | 4/13/16 |

| | | | | |
|---|---|---|---|---|
| 280818 | Brazil | Murad, Andre | Comitê de Ética em Pesquisa em Seres Humanos do Hospital Lifecenter, Avenida Do Contorno, 4747, 30110-921, Belo Horizonte, Minas Gerais, Brazil | 2/19/16 |
| 280817 | Brazil | da Silva, Carlos | Comitê de Ética em Pesquisa Fundação Pio XII Hospital de Câncer de Barretos, Rua Antenor Duarte Villela 1331, 14784-400, Barretos, São Paulo, Brazil | 2/18/16 |
| 281353 | Brazil | Tadokoro, Hakaru | Comite de Etica em Pesquisa da Universidade Federal de Sao Paulo - Hospital Sao Paulo, Rua Botucatu 572, 04023-062, São Paulo, , Brazil | 3/17/16 |
| 280815 | Brazil | Aragao, Bruno | Comitê de Ética em Pesquisa em Seres Humanos do Hospital Socor, Rua Tupis, 1540, 30190-062, Belo Horizonte, Minas Gerais, Brazil | 3/9/16 |
| 280142 | Brazil | Girotto, Gustavo | Comitê de Ética em Pesquisa em Seres Humanos da Faculdade de Medicina de São José do Rio Preto, Avenida Brigadeiro Faria Lima 5416, Vila São Pedro, 15090-000, São José Do Rio Preto, , Brazil | 5/26/15 |
| 281203 | Bulgaria | Ilieva, Rumyana | Ethics Committee for Clinical Trials, 5 Sveta Nedelya Square, 1000, Sofia, Sofia-Grad, Bulgaria | 6/17/15 |
| 281201 | Bulgaria | Mihaylova, Zhasmina | Ethics Committee for Clinical Trials, 5 Sveta Nedelya Square, 1000, Sofia, Sofia-Grad, Bulgaria | 1/27/16 |
| 281200 | Bulgaria | Koynov, Krassimir | Ethics Committee for Clinical Trials, 5 Sveta Nedelya Square, 1000, Sofia, Sofia-Grad, Bulgaria | 6/17/15 |
| 281199 | Bulgaria | Dimitrov, Borislav | Ethics Committee for Clinical Trials, 5 Sveta Nedelya Square, 1000, Sofia, Sofia-Grad, Bulgaria | 6/17/15 |
| 288023 | Canada | Comeau, Reginald | Comite d'ethique de la recherche de l' Hopital Maisonneuve-Rosemont, 5415 Boulevard De L'assomption, H1T 2M4, Montreal, Quebec, Canada | 3/17/16 |
| 281190 | Canada | Conter, Henry | William Osler Health system Research Ethics Board, 2100 Bovaird Drive East, L6W 3J7, Brampton, Ontario, Canada | 5/14/15 |
| 281188 | Canada | Whittom, Renaud | Comite d'ethique de la recherche de l' Hopital Maisonneuve-Rosemont, 5415 Boulevard De L'assomption, H1T 2M4, Montreal, Quebec, Canada | 4/7/16 |
| 281186 | Canada | El-Maraghi, Robert | Royal Victoria Regional Health Centre Research Ethics Board, 201 Georgian Drive, L4M 6M2, Barrie, Ontario, Canada | 7/7/15 |
| 281185 | Canada | Dionne, Jean-Luc | Comite d'ethique de la recherche de l' Hopital Maisonneuve-Rosemont, 5415 Boulevard De L'assomption, H1T 2M4, Montreal, Quebec, Canada | 12/21/15 |
| 280021 | Canada | Rothenstein, Jeffrey | Lakeridge Health REB, 1 Hospital Court, L1G 2B9, Oshawa, Ontario, Canada | 7/15/15 |

| | | | | |
|---|---|---|---|---|
| 280053 | Canada | Cournoyer, Ghislain | Centre de Sante et de Services Sociaux de Saint-Jerome (CSSS) - Comite d'ethique de recherche, 290 Rue de Montigny, J7Z 5T3, Saint Jerome, Quebec, Canada | 6/7/16 |
| 291880 | Chile | Yañez Ruiz, Eduardo | Comité Ético-Científico del Servicio de Salud Metropolitano Oriente (CEC-SSMO), Av. Salvador 364, , Santiago, Región-MetropolitanadeSantiago, Chile | 3/29/16 |
| 288021 | Chile | Orellana Ulunque, Eric | Comité de Ética Científica de Clínica Santa María, Avenida Bellavista 0373, 7520379, Santiago, Región-MetropolitanadeSantiago, Chile | 11/9/15 |
| 279942 | Chile | Orlandi Jorquera, Francisco | Comité Ético Científico del Servicio de Salud Metropolitano Oriente (CEC SSMO), Av. Salvador 364, , Santiago, Región-MetropolitanadeSantiago, Chile | 5/26/15 |
| 279941 | Chile | Ahumada Olea, Mónica | Comité Ético Científico y de Investigación Hospital Clinico Universidad de Chile, Santos Dummont 999, 8380456, Santiago, , Chile | 8/5/15 |
| 281316 | France | Gazaille, Virgile | CPP Sud-Est II, Groupement Hospitalier Est- 59 Boulevard Pinel, 69500, Bron, , France | 6/10/15 |
| 281237 | France | Perol, Maurice | CPP Sud-Est II, Groupement Hospitalier Est- 59 Boulevard Pinel, 69500, Bron, , France | 6/10/15 |
| 281235 | France | Huchot, Eric | CPP Sud-Est II, Groupement Hospitalier Est- 59 Boulevard Pinel, 69500, Bron, , France | 6/10/15 |
| 281233 | France | Dixmier, Adrien | CPP Sud-Est II, Groupement Hospitalier Est- 59 Boulevard Pinel, 69500, Bron, , France | 6/10/15 |
| 281232 | France | Corre, Romain | CPP Sud-Est II, Groupement Hospitalier Est- 59 Boulevard Pinel, 69500, Bron, , France | 6/10/15 |
| 281231 | France | Denis, Fabrice | CPP Sud-Est II, Groupement Hospitalier Est- 59 Boulevard Pinel, 69500, Bron, , France | 6/10/15 |
| 281229 | France | Berard, Henri | CPP Sud-Est II, Groupement Hospitalier Est- 59 Boulevard Pinel, 69500, Bron, , France | 6/10/15 |
| 281227 | France | Becht-Rolly, Catherine | CPP Sud-Est II, Groupement Hospitalier Est- 59 Boulevard Pinel, 69500, Bron, , France | 6/10/15 |
| 279929 | France | Moro-Sibilot, Denis | CPP Sud-Est II, Groupement Hospitalier Est- 59 Boulevard Pinel, 69500, Bron, , France | 6/10/15 |
| 279928 | France | Souquet, Pierre Jean | CPP Sud-Est II, Groupement Hospitalier Est- 59 Boulevard Pinel, 69500, Bron, , France | 6/10/15 |

| | | | | |
|---|---|---|---|---|
| 281230 | France | Coudert, Bruno | CPP Sud-Est II, Groupement Hospitalier Est- 59 Boulevard Pinel, 69500, Bron, , France | 6/10/15 |
| 281236 | France | Dayen, Charles | CPP Sud-Est II, Groupement Hospitalier Est- 59 Boulevard Pinel, 69500, Bron, , France | 6/10/15 |
| 291416 | Germany | Frost, Nikolaj | Landesamt für Gesundheit und Soziales Berlin (LaGeSo) Ethik, Fehrbelliner Platz 1, 10707, Berlin, Berlin, Germany | 5/12/16 |
| 281317 | Germany | Dickgreber, Nicolas | Ethik-Kommission der Ärztekammer Westfalen-Lippe und der Medizinischen Fakultät der WWU Munster, Gartenstrasse 210 - 214, 48147, Münster, Nordrhein-Westfalen, Germany | 8/12/15 |
| 280822 | Germany | Reck, Martin | Ethik-Kommissionen bei der Ärztekammer Schleswig-Holstein, Bismarckallee 8-12, 23795, Bad Segeberg, Schleswig-Holstein, Germany | 8/12/15 |
| 280503 | Germany | Engel-Riedel, Walburga | Ethikkommission der Ärztekammer Nordrhein, Tersteegenstrasse 9, 40474, Düsseldorf, Nordrhein-Westfalen, Germany | 8/12/15 |
| 280502 | Germany | Weißinger, Florian | Ethik-Kommission der Ärztekammer Westfalen-Lippe und der Medizinischen Fakultät der WWU Munster, Gartenstrasse 210 - 214, 48147, Münster, Nordrhein-Westfalen, Germany | 8/12/15 |
| 280501 | Germany | Stauder, Heribert | Ethikkommission der Bayerischen Landesärztekammer, Mühlbaurstrasse 16, 81677, München, Bayern, Germany | 8/12/15 |
| 280500 | Germany | Schütte, Wolfgang | Ethik-Kommission der Ärztekammer Sachsen-Anhalt, Am Kirchtor 9, 6108, Halle an der Saale, , Germany | 8/12/15 |
| 279973 | Germany | Wehler, Thomas | Ethik-Kommission bei der Ärztekammer des Saarlandes, Faktoreistrasse 4, 66111, Saarbrücken, Saarland, Germany | 8/12/15 |
| 279972 | Germany | Loges, Sonja | Ethik-Kommission der Ärztekammer Hamburg, Weidestr. 122 b, 22083, Hamburg, Hamburg, Germany | 8/12/15 |
| 279970 | Germany | Lehmann, Markus | Ethik-Kommission der Ärztekammer Westfalen-Lippe und der Medizinischen Fakultät der WWU Munster, Gartenstrasse 210 - 214, 48147, Münster, Nordrhein-Westfalen, Germany | 8/12/15 |
| 279969 | Germany | Kokowski, Konrad | Ethik-Kommission der Bayerischen Landesärztekammer, Mühlbaurstr.16, 81677, München, , Germany | 8/12/15 |
| 279960 | Germany | Schulz, Christian | Ethikkommission an der Universität Regensburg, Landshuter Straße 4, 93047, Regensburg, , Germany | 8/12/15 |
| 279956 | Germany | Sadjadian, Parvis | Ethik-Kommission der Ärztekammer Westfalen-Lippe und der Medizinischen Fakultät der WWU Munster, Gartenstrasse 210 - 214, 48147, Münster, Nordrhein-Westfalen, Germany | 8/12/15 |

| | | | | |
|---|---|---|---|---|
| 279954 | Germany | Rittmeyer, Achim | Ethikkommission der Landesärztekammer Hessen, Im Vogelsgesang 3, 60488, Frankfurt am Main, Hessen, Germany | 8/12/15 |
| 279951 | Germany | Kimmich, Martin | Ethikkommission der Landesärztekammer Baden-Württemberg, Jahnstrasse 40, 70597, Stuttgart, Baden-Württemberg, Germany | 8/12/15 |
| 279950 | Germany | Heinrich, Bernhard | Ethikkommission der Bayerischen Landesärztekammer, Mühlbaurstrasse 16, 81677, München, Bayern, Germany | 8/12/15 |
| 279948 | Germany | Rückert, Anja | Ethikkommission der Landesärztekammer Baden-Württemberg, Jahnstrasse 40, 70597, Stuttgart, Baden-Württemberg, Germany | 8/12/15 |
| 279947 | Germany | Fischer, Jürgen | Ethikkommission der Landesärztekammer Baden-Württemberg, Jahnstrasse 40, 70597, Stuttgart, Baden-Württemberg, Germany | 8/12/15 |
| 279946 | Germany | Behringer, Dirk | Ethik-Kommission der Ärztekammer Westfalen-Lippe und der Medizinischen Fakultät der WWU Munster, Gartenstrasse 210 - 214, 48147, Münster, Nordrhein-Westfalen, Germany | 8/12/15 |
| 279945 | Germany | Bargon, Joachim | Ethikkommission der Landesärztekammer Hessen, Im Vogelsgesang 3, 60488, Frankfurt am Main, Hessen, Germany | 8/12/15 |
| 279944 | Germany | Wesseler, Claas | Ethik-Kommission der Ärztekammer Hamburg, Weidestr. 122 b, 22083, Hamburg, Hamburg, Germany | 8/12/15 |
| 279974 | Germany | Wermke, Martin | Ethik-Kommission der Medizinischen Fakultät "Carl Gustav Carus" der Technischen Universität Dresden, Fetscherstraße 74, 01307, Dresden, , Germany | 8/12/15 |
| 282806 | Germany | Wiewrodt, Rainer | Ethik-Kommission der Ärztekammer Westfalen-Lippe und der Medizinischen Fakultät der WWU Münster, Gartenstrasse 210 - 214, 48147, Münster, Nordrhein-Westfalen, Germany | 8/31/15 |
| 297763 | Germany | Laack, Eckart | Ethik-Kommission der Ärztekammer Westfalen-Lippe und der Medizinischen Fakultät der WWU Münster, Gartenstrasse 210 - 214, 48147, Münster, Nordrhein-Westfalen, Germany | 2/16/17 |
| 281141 | Israel | Kolin, Maya | Galilee Medical Center EC, Western Galilee Hospital POB 21, 22100, Nahariya, , Israel | 5/21/15 |
| 279781 | Israel | Wollner, Mirjana | Rambam Medical Center Ethics Committee, 8 Haaliya Hashniya Street, 31096, Haifa, Haifa, Israel | 7/26/15 |
| 279779 | Israel | Merimsky, Ofer | Tel Aviv Sourasky EC, 6 Weitzman Street, 6423906, TEL AVIV, , Israel | 5/28/15 |
| 279778 | Israel | Bar, Yair | The Chaim Sheba Medical Center EC, Tel Hashomer, 52621, Ramat Gan, , Israel | 4/28/15 |

| | | | | |
|---|---|---|---|---|
| 279777 | Israel | Cyjon, Arnold | Shamir Medical Center Assaf Harofeh EC, Beer Yaakov 70300, 70300, Zerifin, , Israel | 4/16/15 |
| 279775 | Israel | Dudnik, Julia | Soroka University Medical Center Local EC, Reger Avenu, 84101, Beer Sheva, , Israel | 3/5/15 |
| 279774 | Israel | Stemmer, Salomon | Rabin Medical Center Ethics Committee, 39 Jabotinski St., 49100, Petach Tikva, , Israel | 3/29/15 |
| 279773 | Israel | Nechushtan, Hovav | Hadassah University Hospital Local EC, Kiryat Hadassah, 91120, Jerusalem, , Israel | 4/1/15 |
| 279772 | Israel | Katsnelson, Rivka | Kaplan Medical Center Local EC, Kaplan Medical Center, 76100, Rehovot, , Israel | 5/19/15 |
| 279770 | Israel | Gottfried, Maya | Meir EC, 59 Tchernichovsky Street, 44281, Kfar Saba, HaMerkaz, Israel | 6/11/15 |
| 279863 | Italy | Allegrini, Giacomo | Comitato Etico Regionale Toscana – Area Vasta Nord Ovest, Via Roma 67, 56126, Pisa, Toscana, Italy | 7/23/15 |
| 279869 | Italy | Falcone, Alfredo | Comitato Etico Regionale Toscana – Area Vasta Nord Ovest, Via Roma 67, 56126, Pisa, Toscana, Italy | 7/23/15 |
| 279871 | Italy | Gridelli, Cesare | Comitato Etico Campania Nord, VIA DEGLI IMBIMBO 10-12, 83100, Avellino, Camparia, Italy | 6/23/15 |
| 279873 | Italy | Mencoboni, Manlio | Comitato Etico San Martino IST 2, Largo Rosanna Benzi 10, 16132, Genova, , Italy | 9/15/15 |
| 293924 | Italy | Pedrazzoli, Paolo | Comitato di Bioetica Fondazione IRCCS Policlinico S. Matteo di Pavia, Viale Golgi 19, 27100, Pavia, , Italy | 10/10/16 |
| 279875 | Italy | Morabito, Alessandro | Comitato Etico IRCCS<br>Istituto Nazionale per lo Studio e la Cura dei Tumori Fondazione G Pascale, Via Mariano Semmola, 80131, Napoli, Campania, Italy | 8/3/15 |
| 280015 | Italy | Soto-Parra, Hector | Comitato Etico Catania 1, VIA SANTA SOFIA 78, 95123, Catania, Sicilia, Italy | 6/9/15 |
| 281142 | Italy | Minotti, Vincenzo | Comitato Etico delle Aziende Sanitarie dell’Umbria, Via della Rivoluzione, 16, 06070, Ellera di Corciano, Perugia, Italy | 6/30/15 |
| 284676 | Italy | Galetta, Domenico | Comitato Etico Area 5 presso IRCCS Ospedale Oncologico di Bari Istituto Tumori Giovanni Paolo II, Viale Orazio Flacco 65, 70124, Bari, Puglia, Italy | 3/17/16 |

| | | | | |
|---|---|---|---|---|
| 280649 | Italy | Ciardiello, Fortunato | Comitato Etico dell'Università Federico II, Via Sergio Pansini, 5, 80131, Napoli, Napoli, Italy | 7/23/15 |
| 280480 | Italy | Borra, Roberta | Comitato Etico Azienda Ospedaliera Universitaria Maggiore della Carità, Corso Mazzini 18, 28100, Novara, Piemonte, Italy | 8/27/15 |
| 279874 | Italy | Migliorino, Maria | Comitato Etico Lazio 1, Circonvallazione Gianicolense, 87, 152, Roma, Lazio, Italy | 7/30/15 |
| 279866 | Italy | Cartenì, Giacomo | Comitato Etico Cardarelli-Santobono, Via Antonio Cardarelli 9, 80131, Napoli, Campania, Italy | 10/7/15 |
| 279785 | Latvia | Zvirbule, Zanete | The Ethics Committee for Clinical Trials of Medicinal Products, Aizkraukles Street 21-113, LV-1006, Riga, , Latvia | 4/24/15 |
| 279784 | Latvia | Purkalne, Gunta | The Ethics Committee for Clinical Trials of Medicinal Products, Aizkraukles Street 21-113, LV-1006, Riga, , Latvia | 4/24/15 |
| 281218 | Lithuania | Cicenas, Saulius | Lithuanian Bioethics Committee, Vilniaus str. 16, LT-01402, Vilnius, , Lithuania | 5/20/15 |
| 282504 | Mexico | Dominguez Andrade, Adriana | Comite de Etica en Investigacion de Mexico Centre for Clinical Research SA de CV, Amores 709, 3100, Ciudad de México, Distrito Federal, Mexico | 5/15/15 |
| 280831 | Mexico | Noriega Iriondo, Maria | Comité de Ética en Investigación de la Facultad de Medicina de la UANL y Hospital Universitario “Dr., Av. Francisco I. Madero y Gonzalitos S/N, Colonia Mitras Centro, 64460, Monterrey, Nuevo León, Mexico | 8/24/15 |
| 284134 | Netherlands | Dingemans, Anne-Marie | METC azM/UM, Oxfordlaan 10, 6202 AZ, Maastricht, , Netherlands | 10/29/15 |
| 281348 | Netherlands | Snijders, Dominic | METC Noord Holland, Nassauplein 10, 1815 GM, Alkmaar, , Netherlands | 10/29/15 |
| 281347 | Netherlands | Schramel, Franz | Medical Research Ethics Committees United, Koekoekslaan 1, 3435 CM, Nieuwegein, Utrecht, Netherlands | 12/8/15 |
| 279881 | Netherlands | Van den Borne, Ben | Medical Research Ethics Committees United, Koekoekslaan 1, 3435 CM, Nieuwegein, Utrecht, Netherlands | 10/29/15 |
| 279882 | Netherlands | Wilschut, Frank | Medical Research Ethics Committees United, Koekoekslaan 1, 3435 CM, Nieuwegein, Utrecht, Netherlands | 10/29/15 |
| 280825 | Netherlands | Hashemi, Sayed | Medical Research Ethics Committees United, Koekoekslaan 1, 3435 CM, Nieuwegein, Utrecht, Netherlands | 10/29/15 |

| | | | | |
|---|---|---|---|---|
| 281346 | Netherlands | Aerts, Joachim | Medical Research Ethics Committees United, Koekoekslaan 1, 3435 CM, Nieuwegein, Utrecht, Netherlands | 10/29/15 |
| 282503 | Netherlands | Buikhuisen, Wieneke | Medical Research Ethics Committees United, Koekoekslaan 1, 3435 CM, Nieuwegein, Utrecht, Netherlands | 10/29/15 |
| 283733 | Peru | Cisneros Tipismana, Rocio | Comite de Etica en Investigacion del Instituto Regional de Enfermedades Neoplasicas, Panamericana Norte Km. 558, 12345, Trujillo, , Peru | 10/14/15 |
| 283313 | Peru | Moron Escobar, Hernan | Asociacion Benefica Prisma, Calle Carlos Gonzles 251, Lima 32, Lima, Lima, Peru | 5/28/15 |
| 281315 | Peru | Kobashigawa, Alejandro | Comite de Etica en Investigacion del Hospital Guillermo Almenara Irigoyen, Avenida Grau 800, Lima 13, Lima, Lima, Peru | 10/14/15 |
| 279756 | Peru | Aleman Polanco, Diana Sofía | Asociacion Benefica Prisma, Calle Carlos Gonzles 251, Lima 32, Lima, Lima, Peru | 4/23/15 |
| 278307 | Peru | Mas Lopez, Luis | Comité Institucional de ética en Investigación Instituto Nacional de Enfermedades Neoplasicas, Avenida Angamos Este 2520, Lima34, Lima, Lima, Peru | 5/4/15 |
| 279930 | Portugal | Almodovar, Maria Teresa | Comissão de Ética para a Investigação Clínica - CEIC, Avenida do Brasil, 53, 1749-004- Lisboa, Lisboa, Portugal | 7/10/15 |
| 279931 | Portugal | Araújo, Antonio | Comissão de Ética para a Investigação Clínica - CEIC, Avenida do Brasil, 53, 1749-004- Lisboa, Lisboa, Portugal | 7/10/15 |
| 279932 | Portugal | Barata, Fernando | Comissão de Ética para a Investigação Clínica - CEIC, Avenida do Brasil, 53, 1749-004- Lisboa, Lisboa, Portugal | 7/10/15 |
| 279933 | Portugal | Queiroga, Henrique | Comissão de Ética para a Investigação Clínica - CEIC, Avenida do Brasil, 53, 1749-004- Lisboa, Lisboa, Portugal | 7/10/15 |
| 279934 | Portugal | Rodrigues, Ana | Comissão de Ética para a Investigação Clínica - CEIC, Avenida do Brasil, 53, 1749-004- Lisboa, Lisboa, Portugal | 7/10/15 |
| 279935 | Portugal | Teixeira, Encarnação | Comissão de Ética para a Investigação Clínica - CEIC, Avenida do Brasil, 53, 1749-004- Lisboa, Lisboa, Portugal | 7/10/15 |
| 281362 | Russian Federation | Stroyakovskiy, Daniil | Ethics Committee at Moscow City Oncology Hospital #62 of Moscow Healthcare Department, Krasnogorskiy district, Stepanovskoe, settlement Istra, 27, 143423, Moscow, , Russian Federation | 8/14/15 |
| 281361 | Russian Federation | Zhiltsova, Elena | Ethics Committee at Russian Medical Military Academy n.a. S.M.Kirov, Ulitsa Academika Lebedeva, 6, 194044, St. Petersburg, , Russian Federation | 9/22/15 |

| | | | | |
|---|---|---|---|---|
| 281360 | Russian Federation | Gorbunova, Vera | Ethics Committee at Russian Oncology Research Center n.a. N.N.Blokhin, Kashirskoe Shosse 24, 115478, Moscow, , Russian Federation | 9/8/15 |
| 281238 | Russian Federation | Galiulin, Rinat | Ethics Committee at Clinical Oncology Dispensary, Ulitsa Zavertyayeva, 9 - 1, 644013, Omsk, , Russian Federation | 8/4/15 |
| 281239 | Russian Federation | Karaseva, Nina | Ethics Committee at City Clinical oncologic dispensary, Vtoraya Beryozovaya Alleya 3/5, 197022, St. Petersburg, , Russian Federation | 8/4/15 |
| 281363 | Russian Federation | Kovalenko, Nadezhda | Ethics Committee at Volzhskiy regional clinical oncology dispensary #3, Ulitsa Komsomolskaya, 25, 404100, Volzhskiy, , Russian Federation | 6/27/16 |
| 280020 | Singapore | Soo, Ross | Domain Specific Review Board, Nexus@One-North (South Tower), 138543, Singapore, , Singapore | 4/9/15 |
| 278955 | Singapore | Lim, Darren | Singhealth Centralised Institutional Review Board, 7 Hospital Drive, Singhealth Office Of Research, Blk A, #03-01, Singhealth Research Facilities, 169611, Singapore, Singapore, Singapore | 3/10/15 |
| 283382 | Slovakia | Godal, Robert | Eticka komisia pri Narodnom onkologickom ustave, Klenova 1, 833 01, Bratislava, , Slovakia | 6/29/15 |
| 281215 | Slovakia | Kasan, Peter | Eticka komisia Univerzitna nemocnica Bratislava, Ruzinovska 6, 826 06, Bratislava, , Slovakia | 6/29/15 |
| 281213 | Slovakia | Beniak, Juraj | Eticka komisia Presovskeho samospravneho kraja, Namestie Mieru 2, 080 01, Presov, , Slovakia | 6/29/15 |
| 294325 | Spain | Fuentes Pradera, Jose | CEIC de la Corporacion Sanitaria del Parc Tauli, Calle Parc Tauli, s/n, 8208, Sabadell, Barcelona, Spain | 6/28/16 |
| 282505 | Spain | Palmero, Ramón | CEIC Hospital Universitari de Bellvitge, C/ Feixa Llarga s/n, 8907, L´Hospitalet de Llobregat, Cataluña, Spain | 6/9/15 |
| 280982 | Spain | Rodriguez-Abreu, Delvys | CEIC Hospital Universitario Insular Materno-Infantil de Las Palmas, Avenida Marítima del Sur, s/n, 35016, Las Palmas de Gran Canaria, , Spain | 6/9/15 |
| 280981 | Spain | Blasco Cordellat, Ana | CEIC Consorcio Hospital General Universitario de Valencia, Avenida Tres Cruces, 2, 46014, Valencia, Valencia, Spain | 6/9/15 |
| 280829 | Spain | Ponce Aix, Santiago | CEIC Hospital Universitario 12 de Octubre, Avenida de Cordoba, s/n, 28041, Madrid, Madrid, Spain | 6/9/15 |
| 280827 | Spain | Garrido Lopez, Pilar | CEIC Hospital Universitario Ramon y Cajal, Carretera de Colmenar km. 9.100, 28034, Madrid, Madrid, Spain | 6/9/15 |

| | | | | |
|---|---|---|---|---|
| 280826 | Spain | Felip Font, Enriqueta | CEIC Hospital Universitario Vall d'Hebrón, Passeig de la Vall d'Hebron, 119-129, 8035, Barcelona, , Spain | 6/9/15 |
| 280160 | Spain | Viñolas, Nuria | CEIC Hospital Clinic de Barcelona, Calle Villarroel, 170, 8036, Barcelona, Barcelona, Spain | 6/9/15 |
| 280159 | Spain | Vazquez Estevez, Sergio | CEIC de Galicia (CAEI), Edificio Administrativo San Lázaro, s/n, 15781, Santiago de Compostela, A Coruña, Spain | 6/9/15 |
| 280158 | Spain | Terrasa Pons, Josefa | CEIC Islas Baleares (CEIC-IB), Camí de Jesús, 38 A, 7011, Palma de Mallorca, Baleares, Spain | 6/9/15 |
| 280157 | Spain | Taus, Alvaro | CEIC Parc de Salut Mar, Calle Doctor Aiguader, 88, 8003, Barcelona, Barcelona, Spain | 6/9/15 |
| 280156 | Spain | Oramas, Juana | CEIC Hospital Universitario de Canarias, Calle Ofra, s/n - Planta -2, 38320, La Laguna, Santa Cruz de Tenerife, Spain | 6/9/15 |
| 280155 | Spain | Lopez Brea, Marta | CEIC de Cantabria, Avenida Cardenal Herrera Oria, s/n, 39011, Santander, Cantabria, Spain | 6/9/15 |
| 280154 | Spain | Insa Molla, Amelia | CEIC Hospital Clínico Universitario de Valencia, Avenida Vicente Blasco Ibánez, 17, 46010, Valencia, Valencia, Spain | 6/9/15 |
| 280153 | Spain | Jiménez Munarriz, Beatriz | CEIC Grupo Hospital de Madrid, Avenida Monteprincipe, 25, 28660, Boadilla del Monte, Madrid, Spain | 6/9/15 |
| 280152 | Spain | Gonzalez Larriba, Jose Luis | CEIC Hospital Clinico San Carlos, Calle Profesor Martin Lagos, s/n, 28040, Madrid, Madrid, Spain | 6/9/15 |
| 280151 | Spain | Alvarez, Rosa | CEIC Hospital General Universitario Gregorio Marañon, Calle Doctor Esquerdo, 46, 28007, Madrid, Madrid, Spain | 6/9/15 |
| 280150 | Spain | Garcia Campelo, Rosario | CEIC de Galicia (CAEI), Edificio Administrativo San Lázaro, s/n, 15781, Santiago de Compostela, A Coruña, Spain | 6/9/15 |
| 280148 | Spain | Domine Gomez, Manuel | CEIC Fundación Jiménez Díaz, Avenida Reyes Católicos, 2, 28040, Madrid, Madrid, Spain | 6/9/15 |
| 280147 | Spain | De Castro Carpeño, Javier | CEIC Hospital Universitario La Paz, Paseo de la Castellana, 261, 28046, Madrid, Madrid, Spain | 6/9/15 |
| 280146 | Spain | Barneto Aranda, Isidoro Carlos | CEIC de Andalucia (CCEIBA), Avenida de la Innovación s/n, 41020, Sevilla, Andalucía, Spain | 6/9/15 |

| | | | | |
|---|---|---|---|---|
| 280145 | Spain | Artal-Cortes, Angel Fernando | CEIC de Aragon (CEICA), Avenida San Juan Bosco, 13, 50009, Zaragoza, Zaragoza, Spain | 6/9/15 |
| 280144 | Spain | Majem Tarruella, Margarita | CEIC Hospital Santa Creu i Sant Pau, Avenida Sant Antoni Maria Claret, 167, 8025, Barcelona, Barcelona, Spain | 6/9/15 |
| 280143 | Spain | Garcia, Yolanda | CEIC de la Corporacion Sanitaria del Parc Tauli, Calle Parc Tauli, s/n, 8208, Sabadell, Barcelona, Spain | 6/9/15 |
| 280479 | Switzerland | Ochsenbein, Adrian | Kantonale Ethikkommission Bern (KEK), Murtenstraße 31, 3010, Bern, , Switzerland | 3/2/16 |
| 286358 | Taiwan, Province of China | Su, Ying Wen | Mackay Memorial Hospital Institutional Review Board, No.92, Section2, Chung-shan North Road, 104, Taipei, , Taiwan, Province of China | 8/17/15 |
| 280823 | Taiwan, Province of China | Liu, Chien-Ying | Chang Gung Medical Foundation, 199 Tung Hwa North Road, 10507, Taipei, , Taiwan, Province of China | 5/11/15 |
| 280505 | Taiwan, Province of China | Yu, Chong-Jen | Institution Review Board of National Taiwan University Hospital, No.1, Changde-de Street, Zhongzheng Dist, 100, Taipei, , Taiwan, Province of China | 6/22/15 |
| 278964 | Taiwan, Province of China | Chen, Yuh-Min | Institutional Review Board Taipei Veterans General Hospital, No. 201, Sec.2 Shipei Road, Beitou Dist., 11217, Taipei, , Taiwan, Province of China | 6/11/15 |
| 279753 | Taiwan, Province of China | Chen, Wei-Teing | Cheng-Hsin General Hospital Institutional Review Board, 1F, No. 45, Chenghsin Street, Beitou District, 112, Taipei City, , Taiwan, Province of China | 8/11/15 |
| 279752 | Taiwan, Province of China | Ho, Ching-Liang | Institutional Review Board of Tri-Service General Hospital, No.325,Section 2, Cheng-Kung Road, 11490, Taipei, , Taiwan, Province of China | 6/9/15 |
| 279751 | Taiwan, Province of China | Lin, Yu-Ching | Chang Gung Medical Foundation, 199 Tung Hwa North Road, 10507, Taipei, , Taiwan, Province of China | 5/11/15 |
| 279750 | Taiwan, Province of China | Ji, Bin-Chuan | Institutional Review Board, Changhua Christian Hospital, No.135 Nansiao Street, 50006, Changhua, , Taiwan, Province of China | 6/9/15 |
| 278968 | Taiwan, Province of China | Hung, Jen-Yu | Institutional Review Board Kaohsiung Medical University Chung-Ho Memorial Hospital, No.100, Tzyou 1st Road, 807, Kaohsiung City, , Taiwan, Province of China | 6/2/15 |
| 278965 | Taiwan, Province of China | Chang, Gee-Chen | The Institutional Review Board of Taichung Veterans General Hospital, No.160 Section 3 Chung-Kang Road, 40705, Taichung, , Taiwan, Province of China | 6/10/15 |
| 278960 | Taiwan, Province of China | Chen, Chao-Hsun | Institutional Review Board of the Chi Mei Medical Center, 4F, 3rd Medical building, No. 901 Chung-Huwa Rd., Young-Kang Dist. Tainan, Taiwan., , Tainan, , Taiwan, Province of China | 5/20/15 |

| | | | | |
|---|---|---|---|---|
| 278956 | Taiwan, Province of China | Hsia, Te-Chun | China Medical University and Hospital Research Ethics Committee, No.2, Yuh-Der Road, 40447, Taichung, , Taiwan, Province of China | 6/13/15 |
| 280760 | Ukraine | Vynnychenko, Ihor | CEQ of Regional Municipal Institution Sumy Regional Clinical Oncology Dispensary, Vulytsya Pryvokzalna 31, 40005, Sumy, , Ukraine | 3/2/15 |
| 280761 | Ukraine | Kobziev, Oleh | CEQ of Municipal Noncommercial Institution Regional Center of Oncology, Vulytsya Lisoparkivska 4, 61070, Kharkiv, , Ukraine | 3/20/15 |
| 280762 | Ukraine | Vasylyev, Leonid | CEQ of SI Institute of Medical Radiology n.a. S.P. Hryhoriev of NAMS of Ukraine, 82 Pushkinska str., 61024, Kharkiv, , Ukraine | 3/17/15 |
| 280763 | Ukraine | Shparyk, Yaroslav | CEQ of Lviv State Oncology Regional Treatment Diagnostic Center, 2-A Yaroslava Hasheka str., 79031, Lviv, , Ukraine | 3/18/15 |
| 280764 | Ukraine | Rusyn, Andriy | CEQ of Transcarpathian Regional Clinical Oncology Dispensary, Vulytsya Brodlakovycha, 2, 88014, Uzhgorod, , Ukraine | 3/10/15 |
| 280766 | Ukraine | Ivashchuk, Oleksandr | Commission of Ethics Questions on the basis of the Chernivtsi Regional Clinical Oncology Dispensary, Vulytsya Chervonoarmiyska 242, 58013, Chernivtsi, , Ukraine | 5/22/15 |
| 280767 | Ukraine | Hotko, Yevhen | Commission on Ethics Questions of MNPE Central City Clinical Hospital of Uzhhorod City Council, Vulytsya Gryboedova 20, 88000, Uzhgorod, , Ukraine | 3/3/15 |
| 280768 | Ukraine | Chornobai, Anatolii | CEQ of Poltava Regional Clinical Oncology Dispensary of Poltava Regional Council, 7a, Volodarskoho Str., 36021, Poltava, , Ukraine | 2/26/15 |
| 280769 | Ukraine | Bondarenko, Igor | LEC of Municipal Non-profit Enterprise "City Clinical Hospital # 4" of Dnipro City Council, Vulytsya Blyzhnya 31, 49102, Dnipropetrovsk, Dnipropetrovs'ka Oblast , Ukraine | 3/19/15 |
| 280770 | Ukraine | Andrusenko, Orest | CEQ of Treatment and Prevention Institution Volyn Regional Oncology Dispensary, Vulytsya Tymiryazeva 1, 43018, Lutsk, , Ukraine | 5/15/15 |
| 280771 | Ukraine | Adamchuk, Hryhoriy | CEQ of MI Kryvyi Rih Oncology Dispensary of Dnipropetrovsk Regional Council, 41 Dnipropetrovske Road, 50048, Kryvyi Rih, , Ukraine | 2/27/15 |
| 280821 | Ukraine | Goloborodko, Oleksandr | CEQ of MI of Zaporizhzhia Regional Council Zaporizhzhia Regional Clinical Oncology Dispensary, 177-A Kulturna str., 69040, Zaporizhzhia, Zaporiz'ka Oblast, Ukraine | 6/25/15 |
| 288092 | Ukraine | Shamrai, Volodymyr | Commission on Ethics Questions of Vinnytsya Regional Clinical Oncology Dispensary, 84 Khmelnytskyy prospekt, 21029, Vinnytsya, , Ukraine | 8/27/15 |
| 293804 | United States | MacKintosh, Frederick | University of Nevada Reno Biomedical Institutional Review Board, 1664 North Virginia Street, 89557, Reno, Nevada, United States | 7/21/16 |

| | | | | |
|---|---|---|---|---|
| 292662 | United States | Drew, David | Copernicus Group Independent Review Board, 1 Triangle Drive, 27709, Research Triangle Park, North Carolina, United States | 4/12/16 |
| 292661 | United States | Jamil, Rodney | Pinnacle Health Hospitals Institutional Review Board, 205 South Front Street, 17104, Harrisburg, Pennsylvania, United States | 4/12/16 |
| 288792 | United States | Rothschild, Neal | Western Institutional Review Board, 1019 39th Avenue Southeast, 98374, Puyallup, Washington, United States | 10/16/15 |
| 288791 | United States | Suga, Jennifer Marie | Kaiser Permanente Northern California Institutional Review Board, 1800 Harrison Street, 94162, Oakland, California, United States | 6/16/15 |
| 288789 | United States | Suga, Jennifer Marie | Kaiser Permanente Northern California Institutional Review Board, 1800 Harrison Street, 94162, Oakland, California, United States | 6/16/15 |
| 288788 | United States | Suga, Jennifer Marie | Kaiser Permanente Northern California Institutional Review Board, 1800 Harrison Street, 94162, Oakland, California, United States | 6/16/15 |
| 288787 | United States | Suga, Jennifer Marie | Kaiser Permanente Northern California Institutional Review Board, 1800 Harrison Street, 94162, Oakland, California, United States | 6/16/15 |
| 288786 | United States | Suga, Jennifer Marie | Kaiser Permanente Northern California Institutional Review Board, 1800 Harrison Street, 94162, Oakland, California, United States | 6/16/15 |
| 288785 | United States | Suga, Jennifer Marie | Kaiser Permanente Northern California Institutional Review Board, 1800 Harrison Street, 94162, Oakland, California, United States | 6/16/15 |
| 288784 | United States | Suga, Jennifer Marie | Kaiser Permanente Northern California Institutional Review Board, 1800 Harrison Street, 94162, Oakland, California, United States | 6/16/15 |
| 288783 | United States | Suga, Jennifer Marie | Kaiser Permanente Northern California Institutional Review Board, 1800 Harrison Street, 94162, Oakland, California, United States | 6/16/15 |
| 288782 | United States | Sullivan, Kevin | Western Institutional Review Board, 1019 39th Avenue Southeast, 98374, Puyallup, Washington, United States | 3/28/16 |
| 288781 | United States | Fabregas, Jesus | Copernicus Group Independent Review Board, 1 Triangle Drive, 27709, Research Triangle Park, North Carolina, United States | 10/21/15 |
| 288020 | United States | Patel, Pareshkumar | Western Institutional Review Board, 1019 39th Avenue Southeast, 98374, Puyallup, Washington, United States | 9/11/15 |
| 288018 | United States | Ikpeazu, Chukwuemeka | University Of Miami, 1500 N.w. 12th Avenue, 33136, Miami, Florida, United States | 12/1/15 |

| | | | | |
|---|---|---|---|---|
| 287057 | United States | Johns, Mark | US Oncology Inc. Institutional Review Board, 10101 Woodloch Forest, 77380, The Woodlands, Texas, United States | IEC since 30/Sep/2016 |
| 287057 | United States | Johns, Mark | Western Institutional Review Board, 1019 39th Avenue Southeast, 98374, Puyallup, Washington, United States | 9/29/15 |
| 287056 | United States | Daniel, Davey | Western Institutional Review Board, 1019 39th Avenue Southeast, 98374, Puyallup, Washington, United States | 9/3/15 |
| 287054 | United States | Arnaoutakis, Konstantinos | University of Arkansas IRB, 4301 W. Markham Street, 72205, Little Rock, Arkansas, United States | 6/29/16 |
| 287050 | United States | Alnsour, Mohammad | Mercy Saint Vincent Medical Center Institutional Review Board, 2213 Cherry Street, 43608, Toledo, Ohio, United States | 12/15/15 |
| 287048 | United States | Page, Ray | Western Institutional Review Board, 1019 39th Avenue Southeast, 98374, Puyallup, Washington, United States | 9/11/15 |
| 284314 | United States | Spira, Alexander | US Oncology Inc. Institutional Review Board, 10101 Woodloch Forest, 77380, The Woodlands, Texas, United States | 6/18/15 |
| 284312 | United States | Narang, Mohit | US Oncology Inc. Institutional Review Board, 10101 Woodloch Forest, 77380, The Woodlands, Texas, United States | 6/18/15 |
| 284133 | United States | Cetnar, Jeremy | Oregon Health & Science University IRB, 3181 S.W. Sam Jackson Park Road, 97239-3098, Portland, Oregon, United States | 2/12/16 |
| 284132 | United States | Socoteanu, Matei | US Oncology Inc. Institutional Review Board, 10101 Woodloch Forest, 77380, The Woodlands, Texas, United States | 6/18/15 |
| 283311 | United States | Herman, James | WIRB Copernicus Group, 1 Triangle Drive, 27709, Research Triangle Park, North Carolina, United States | 2/18/16 |
| 283308 | United States | Silberberg, Jeffrey | Copernicus Group Independent Review Board, 1 Triangle Drive, 27709, Research Triangle Park, North Carolina, United States | 5/28/15 |
| 283306 | United States | Nissenblatt, Michael | Copernicus Group Independent Review Board, 1 Triangle Drive, 27709, Research Triangle Park, North Carolina, United States | 6/1/15 |
| 283305 | United States | McCleod, Michael | Western Institutional Review Board, 1019 39th Avenue Southeast, 98374, Puyallup, Washington, United States | 9/4/15 |
| 283286 | United States | Hussein, Maen | Western Institutional Review Board, 1019 39th Avenue Southeast, 98374, Puyallup, Washington, United States | 9/11/15 |

| | | | | |
|---|---|---|---|---|
| 283284 | United States | Gurubhagavatula, Sarada | Copernicus Group Independent Review Board, 1 Triangle Drive, 27709, Research Triangle Park, North Carolina, United States | 5/28/15 |
| 282386 | United States | Wender, Donald | Siouxland Institutional Review Board, 230 Nebraska Street, 51101, Sioux City, Iowa, United States | 8/12/15 |
| 282385 | United States | Hoffman, Philip | University of Chicago Hospitals Institutional Review Board, 5751 South Woodlawn Avenue, 60637, Chicago, Illinois, United States | 8/4/15 |
| 282383 | United States | Gunturu, Krishna | Lahey Clinic, Inc. Institutional Review Board, 41 Mall Road, 1805, Boston, Massachusetts, United States | 3/9/16 |
| 282382 | United States | Seng, Sonia | New England Institutional Review Board, 85 Wells Avenue, 2459, Newton, Massachussets, United States | 3/31/16 |
| 282381 | United States | Hooberman, Arthur | Copernicus Group Independent Review Board, 1 Triangle Drive, 27709, Research Triangle Park, North Carolina, United States | 4/20/15 |
| 281314 | United States | Paschold, John | US Oncology Inc. Institutional Review Board, 10101 Woodloch Forest, 77380, The Woodlands, Texas, United States | 6/18/15 |
| 281313 | United States | Knapp, Mark | Copernicus Group Independent Review Board, 1 Triangle Drive, 27709, Research Triangle Park, North Carolina, United States | 6/15/15 |
| 280978 | United States | Sumrall, Bradley | Copernicus Group Independent Review Board, 1 Triangle Drive, 27709, Research Triangle Park, North Carolina, United States | 7/9/15 |
| 280977 | United States | Montero, Aldemar | Copernicus Group Independent Review Board, 1 Triangle Drive, 27709, Research Triangle Park, North Carolina, United States | 3/31/15 |
| 280977 | United States | Montero, Aldemar | Western Institutional Review Board, 1019 39th Avenue Southeast, 98374, Puyallup, Washington, United States | IEC since 24-Sep-2015 |
| 280976 | United States | Matrana, Marc | Ochsner Clinic Foundation Institutional Review Board, 1514 Jefferson Highway, 70121, New Orleans, Louisiana, United States | 5/12/15 |
| 280975 | United States | Koh, Han | Kaiser Permanente Southern California Institutional Review Board., 393 E. Walnut, 91188, Pasadena, California, United States | 5/19/15 |
| 280974 | United States | Kellum, Andrew | North Mississippi Health Services, 830 South Gloster Street, 38801, Tupelo, Mississippi, United States | 5/21/15 |
| 280973 | United States | Goueli, Basem | Saint Luke's Hospital Institutional Review Board, 915 East First Street, 55805, Duluth, Minnesota, United States | 9/1/15 |

| | | | | |
|---|---|---|---|---|
| 280972 | United States | Cho, Jonathan | Copernicus Group Independent Review Board, 1 Triangle Drive, 27709, Research Triangle Park, North Carolina, United States | 3/26/15 |
| 280971 | United States | Nikolinakos, Petros | Copernicus Group Independent Review Board, 1 Triangle Drive, 27709, Research Triangle Park, North Carolina, United States | 3/20/15 |
| 280185 | United States | Jotte, Robert | US Oncology Inc. Institutional Review Board, 10101 Woodloch Forest, 77380, The Woodlands, Texas, United States | 6/18/15 |
| 280181 | United States | Desai, Meghna | Springfield Committee for Research Involving Human Subjects (SCRIHS), 801 North Rutledge Street, 62702, Springfield, Illinois, United States | 11/11/15 |
| 280179 | United States | Rich (Thompson), Patricia | Western Institutional Review Board, 1019 39th Avenue Southeast, 98374, Puyallup, Washington, United States | 10/17/15 |
| 280177 | United States | Thomas, Christian | Copernicus Group Independent Review Board, 1 Triangle Drive, 27709, Research Triangle Park, North Carolina, United States | 5/8/15 |
| 280176 | United States | Stella, Philip | St. Joseph Mercy Health System Institutional Review Board #2 - Oncology Central IRB, 5301 East Huron River Drive, 48106, Ann Arbor, Michigan, United States | 12/18/15 |
| 280174 | United States | McCune, Steven | Copernicus Group Independent Review Board, 1 Triangle Drive, 27709, Research Triangle Park, North Carolina, United States | 10/1/15 |
| 280172 | United States | Nagasaka, Misako | Western Institutional Review Board, 1019 39th Avenue Southeast, 98374, Puyallup, Washington, United States | 7/10/15 |
| 280171 | United States | Finley, Gene | Copernicus Group Independent Review Board, 1 Triangle Drive, 27709, Research Triangle Park, North Carolina, United States | 6/16/15 |
| 280169 | United States | Suga, Jennifer Marie | Kaiser Permanente Northern California Institutional Review Board, 1800 Harrison Street, 94162, Oakland, California, United States | 6/16/15 |
| 280164 | United States | Czerlanis, Cheryl | Loyola University Institutional Review Board, 2160 South First Avenue, 60153, Maywood, Illinois, United States | 6/30/16 |
| 280163 | United States | Goodman, Michael | W.G. 'Bill' Hefner VA Medical Center, 1601 Brenner Avenue, 28144, Salisbury, North Carolina, United States | 3/23/16 |
| 280162 | United States | Kundra, Ajay | Western Institutional Review Board, 1019 39th Avenue Southeast, 98374, Puyallup, Washington, United States | 4/15/15 |
| 280161 | United States | Cohenuram, Michael | Copernicus Group Independent Review Board, 1 Triangle Drive, 27709, Research Triangle Park, North Carolina, United States | 3/13/15 |

| | | | | |
|---|---|---|---|---|
| 279749 | United States | Chitneni, Shobha | Copernicus Group Independent Review Board, 1 Triangle Drive, 27709, Research Triangle Park, North Carolina, United States | 1/29/15 |
| 279748 | United States | Tsai, Frank Yung-Chin | Western Institutional Review Board, 1019 39th Avenue Southeast, 98374, Puyallup, Washington, United States | 7/8/15 |
| 279747 | United States | Swanson, Paul | Copernicus Group Independent Review Board, 1 Triangle Drive, 27709, Research Triangle Park, North Carolina, United States | 2/20/15 |
| 279746 | United States | Subramanian, Janakiraman | Saint Luke's Hospital Institutional Review Board, 4401 Wornall Road, 64111, Kansas City, Missouri, United States | 6/8/15 |
| 279745 | United States | Mitchell, Reed | Copernicus Group Independent Review Board, 1 Triangle Drive, 27709, Research Triangle Park, North Carolina, United States | 1/27/15 |
| 279744 | United States | Kirshner, Eli | Western Institutional Review Board, 1019 39th Avenue Southeast, 98374, Puyallup, Washington, United States | 5/14/15 |
| 279743 | United States | Halibey, Bohdan | Copernicus Group Independent Review Board, 1 Triangle Drive, 27709, Research Triangle Park, North Carolina, United States | 5/29/15 |
| 279742 | United States | Goldschmidt, Jerome | Copernicus Group Independent Review Board, 1 Triangle Drive, 27709, Research Triangle Park, North Carolina, United States | 2/18/15 |
| 279741 | United States | DeVore, Russell | Copernicus Group Independent Review Board, 1 Triangle Drive, 27709, Research Triangle Park, North Carolina, United States | 6/3/15 |
| 279740 | United States | Chaudhry, Arvind | Copernicus Group Independent Review Board, 1 Triangle Drive, 27709, Research Triangle Park, North Carolina, United States | 1/30/15 |
| 279739 | United States | Belman, Neil | St. Luke’s Hospital & Health Network IRB, 801 Ostrum Street, 18015, Bethlehem, Pennsylvania, United States | 3/17/15 |
| 279736 | United States | Almubarak, Mohammed | Advarra Institutional Review Board, 6940 Columbia Gateway Drive, 21046, Columbia, Maryland, United States | 8/13/15 |
| 279735 | United States | Morris, John | Western Institutional Review Board, 1019 39th Avenue Southeast, 98374, Puyallup, Washington, United States | 2/19/16 |
| 279732 | United States | Kerr, Samuel | Lancaster General Hospital IRB, 555 North Duke Street, 17604, Lancaster, Pennsylvania, United States | 3/26/15 |
| 279731 | United States | Bailey, Samuel | Copernicus Group Independent Review Board, 1 Triangle Drive, 27709, Research Triangle Park, North Carolina, United States | 12/11/15 |

| | | | | |
|---|---|---|---|---|
| 279730 | United States | Coleman, Morton | Copernicus Group Independent Review Board, 1 Triangle Drive, 27709, Research Triangle Park, North Carolina, United States | 5/19/15 |
| 279729 | United States | Brzezniak, Christina | Walter Reed National Military Medical Center IRB, 503 Robert Grant Avenue, 20910-7500, Silver Spring, Maryland, United States | 1/7/16 |
| 279727 | United States | Ali, Muhammad | Copernicus Group Independent Review Board, 1 Triangle Drive, 27709, Research Triangle Park, North Carolina, United States | 4/3/15 |
| 278280 | United States | Shtivelband, Mikhail | Copernicus Group Independent Review Board, 1 Triangle Drive, 27709, Research Triangle Park, North Carolina, United States | 6/8/15 |
| 278279 | United States | Erickson, Brian | Copernicus Group Independent Review Board, 1 Triangle Drive, 27709, Research Triangle Park, North Carolina, United States | 2/25/15 |
| 278278 | United States | Burhani, Nafisa | Copernicus Group Independent Review Board, 1 Triangle Drive, 27709, Research Triangle Park, North Carolina, United States | 1/30/15 |
| 278274 | United States | Hamm, John | Western Institutional Review Board, 1019 39th Avenue Southeast, 98374, Puyallup, Washington, United States | 6/2/15 |
| 278272 | United States | Beck, Joseph | Copernicus Group Independent Review Board, 1 Triangle Drive, 27709, Research Triangle Park, North Carolina, United States | 1/7/15 |
| 278271 | United States | Sadiq, Ahad | Copernicus Group Independent Review Board, 1 Triangle Drive, 27709, Research Triangle Park, North Carolina, United States | 3/18/15 |
| 279734 | United States | Wilks, Sharon | Copernicus Group Independent Review Board, 5000 CentreGreen Way, 27513, Cary, North Carolina, United States | 2/9/15 |
| 279768 | United States | Martin, William | St Charles Medical Center, 2500 Northeast Neff Road, 97701, Bend, Oregon, United States | 5/7/15 |
| 288790 | United States | Suga, Jennifer Marie | Kaiser Permanente Northern California Institutional Review Board, 1800 Harrison Street, 94162, Oakland, California, United States | 6/16/15 |
