## Supplementary material for "Allelic Variation in *HLA-DRB1* is Associated with Development of Anti-Drug Antibodies in Cancer Patients Treated with Atezolizumab that are Neutralizing *in Vitro*": Ethics Committee approvals: IMpower132 EC IRB.pdf

### PROTOCOL GO29438 (IMPOWER132) – LIST OF IRBs/IECs

| Site ID | Country | Investigator | Institution | IRB/IEC Name | IRB/IEC Approval Date |
|---|---|---|---|---|---|
| 290062 | USA | Jaslowski, Anthony | St. Vincent Hospital | St. Vincent Hospital IRB | 14-Mar-16 |
| 290064 | USA | Santana-Davila, Rafael | University of Washington | Western Institutional Review Board | 9-Feb-16 |
| 290456 | USA | Del Prete, Salvatore | Stamford Hospital; BCC, MOHR | Copernicus Group Independent Review Board | 17-Dec-15 |
| 290695 | USA | Schneider, Bryan | University of Michigan | University of Michigan Medical School Institutional Review Board | 13-May-16 |
| 290698 | USA | Sanborn, Rachel | Providence Portland Medical Center | Providence Health and Services IRB | 18-Apr-16 |
| 290770 | Hungary | Kiraly, Zsolt | Veszprem Megyei Tudogyogyintezet | Egeszsegugyi Tudomanyos Tanacs Klinikai Farmakologiai Etikai Bizottsaga | 29-Mar-16 |
| 290815 | Portugal | Parente, Barbara | Hospital CUF Porto; Servico de Imunoalergologia | Comissão de Ética para a Investigação Clínica - CEIC | 18-Apr-16 |
| 290822 | Spain | Garcia Giron, Carlos | C.A.U de Burgos- Hospital Universitario de Burgos; Servicio de Oncologia | CEIC de Galicia (CAEI) | 12-Apr-16 |
| 290823 | Spain | Massuti Sureda, Bartomeu | Hospital General Univ. De Alicante | CEIC de Galicia (CAEI) | 12-Apr-16 |
| 290825 | Chile | Acevedo Gaete, Alejandro Andres | Hospital Clinico Vina del Mar | Comité Ético Científico del Servicio de Salud Viña del Mar Quillota-Hospital Gustavo Fricke | 15-Dec-16 |
| 290982 | USA | Johnson, Tirrell Tremayne | Orlando Health Inc. | Orlando Health Medical Center Institutional Review Board | 7-Apr-16 |
| 290983 | USA | Maclaughlin, William W. | Peninsula Cancer Institute | Copernicus Group Independent Review Board | 18-Feb-16 |
| 290985 | Australia | Lee, Chee | St George Hospital; Medical Oncology | St Vincent's Hospital Human Research Ethics Committee | 20-Jun-16 |
| 290986 | Hungary | Szalai, Zsuzsanna | Petz Aladar Megyei Oktato Korhaz | Egeszsegugyi Tudomanyos Tanacs Klinikai Farmakologiai Etikai Bizottsaga | 29-Mar-16 |
| 290988 | Spain | Muñoz Quintana, Miguel Angel | Instituto Valenciano Oncologia; Oncologia Medica | CEIC de Galicia (CAEI) | 12-Apr-16 |

| Site ID | Country | Investigator | Institution | IRB/ IEC Name | IRB/ IEC Approval Date |
|---|---|---|---|---|---|
| 290989 | Spain | Cobo Dols, Manuel | Hospital Regional Universitario de Malaga – Hospital General; Servicio de Neurologia | CEIC de Galicia (CAEI) | 12-Apr-16 |
| 290990 | Spain | Amenedo Gancedo, Margarita | Centro Oncologico de Galicia COG | CEIC de Galicia (CAEI) | 12-Apr-16 |
| 290992 | Netherlands | Custers, Frank L.J. | Zuyderland Medisch Centrum | Medical Research Ethics Committees United | 25-Oct-16 |
| 290993 | Belgium | Germonpré, Paul | AZ Maria Middelares | AZ Maria Middelares - Commissie voor Ethiek | 23-May-16 |
| 290995 | Belgium | Demedts, Ingel | AZ Delta (Campus Wilgenstraat), Apotheek | AZ Delta - Medisch Ethisch Comité | 23-May-16 |
| 290996 | Israel | Brenner, Ronen | Edith Wolfson Medical Center | Edith Wolfson MC Local EC | 8-Mar-16 |
| 290998 | Russia | Fadeeva, Natalia | Chelyabinsk Regional Clinical Oncology Dispensary | Ethics Committee at Chelyabinsk Regional Clinical Oncology Dispensary | 31-Aug-16 |
| 290999 | Taiwan | Chen, Chao-Hsun | Chi Mei Medical Center Liou Ying Campus | Institutional Review Board of the Chi Mei Medical Center | 28-Jun-16 |
| 291000 | Taiwan | Chian, Chih-Feng | Tri-Service General Hospital | Institutional Review Board of Tri-Service General Hospital | 7-Jun-16 |
| 291178 | Australia | Bigby, Kieron | Redcliffe Hospital | St Vincent's Hospital Human Research Ethics Committee | 20-Jun-16 |
| 291179 | Lithuania | Preimontiene, Daiva | Panevezys Hospital | Lithuanian Bioethics Committee | 18-Mar-16 |
| 291181 | Netherlands | Gans, Stephanus Jacobus Maria | Ziekenhuis St. Jansdal | Medical Research Ethics Committees United | 25-Oct-16 |
| 291185 | Spain | Gurpide Ayarra, Luis Alfonso | Clinica Universitaria de Navarra; Servicio de Oncologia | CEIC de Galicia (CAEI) | 12-Apr-16 |
| 291186 | Spain | Gonzalez Cao, Maria | Hospital Universitari Dexeus - Grupo Quironsalud; Servicio de Oncologia Medica | CEIC de Galicia (CAEI) | 12-Apr-16 |
| 291187 | Spain | Lahuerta Martinez, Ainhara | Onkologikoa; Ensayos Clinicos | CEIC de Galicia (CAEI) | 12-Apr-16 |
| 291188 | Taiwan | Shih, Jin-Yuan | National Taiwan Uni Hospital; Internal Medicine | Research Ethics Committee of National Taiwan University Hospital | 11-May-16 |

| Site ID | Country | Investigator | Institution | IRB/ IEC Name | IRB/ IEC Approval Date |
|---|---|---|---|---|---|
| 291194 | USA | Russell, Karen | Tallahassee Memorial Hospital | Tallahassee Memorial HealthCare Institutional Review Board | 26-Aug-16 |
| 291195 | USA | Reddy, Sreekanth | Northside Hospital | WIRB Copernicus Group | 11-May-16 |
| 291196 | USA | West, Howard | Swedish Cancer Institute | Western Institutional Review Board (WIRB) | 6-Apr-16 |
| 291231 | Ukraine | Ursol, Grygoriy | Private Enterprise Private Manufacturing Company Acinus | CEQ of Private Enterprise Private Manufactire Company Acinus | 6-Sep-16 |
| 291259 | Russia | Kislov, Nikolay | Regional Clinical Oncology Hospital | Independent Interdisciplinary Committee at Clinical Trials Ethical Expertise | 26-Sep-16 |
| 291260 | Belgium | Bosquee, Léon | Clinique André Renard; Pneumologie | Clinique André Renard - Comité d'Ethique | 23-May-16 |
| 291444 | USA | Peguero, Julio | Oncology Consultants PA | Copernicus Group Independent Review Board | 14-Jan-16 |
| 291445 | USA | Arnold, Susanne | University of Kentucky; Markey Cancer Center | University of Kentucky Institutional Review Board | 9-Mar-16 |
| 291494 | Australia | Dear, Rachel | St Vincent's Hospital Sydney | St Vincent's Hospital Human Research Ethics Committee | 20-Jun-16 |
| 291495 | Australia | Marx, Gavin | Sydney Adventist Hospital; Clinical Trial Unit | St Vincent's Hospital Human Research Ethics Committee | 20-Jun-16 |
| 291592 | Hungary | Csejtei, Andras | Markusovszky Hospital | Egeszsegugyi Tudomanyos Tanacs Klinikai Farmakologiai Etikai Bizottsaga | 29-Mar-16 |
| 291710 | Australia | Brown, Stephen | Ballarat Health Services | St Vincent's Hospital Human Research Ethics Committee | 20-Jun-16 |
| 291711 | France | Dubray Longeras, Pascale | Centre Jean Perrin Centre Regional de Lutte Contre Le Cancer D'Auvergne | CPP Sud-Méditerranée 1 | 8-Jun-16 |
| 291712 | France | Genet, Dominique | Polyclinique de Limoges - Site Chenieux; Oncologie Medicale | CPP Sud-Méditerranée 1 | 8-Jun-16 |
| 291713 | Italy | Ferrau, Francesco | Ospedale San Vincenzo Taormina :Divisione di Oncologia Medica | Comitato Etico Interaziendale della Provincia di Messina | 23-May-16 |

| Site ID | Country | Investigator | Institution | IRB/ IEC Name | IRB/ IEC Approval Date |
|---|---|---|---|---|---|
| 291714 | Italy | Novello, Silvia | Azienda Sanitaria Ospedaliera S Luigi Gonzaga; S.C.D.U. di Oncologia Toracica | Comitato Etico Interaziendale A.O.U. San Luigi Gonzaga di Orbassano | 20-May-16 |
| 291715 | Italy | Tiseo, Marcello | Azienda Ospedaliero Universitaria di Parma | Comitato Etico Unico per la Provincia di Parma | 29-Nov-16 |
| 291716 | Malaysia | Appalanaido, Gokula Kumar | Advanced Medical and Dental Institute; Kompleks Klinikal | Research Ethics Committee (JEPeM) | 17-Nov-16 |
| 291719 | Portugal | Estevinho, Fernanda | Unidade Local de Saude de Matosinhos SA | Comissão de Ética para a Investigação Clínica - CEIC | 18-Apr-16 |
| 291724 | Romania | Udrea, Anghel Adrian | Medisprof SRL | National Bioethics Committee for Medicine and Medical Devices | 26-Sep-16 |
| 291726 | Romania | Mazilu, Laura | Spitalul Clinic Judetean de Urgenta Sf. Apostol Andrei Constanta | National Bioethics Committee for Medicine and Medical Devices | 26-Sep-16 |
| 291732 | Argentina | Pilnik, Norma | Fundacion Clinica Colombo | Comité Institucional de Etica de Investigación en Salud Clínica Colombo | 4-Mar-16 |
| 291734 | Netherlands | Kloover, Jeroen | ETZ Elisabeth | Medical Research Ethics Committees United | 25-Oct-16 |
| 291784 | Spain | Alonso, Jose Luis | Hospital Universitario Virgen de La Arrixaca; Servicio De Oncologia | CEIC de Galicia (CAEI) | 12-Apr-16 |
| 291785 | Spain | Pardo Aranda, Nuria | Hospital Univ Vall d'Hebron; Servicio de Oncologia | CEIC de Galicia (CAEI) | 12-Apr-16 |
| 291788 | USA | Seneviratne, Lasika | Los Angeles Hematology Oncology Medical Group | Copernicus Group Independent Review Board | 11-Feb-16 |
| 291840 | Ukraine | Vynnychenko, Ihor | Regional Municipal Institution Sumy Regional Clinical Oncology Dispensary; Oncothoracic department | CEQ of Regional Municipal Institution Sumy Regional Clinical Oncology Dispensary | 10-May-16 |
| 291842 | UK | Steele, Nicola | Gartnavel General Hospital; Beatson West of Scotland Cancer Centre | NRES Committee South Central – Oxford C | 29-Apr-16 |
| 291843 | UK | Benepal, Tim | St George's Hospital | NRES Committee South Central – Oxford C | 29-Apr-16 |
| 292072 | Israel | Charkovsky, Tatiana | Barzilai Medical Center | Barzilai Medical Center Local EC | 18-Apr-16 |

| Site ID | Country | Investigator | Institution | IRB/ IEC Name | IRB/ IEC Approval Date |
|---|---|---|---|---|---|
| 292077 | Portugal | Felizardo, Margarida | Hospital Beatriz Angelo | Comissão de Ética para a Investigação Clínica - CEIC | 18-Apr-16 |
| 292137 | Spain | Lianes Barragan, Pilar | Hospital de Mataro | CEIC de Galicia (CAEI) | 12-Apr-16 |
| 292138 | Spain | Garcia Garcia, Yolanda | Corporacio Sanitaria Parc Tauli; Servicio de Oncologia | CEIC de Galicia (CAEI) | 12-Apr-16 |
| 292139 | Spain | Guirado Risueño, Maria | Hospital General Universitario de Elche; Servicio de Oncologia | CEIC de Galicia (CAEI) | 12-Apr-16 |
| 292140 | Spain | Hernández Marin, Berta | Complejo Hospitalario de Navarra; Servicio de Oncologia | CEIC de Galicia (CAEI) | 12-Apr-16 |
| 292142 | Ukraine | Vereshchako, Roman | Kyiv Railway Clinical Hospital #3 of Branch Health Center of PJSC Ukrainian Railway; Surgery Dept | CEQ of Kyiv Railway Clinical Hospital #3 of Branch Health Center of the PJSC Ukrainian Railway | 7-Jun-16 |
| 292233 | USA | Rao, Subramanya | HealthCare Research Network II, LLC - PPDS | Copernicus Group Independent Review Board | 20-Apr-16 |
| 292234 | USA | Langdon, Robert | Nebraska Methodist Hospital | Nebraska Methodist Hospital Institutional Review Board | 23-May-16 |
| 292315 | UK | Lewanski, Conrad | Charing Cross Hospital | NRES Committee South Central – Oxford C | 29-Apr-16 |
| 292316 | USA | Lee, Wes | St. Joseph Heritage Healthcare | St. Joseph Health IRB | 8-Mar-16 |
| 292325 | UK | Ball, Simon | Queen's Hospital | NRES Committee South Central – Oxford C | 29-Apr-16 |
| 292326 | Italy | Baldini, Editta | Ospedale San Luca - USL2 Lucca | Comitato Etico Area Vasta Nord Ovest presso Azienda Ospedaliero Universitaria Pisana di Pisa | 29-Jun-16 |
| 292468 | Peru | Acevedo Zanabria, Carmen Laura | Hospital Nacional Cayetano Heredia | Comite Institucional de Etica en la Investigacion del Hospital Nacional Cayetano Heredia | 8-Apr-16 |
| 292541 | Chile | Yanez, Eduardo | Instituto Clínico Oncologico del Sur | Comité Ético-Científico del Servicio de Salud Metropolitano Oriente (CEC SSMO) | 13-Sep-16 |

| Site ID | Country | Investigator | Institution | IRB/ IEC Name | IRB/ IEC Approval Date |
|---|---|---|---|---|---|
| 292605 | Hungary | Lantos, Ákos | Tudogyogyintezet Torokbalint | Egeszsegugyi Tudomanyos Tanacs Klinikai Farmakologiai Etikai Bizottsaga | 29-Mar-16 |
| 292607 | Spain | Lopez Criado, Maria Pilar | MD Anderson Cancer Center | CEIC de Galicia (CAEI) | 1-Jun-16 |
| 292634 | France | Hilgers, Werner | Institut Sainte Catherine | CPP Sud-Méditerranée 1 | 8-Jun-16 |
| 292635 | France | Quantin, Xavier | Centre Regional de Lutte contre le Cancer Val d Aurelle - Paul Lamarque; Service d oncologie | CPP Sud-Méditerranée 1 | 8-Jun-16 |
| 292638 | Romania | Volovat, Constantin | Euroclinic Center of Oncology SRL | National Bioethics Committee for Medicine and Medical Devices | 26-Sep-16 |
| 292642 | Ukraine | Paramonov, Victor | MICR Oncology Dispensary of Cherkasy Regional Council; Regional Center of Clinical Oncology | CEQ of MI Cherkasy Regional Oncology Dispensary of Cherkasy Regional Council | 19-May-16 |
| 292643 | Ukraine | Kolesnik, Oleksii | MI Dnipropetrovsk City Multifield Clinical Hospital 4 of Dnipropetrovsk Regional Council | CEQ of MI of Zaporizhzhia Regional Council Zaporizhzhia Regional Clinical Oncology Dispensary | 5-May-16 |
| 292644 | France | Mastroianni, Benedicte | Hopital Louis Pradel; Pneumologie | CPP Sud-Méditerranée 1 | 8-Jun-16 |
| 292669 | Netherlands | Codrington, H.E. | Haga Ziekenhuis | Medical Research Ethics Committees United | 25-Oct-16 |
| 292682 | Taiwan | Wei, Yu-Feng | E-DA Hospital; Chest | Institutional Review Board, E-DA Hospital | 6-Jul-16 |
| 292744 | USA | Copur, Mehmet | CHI Health St. Francis | Copernicus Group Independent Review Board | 29-Apr-16 |
| 292830 | Argentina | Batagelj, Emilio | Hospital Militar Central Cirujano Mayor Dr. Cosme Argerich | Comité Institucional de Revisión de Ensayos Clínicos | 16-Aug-16 |
| 292832 | France | Barlesi, Fabrice | Hopital Nord AP-HM | CPP Sud-Méditerranée 1 | 8-Jun-16 |
| 292955 | UK | Talbot, Toby | Royal Cornwall Hospital | NRES Committee South Central – Oxford C | 29-Apr-16 |
| 292956 | UK | MacGregor, Carol | Raigmore Hospital | NRES Committee South Central – Oxford C | 29-Apr-16 |
| 292957 | Italy | Cognetti, Francesco | Istituto Nazionale Tumori Regina Elena | CE IRCCS Lazio Comitato Etico IRCCS Istituti Fisioterapici Ospitalieri | 20-Jul-16 |
| 292959 | Italy | Romano, Gianpiero | Presidio Ospedaliero Vito Fazzi; Unita Operativa Di Oncologia Medica | Comitato Etico della AUSL LE di Lecce | 2-Jan-17 |

| Site ID | Country | Investigator | Institution | IRB/IEC Name | IRB/IEC Approval Date |
|---|---|---|---|---|---|
| 292960 | Italy | Maiello, Evaristo | Ospedale Casa Sollievo Della Sofferenza IRCCS | Sezione del Comitato Etico IRCCS Ist. Tumori - Giovanni Paolo II di Bari c/o Fondazione Casa Solliev | 6-Sep-16 |
| 293018 | Republic of Korea | Lee, Se-Hoon | Samsung Medical Center | Samsung Medical Center Institutional Review Board | 11-Aug-16 |
| 293023 | USA | Kio, Ebenezer | Goshen Health System | Western Institutional Review Board (WIRB) | 25-Mar-16 |
| 293165 | Spain | Fuentes Pradera, Jose | Complejo Hospitalario Nuestra Senora de Valme | CEIC de Galicia (CAEI) | 2-Jun-16 |
| 293166 | Japan | Okazaki, Tatsuma | Tohoku University Hospital | National University Corporation Tohoku University Tohoku University Hospital IRB | 23-May-16 |
| 293167 | Japan | Nishio, Makoto | The Cancer Institute Hospital of JFCR | The Cancer Institute Hospital of JFCR Institutional Review Board | 28-Jun-16 |
| 293168 | Japan | Kubota, Kaoru | Nippon Medical School Hospital | Nippon Medical School Hospital Institutional Review Board | 8-Jun-16 |
| 293169 | Japan | Fujita, Yuka | National Hospital Organization Asahikawa Medical Center | National Hospital Organization Asahikawa Medical Center Institutional Review Board | 19-Apr-16 |
| 293170 | Japan | Goto, Koichi | National Cancer Center Hospital East | National Cancer Center Hospital IRB | 28-Sep-16 |
| 293171 | Japan | Saito, Haruhiro | Kanagawa Cancer Center | Kanagawa Cancer Center Institutional Review Board | 10-May-16 |
| 293172 | Japan | Kasahara, Kazuo | Kanazawa University Hospital | Kanazawa University Hospital IRB | 14-Apr-16 |
| 293173 | Japan | Saka, Hideo | National Hospital Organization Nagoya Medical Center | National Hospital Organization Nagoya Medical Center IRB | 25-May-16 |
| 293174 | Japan | Fujisaka, Yasuhito | Osaka Medical College Hospital | Osaka Medical College Hospital IRB | 31-May-16 |
| 293175 | Japan | Kida, Hiroshi | Osaka University Hospital | Osaka University Hospital IRB | 24-May-16 |
| 293176 | Japan | Usui, Kazuhiro | NTT Medical Center Tokyo | NTT Medical Center Tokyo Institutional Review Board | 19-Apr-16 |

| Site ID | Country | Investigator | Institution | IRB/ IEC Name | IRB/ IEC Approval Date |
|---|---|---|---|---|---|
| 293177 | Japan | Chikamori, Kenichi | National Hospital Organization Yamaguchi - Ube Medical Center | National Hospital Organization Yamaguchi-Ube Medical Center Institutional Review Board | 27-Apr-16 |
| 293178 | Japan | Okada, Morihito | Hiroshima University Hospital | Hiroshima University Hospital IRB | 4-Apr-16 |
| 293179 | Japan | Nishioka, Yasuhiko | Tokushima University Hospital | Tokushima University Hospital Institutional Review Board | 31-May-16 |
| 293180 | Japan | Aragane, Naoko | Saga University Hospital | Saga University Hospital IRB | 9-May-16 |
| 293181 | Japan | Watanabe, Satoshi | Niigata University Medical & Dental Hospital | Niigata University Medical & Dental Hospital IRB | 25-May-16 |
| 293182 | Japan | Okuma, Yusuke | Tokyo Metropolitan Cancer and Infectious diseases Center Komagome Hospital | Tokyo Metropolitan Komagome Hospital IRB | 28-Apr-16 |
| 293183 | Japan | Nakahara, Yasuharu | National Hospital Organization Himeji Medical Center | Institutional Review Board of National Hospital Organization Himeji Medical Center | 15-Apr-16 |
| 293312 | Spain | Bosch Barrera, Joaquim | Hospital Universitari de Girona Dr Josep Trueta; Departamento de Oncologia Medica | CEIC de Galicia (CAEI) | 1-Jun-16 |
| 293314 | Australia | Singh, Madhu | Barwon Health | St Vincent's Hospital Human Research Ethics Committee | 20-Jun-16 |
| 293324 | Japan | Mizuno, Keiko | Kagoshima University Hospital | Kagoshima University Medical And Dental Hospital Institutional Review Board | 30-May-16 |
| 293624 | Japan | Kato, Motoyasu | Juntendo University Hospital | Juntendo University Hospital IRB | 27-May-16 |
| 293769 | UK | Chitnis, Meenali | Churchill Hospital | South Central - Oxford C Research Ethics Committee | 29-Apr-16 |
| 293770 | USA | Finley, Gene | Allegheny Cancer Center | Western Institutional Review Board (WIRB) | 20-Mar-17 |
| 293771 | Australia | Jain, Vikram | Mater Adult Hospital | St Vincent's Hospital Human Research Ethics Committee | 20-Jun-16 |
| 293772 | Hungary | Papai-Szekely, Zsolt | Fejer Megyei Szent Gyorgy Egyetemi Oktato Korhaz | Egeszsegugyi Tudomanyos Tanacs Klinikai Farmakologiai Etikai Bizottsaga | 25-May-16 |

| Site ID | Country | Investigator | Institution | IRB/ IEC Name | IRB/ IEC Approval Date |
|---|---|---|---|---|---|
| 293773 | Spain | Juan Vidal, Oscar | Hospital Universitari i Politecnic La Fe de Valencia | CEIC de Galicia (CAEI) | 28-Oct-16 |
| 293775 | Taiwan | Chiu, Chao-Hua | Taipei Veterans General Hospital | Institutional Review Board Taipei Veterans General Hospital | 15-Jun-16 |
| 293777 | Republic of Korea | Kim, Dong-Wan | Seoul National University Hospital | Seoul National University Hospital IRB | 4-Aug-16 |
| 293833 | Malaysia | Thiagarajan, Muthukkumaran | Hospital Kuala Lumpur | Medical Research Ethic Committee Institute for Health Management | 17-Jun-16 |
| 293870 | Latvia | Zvirbule, Zanete | Riga East Clinical University Hospital Latvian Oncology Centre | The Ethics Committee for Clinical Trials of Medicinal Products | 8-Jul-16 |
| 294121 | France | Monnet, Isabelle | Centre Hospitalier Intercommunal; Service de Pneumologie | CPP Sud-Méditerranée 1 | 8-Jun-16 |
| 294124 | Italy | Cortesi, Enrico | Azienda Policlinico Umberto I | Comitato Etico Azienda Policlinico Umberto I | 16-Jan-17 |
| 294126 | Spain | Jimenez Munarriz, Beatriz | HM Sanchinarro – CIOCC | CEIC de Galicia (CAEI) | 1-Jun-16 |
| 294128 | Republic of Korea | Lee, Dae Ho | Asan Medical Center | Asan Medical Center Institutional Review Board | 22-Aug-16 |
| 294324 | UK | Roy, Amy | Derriford Hospital; Plymouth Oncology Centre | South Central - Oxford C Research Ethics Committee | 29-Apr-16 |
| 294469 | USA | Sadiq, Ahad | Fort Wayne Med Oncology & Hematology Inc | Copernicus IRB | 21-Nov-16 |
| 294498 | France | Debieuvre, Didier | Centre Hospitalier de Mulhouse - Hopital Emile Muller | CPP Sud-Méditerranée 1 | 8-Jun-16 |
| 294500 | Netherlands | van der Leest, Kornelis | Amphia Ziekenhuis | Medical Research Ethics Committees United | 6-Jan-17 |
| 294583 | Russia | Stroyakovskii, Daniil | Moscow City Oncology Hospital #62 | Ethics Committee at Moscow City Oncology Hospital #62 of Moscow Healthcare Department | 28-Nov-16 |
| 295125 | Taiwan | Lai, Chun-Liang | Buddhist Dalin Tzuchi General Hospital | Tzu Chi General Hospital Research Ethics Committee | 16-Sep-16 |

| Site ID | Country | Investigator | Institution | IRB/ IEC Name | IRB/ IEC Approval Date |
|---|---|---|---|---|---|
| 295586 | Argentina | Kowalyszyn, Rubén Dario | Centro de Investigacion; Clinica - Clinica Viedma S.A. | Comité Independiente de Etica en investigación clínica "Dr. Carlos A. Barclay | 7-Jul-16 |
| 295711 | Ireland | Higgins, Michaela | Mater Misecordiae University Hospital | Clinical Research Ethics Committee Of The Cork Teaching Hospitals | 13-Feb-17 |
| 295715 | Ireland | Cuffe, Sinead | St James's Hospital | Clinical Research Ethics Committee Of The Cork Teaching Hospitals | 20-Feb-17 |
| 295716 | Italy | Cappuzzo, Federico | Ospedale Santa Maria Delle Croci | Comitato Etico IRST IRCCS e Area Vasta Romagna | 21-Feb-17 |
| 295745 | Spain | Moran Bueno, Teresa | Hospital Universitario Germans Trias i Pujol | CEIC de Galicia (CAEI) | 28-Oct-16 |
| 295748 | UK | Brewster, Alison | Velindre Hospital | NRES Committee South Central – Oxford C | 14-Jun-16 |
| 295749 | USA | Khair, Tina | Gettysburg Cancer Center | Copernicus Group Independent Review Board | 17-Jun-16 |
| 295750 | USA | Goldschmidt-Jr, Jerome | Blue Ridge Cancer Care | Copernicus Group Independent Review Board | 30-Jun-16 |
| 295795 | Chile | Orlandi, Franscico | Health & Care SPA | Comité Ético-Científico del Servicio de Salud Metropolitano Oriente (CEC SSMO) | 11-Oct-16 |
| 295796 | Ukraine | Bondarenko, Igor | MI Dnipropetrovsk City Multifield Clinical Hospital 4 of Dnipropetrovsk Regional Council | CEQ of MI Dnipropetrovsk City Multifield Clinical Hospital #4 of Dnipropetrovsk Regional Council | 1-Sep-16 |
| 296733 | Spain | Garrido Lopez, Pilar | Hospital Universitario Ramon y Cajal | CEIC de Galicia (CAEI) | 28-Oct-16 |
| 296734 | Spain | Nadal, Ernest | ICO L'Hospitalet; Servicio de oncologia medica | CEIC de Galicia (CAEI) | 28-Oct-16 |
| 296735 | Spain | Majem Tarruella, Margarita | Hospital de la Santa Creu i Sant Pau | CEIC de Galicia (CAEI) | 28-Oct-16 |
| 296739 | USA | Cheng, Haiying | Montefiore Medical Center | BRANY IRB | 28-Dec-16 |
| 296743 | Australia | Ganju, Vinod | Peninsula and South Eastern Haematology and Oncology Grou | Bellberry Human Research Ethics Committee | 27-Jan-17 |
| 296746 | Netherlands | de Jong, Wouter | Ziekenhuis Gelderse Vallei | Medical Research Ethics Committees United | 8-Aug-17 |

| Site ID | Country | Investigator | Institution | IRB/ IEC Name | IRB/ IEC Approval Date |
|---|---|---|---|---|---|
| 297185 | Australia | Nott, Louise | Royal Hobart Hospital | Tasmania Health and Medical Human Research Ethics Committee | 19-Dec-16 |
| 297613 | USA | Wallace, James | Ingalls Memorial Hospital | Ingalls Memorial Hospital IRB | 20-Oct-16 |
| 297699 | France | Berard, Henri | Hopital d Instruction des Armees de Sainte Anne | CPP Sud-Méditerranée 1 | 14-Sep-16 |
| 297702 | UK | Ayre, Gareth | Bristol Haematology and Oncology Centre | NRES Committee South Central – Oxford C | 29-Apr-16 |
| 297704 | Spain | Vazquez Estevez, Sergio | Hospital Lucus Augusti; Servicio de Oncologia | CEIC de Galicia (CAEI) | 28-Oct-16 |
| 297708 | Israel | Zer, Alona | Rabin Medical Center | Institutional Helsinki Committee, Rabin Medical Center | 8-Feb-17 |
| 298355 | Bulgaria | Koynov, Krasimir | Multiprofile Hospital for Active Treatment Serdika EOOD | Ethics Committee for Multi-Centre Trials | 25-Jan-17 |
| 298356 | Austria | Eckmayr, Josef | Klinikum Wels-Grieskirchen GmbH | Ethik-Kommission der Medizinischen Universität Wien und des Allgemeinen Krankenhauses der Stadt Wien | 14-Dec-16 |
| 299739 | USA | Spira, Alexander I. | Virginia Cancer Specialists (Fairfax) - USOR | Copernicus Group Independent Review Board | 9-Feb-17 |
| 301595 | USA | Gerstner, Gregory J. | Illinois Cancer Care | Copernicus Group Independent Review Board | 1-Feb-17 |
| 302325 | France | Helissey, Carole | Hopital d'Instruction des Armees de Begin | CPP Sud-Méditerranée 1 | 8-Mar-17 |
