## Supplementary material for "Allelic Variation in *HLA-DRB1* is Associated with Development of Anti-Drug Antibodies in Cancer Patients Treated with Atezolizumab that are Neutralizing *in Vitro*": Ethics Committee approvals: IMpower133 EC IRB.pdf

| Site # | Investigator | EC/IRB Name and Address | IRB/EC Approval |
|---|---|---|---|
| 290929 | Navarro, Alejandro | Hospital Ramon y Cajal ;Comité Etico de Investigación Clínica, Ctra. Colmenar Viejo, km 9,1, 28034, Madrid, MADRID, SPAIN | 29-Jun-2016 |
| 290930 | Isla Casado, Dolores | Hospital Ramon y Cajal ;Comité Etico de Investigación Clínica, Ctra. Colmenar Viejo, km 9,1, 28034, Madrid, MADRID, SPAIN | 29-Jun-2016 |
| 290931 | De Castro Carpeno, Javier | Hospital Ramon y Cajal ;Comité Etico de Investigación Clínica, Ctra. Colmenar Viejo, km 9,1, 28034, Madrid, MADRID, SPAIN | 29-Jun-2016 |
| 290932 | Olmedo, Maria Eugenia | Hospital Ramon y Cajal ;Comité Etico de Investigación Clínica, Ctra. Colmenar Viejo, km 9,1, 28034, Madrid, MADRID, SPAIN | 29-Jun-2016 |
| 290933 | Trigo Perez, Jose Manuel | Hospital Ramon y Cajal ;Comité Etico de Investigación Clínica, Ctra. Colmenar Viejo, km 9,1, 28034, Madrid, MADRID, SPAIN | 29-Jun-2016 |
| 291027 | Lamprecht, Bernd | Ethikkommission d. Landes Oberösterreich, Wagner-Jauregg-Weg 15, 4020, Linz, AUSTRIA | 27-Jul-2016 |
| 291146 | Ahn, Myung-Ju | Samsung Medical Center EC, 81, Irwon-ro, Gangnam-gu, 06351, Seoul, KOREA, REPUBLIC OF | 30-Jun-2016 |
| 291147 | Kim, Dong-Wan | Seoul National University Hospital; IRB, 101, Daehak-ro, Jongno-gu, 03080, Seoul, KOREA, REPUBLIC OF | 07-Jun-2016 |
| 291154 | KIM, SANG-WE | Asan Medical Center Ethics Committee; Asan Medical Center; IRB | 28-Jun-2016 |
| 291448 | SAMANTAS, EPAMINONDAS | National Ethics Committee, Ministry of Health and Social Wellfare, 284, Messogion Avenue, 15562, Cholargos, GREECE | 21-Jul-2016 |
| 291450 | Syrigos, Konstantinos | National Ethics Committee, Ministry of Health and Social Wellfare, 284, Messogion Avenue, 15562, Cholargos, GREECE | 28-Jul-2016 |
| 291451 | KALOFONOS, HARALABOS | National Ethics Committee, Ministry of Health and Social Wellfare, 284, Messogion Avenue, 15562, Cholargos, GREECE | 27-Jul-2016 |
| 291477 | Garassino, Marina | Comitato Etico Indipendente della Fondazione IRCCS Istituto Nazionale dei Tumori di Milano, Via Giacomo Veneziani 1, 20133, Milano, Lombardia, ITALY | 11-Aug-2016 |
| 291478 | Tiseo, Marcello | Comitato Etico per Parma, Via Gramsci, 14, 43126, Parma, Emilia-Romagna, ITALY | 24-Oct-2016 |
| 291479 | TONINI, GIUSEPPE | Università Campus Bio-Medico di Roma, Via Alvaro del Portillo 200, 00128, Roma, Lazio, ITALY | 14-Sep-2016 |
| 291481 | MAIELLO, EVARISTO | COMITATO ETICO DELL'IRCCS GIOVANNI PAOLO II DI BARI PRESSO IRCCS CASA SOLLIEVO DELLA SOFFERENZA, V. le Cappuccini 1, 71013, San Giovanni Rotondo, Puglia, ITALY | 29-Sep-2016 |
| 291582 | DE MARINIS, FILIPPO | Comitato Etico Degli IRCCS Istituto Europeo di Oncologia e Centro Cardiologico Monzino, VIA RIPAMONTI 435, 20141, MILANO, Lombardia, ITALY | 21-Sep-2016 |
| 291583 | Chella, Antonio | Comitato Etico Regione Toscana - Area Vasta Nord Ovest , Via Roma 67, c/o Presidio Ospedaliero, 56126, Pisa, Toscana, ITALY | 14-Dec-2016 |
| 291672 | STUDNICKA, MICHAEL | Ethikkommission d. Landes Oberösterreich, Wagner-Jauregg-Weg 15, 4020, Linz, AUSTRIA | 27-Jul-2016 |
| 291681 | BURGHUBER, OTTO | Ethikkommission d. Landes Oberösterreich, Wagner-Jauregg-Weg 15, 4020, Linz, AUSTRIA | 27-Jul-2016 |
| 291745 | Hartl, Sylvia | Ethikkommission d. Landes Oberösterreich, Wagner-Jauregg-Weg 15, 4020, Linz, AUSTRIA | 27-Jul-2016 |
| 291771 | Thomas, Christian | Copernicus Group IRB, 5000 CentreGreen Way, Suite 200, CARY, NC, 27513, UNITED STATES | 08-Mar-2016 |

| Site # | Investigator | EC/IRB Name and Address | IRB/EC Approval |
|---|---|---|---|
| 291773 | Kirshner, Eli | Lehigh Valley Health Network IRB, 1019 39th Avenue SE, Suite 120, Puyallup, WA, 98374-2115, UNITED STATES | 26-May-2016 |
| 291776 | Veatch, Andrea | Copernicus Group IRB, 5000 CentreGreen Way, Suite 200, CARY, NC, 27513, UNITED STATES | 15-Mar-2016 |
| 291780 | Leal, Ticiana | Lehigh Valley Health Network IRB, 1019 39th Avenue SE, Suite 120, Puyallup, WA, 98374-2115, UNITED STATES | 04-Nov-2016 |
| 291781 | MacVicar, Gary | Copernicus Group IRB, One Triangle Drive, Suite 100, Research Triangle Park, NC, 27709, UNITED STATES | 31-Mar-2016 |
| 291820 | Mekhail, Tarek | Florida Hospital IRB, 901 N. Lake Destiny Rd, Suite 400, Maitland, FL, 32751, UNITED STATES | 11-Apr-2016 |
| 291820 | Mekhail, Tarek | WIRB-Western Institutinal Review Board, 1019 39th Avenue SE, Suite 120, Puyallup, WA, 98374, UNITED STATES | 11-Apr-2016 |
| 291821 | McCune, Steven L. | Copernicus Group IRB, 5000 CentreGreen Way, Suite 200, CARY, NC, 27513, UNITED STATES | 10-Jun-2016 |
| 291834 | DE BOER, RICHARD | Melbourne Health Human Research Ethics Committee, Flemington Rd, Office for Research Level 6 East, Main Building, 3050, Victoria, Victoria, AUSTRALIA | 04-Jul-2016 |
| 291837 | Kao, Steven | Concord Repatriation General Hospital HREC, Ground Floor - Building 20, Hospital Road, 2139, Concord, New South Wales, AUSTRALIA | 27-Jul-2016 |
| 291868 | Goldschmidt-Jr, Jerome | Copernicus Group IRB, One Triangle Drive, Suite 100, Research Triangle Park, NC, 27709, UNITED STATES | 10-Mar-2016 |
| 291881 | Hamm, John | Lehigh Valley Health Network IRB, 1019 39th Avenue SE, Puyallup, WA, 98374, UNITED STATES | 26-Jul-2016 |
| 291882 | Horn, Leora | Vanderbilt University Institutional Review Board, 1313 21st Ave. South, 504 Oxford House, Nashville, TN, 37232-4315, UNITED STATES | 24-May-2016 |
| 291894 | Sanchez, Amparo | Hospital Ramon y Cajal ;Comité Etico de Investigación Clínica, Ctra. Colmenar Viejo, km 9,1, 28034, Madrid, MADRID, SPAIN | 29-Jun-2016 |
| 291977 | Gershenhorn, Bruce | Lehigh Valley Health Network IRB, 1019 39th Avenue SE, Suite 120, Puyallup, WA, 98374-2115, UNITED STATES | 21-Sep-2016 |
| 291990 | Spira, Alexander I. | Copernicus Group IRB, One Triangle Drive, Suite 100, Research Triangle Park, NC, 27709, UNITED STATES | 23-May-2016 |
| 291998 | Batus, Marta | Rush University Medical Center; Rush University Research and Clinical Trials Administration Office, 1653 West Congress Parkway, Chicago, IL, 60612-3833, UNITED STATES | 28-Jul-2016 |
| 292005 | Braiteh, Fadi | Western Institutional Review Board, 3535 Seventh Avenue SW, Olympia, WA, 98502, UNITED STATES | 06-Jun-2016 |
| 292009 | Induru, Raghava | Chesapeake Research Review; IRB, 7063 Columbia Gateway Drive, Suite 110, Columbia, MD, 21046, UNITED STATES | 18-Aug-2016 |
| 292010 | Harris, Ronald | Copernicus Group IRB, 5000 CentreGreen Way, Suite 200, CARY, NC, 27513, UNITED STATES | 08-Mar-2016 |
| 292084 | SZCZESNA, ALEKSANDRA | Niezalezna Komisja Bioetyczna ds. Badan Naukowych przy GUMed, ul. M. Sklodowskiej-Curie 3a, 80-210, Gdansk, POLAND | 12-May-2016 |
| 292085 | Kazarnowicz, Andrzej | Niezalezna Komisja Bioetyczna ds. Badan Naukowych przy GUMed, ul. M. Sklodowskiej-Curie 3a, 80-210, Gdansk, POLAND | 12-May-2016 |
| 292086 | Dziadziuszko, Rafal | Niezalezna Komisja Bioetyczna ds. Badan Naukowych przy GUMed, ul. M. Sklodowskiej-Curie 3a, 80-210, Gdansk, POLAND | 12-May-2016 |
| 292087 | Symonowicz, Igor | Niezalezna Komisja Bioetyczna ds. Badan Naukowych przy GUMed, ul. M. Sklodowskiej-Curie 3a, 80-210, Gdansk, POLAND | 12-May-2016 |
| 292088 | Bryl, Maciej | Niezalezna Komisja Bioetyczna ds. Badan Naukowych przy GUMed, ul. M. Sklodowskiej-Curie 3a, 80-210, Gdansk, POLAND | 12-May-2016 |

| Site # | Investigator | EC/IRB Name and Address | IRB/EC Approval |
|---|---|---|---|
| 292093 | MARK, ZSUZSANNA | Gyogyszereszeti es Egeszsegugyi Minoseg- es Szervezetfejlesztesi Intezet; OGYI, Zrinyi u. 3, H-1051, Budapest, HUNGARY | 19-Jul-2016 |
| 292094 | OSTOROS, GYULA | Gyogyszereszeti es Egeszsegugyi Minoseg- es Szervezetfejlesztesi Intezet; OGYI, Zrinyi u. 3, H-1051, Budapest, HUNGARY | 19-Jul-2016 |
| 292096 | Havel, Libor | Etická komise pri IKEM a TN, Vídeňská 800, 14059, Prague, CZECH REPUBLIC | 31-Jul-2016 |
| 292172 | Hussein, Maen | Western Institutional Review Board, 1019 39th Avenue SE, Ste 120, Puyallup, WA, 98374, UNITED STATES | 25-Sep-2016 |
| 292175 | Mansfield, Aaron | Mayo Clinic Institutional Rev Bd Rochester, 200 First Street SW, 201 Building, Room 4-60, Rochester, MN, 55905, UNITED STATES | 23-Sep-2016 |
| 292178 | Califano, Raffaele | NRES Committee East Midlands - Leicester, The Old Chapel, Royal Standard Place,, Nottingham, NG1 6FS, UNITED KINGDOM | 24-Oct-2016 |
| 292179 | Conibear, John | NRES Committee East Midlands - Leicester, The Old Chapel, Royal Standard Place,, Nottingham, NG1 6FS, UNITED KINGDOM | 05-Jul-2016 |
| 292181 | Percent, Ivor J. | Western Institutional Review Board, 1019 39th Avenue SE, Ste 120, Puyallup, WA, 98374, UNITED STATES | 25-Sep-2016 |
| 292182 | Johnson, Melissa | Lehigh Valley Health Network IRB, 1019 39th Avenue SE, Puyallup, WA, 98374, UNITED STATES | 21-Sep-2016 |
| 292188 | Daniel, Davey | Lehigh Valley Health Network IRB, 1019 39th Avenue SE, Puyallup, WA, 98374, UNITED STATES | 25-Sep-2016 |
| 292221 | Karapanagiotou, Eleni | NRES Committee East Midlands - Leicester, The Old Chapel, Royal Standard Place,, Nottingham, NG1 6FS, UNITED KINGDOM | 05-Jul-2016 |
| 292225 | ANDRIC, ZORAN | Clinical Hospital Center Bezanijska kosa; Ethics Committee Clinical Hospital Center Bezanijska kosa, Bezanijska kosa bb, 11000, Belgrade, SERBIA | 07-Apr-2016 |
| 292226 | Rancic, Milan | Ethics Committee Clinical Center Nis, Bulevar Dr Zorana Dindica 48, 18-000, Nis, SERBIA | 05-Apr-2016 |
| 292227 | KARASEVA, NINA | EC at the St. Petersburg City Clinical Oncol. Disp., PROSPEKT VETERANOV, 56, 198255, ST PETERSBURG, RUSSIAN FEDERATION | 05-Jul-2016 |
| 292228 | SMOLIN, ALEXEY | Ethics Committee of the Main Military Clinical Hospital n.a. N.N.Burdenko, 3 Gospitalnaya square, 105229, Moscow, RUSSIAN FEDERATION | 27-Jul-2016 |
| 292229 | Stroyakovskii, Daniil | CITY CLINICAL ONCOLOGY HOSPITAL; Onco, KRASNOGORSKI DISTRICT, p/o Stepanovskoe, 143423, MOSCOW, RUSSIAN FEDERATION | 29-Jun-2016 |
| 292230 | GORBUNOVA, VERA | Blokhin Russian Cancer Research Center Ethics Committee , Kashirskoye shosse,24, Moscow, RUSSIAN FEDERATION | 14-Jun-2016 |
| 292231 | Levchenko, Evgeny | FSBI Research Oncology Institute n.a. N.N.Petrov of Ministry of Health of Russian Federation; onco, Persochny , Leningradskaya Str., bld. 68, 197758, Saint-Petersburg, RUSSIAN FEDERATION | 12-Jul-2016 |
| 292232 | Filippov, Alexander | E C at City Clinical Hospital No 1, Ulitsa Zalesskogo 6, 630047, Novosibirsk, RUSSIAN FEDERATION | 27-Jul-2016 |
| 292235 | SZILASI, MARIA | Gyogyszereszeti es Egeszsegugyi Minoseg- es Szervezetfejlesztesi Intezet; OGYI, Zrinyi u. 3, H-1051, Budapest, HUNGARY | 19-Jul-2016 |
| 292236 | Losonczy, Gyorgy | Gyogyszereszeti es Egeszsegugyi Minoseg- es Szervezetfejlesztesi Intezet; OGYI, Zrinyi u. 3, H-1051, Budapest, HUNGARY | 19-Jul-2016 |

| Site # | Investigator | EC/IRB Name and Address | IRB/EC Approval |
|---|---|---|---|
| 292237 | MELICHAR, BOHUSLAV | Eticka Komise Fakultni Nemocnice Olomouc, I.P. PAVLOVA 185/6, 779 00, Olomouc, CZECH REPUBLIC | 11-Jul-2016 |
| 292242 | Opalka, Petr | Eticka komise Nemocnice Na Bulovce, Budinova 2, 180 81, Praha 8, CZECH REPUBLIC | 02-Aug-2016 |
| 292258 | KRZAKOWSKI, MACIEJ | Niezalezna Komisja Bioetyczna ds. Badan Naukowych przy GUMed, ul. M. Sklodowskiej-Curie 3a, 80-210, Gdansk, POLAND | 12-May-2016 |
| 292327 | Popevic, Spasoje | Ethics Committee Clinical Center Of Serbia, PASTEROVA 2, 11000, BELGRADE, SERBIA | 19-May-2016 |
| 292335 | Castro Junior, Gilberto | CEP para Analise de Projetos de Pesquisa do HCFMUSP e da FMUSP;Hospital da Universidade de São Paulo, Rua Doutor Arnaldo, 455 - 01246-903, 05403-010, São Paulo, SP, BRAZIL | 08-Nov-2016 |
| 292337 | Pereira, Rodrigo | Comite de Etica do Hospital de Clinicas de Porto Alegre, Rua Ramiro Barcelos, 2350, 90035-903, Porto Alegre, RS, BRAZIL | 30-Aug-2016 |
| 292341 | Andrade, Livia | Hospital Santa Izabel - Santa Casa de Misericordia da Bahia, Praça Almeida Couto 500, 40050-410, Salvador, BA, BRAZIL | 01-Nov-2016 |
| 292354 | Reck, Martin | Ethikkommission der Ärztekammer Schleswig-Holstein, Bismarckallee 8-12, 23795, Bad Segeberg, GERMANY | 24-Nov-2016 |
| 292462 | YU, CHONG-JEN | Research Ethics Committee, Nat. Taiwan Univ. Hosp., 7 CHUNG-SHAN SOUTH ROAD , 100, TAIPEI, TAIWAN | 04-Jul-2016 |
| 292463 | Chiu, Chao-Hua | TVGH Institutional Review Board, No.201, Shih-Pai Road, Sec.2, 112, Taipei, TAIWAN | 04-May-2016 |
| 292464 | Yang, Cheng-Ta | Chang Gung Med Found, Institutional Review Board, No. 123, Dunghu Rd., Jioulu Village, Taoyuan County, 333, Gueishan Township, TAIWAN | 08-Aug-2016 |
| 292511 | Aren, Osvaldo | Comite de Etica Servicio de Salud Metropolitano Norte, Maruri 272, Independencia, 8380656, Santiago, CHILE | 20-Jun-2016 |
| 292544 | ORLANDI, FRANSCICO | Comite de Etica Servicio de Salud Metropolitano Oriente, Av. Salvador 364, Providencia, 7500922, Santiago, CHILE | 16-Aug-2016 |
| 292612 | LEE, JONG-SEOK | Seoul National University Bundang Hospital IRB, 82, Gumi-Ro 173 Beon-Gil, Bundang-Gu, 463-707, Seongnam-Si, Gyeonggi-Do, KOREA, REPUBLIC OF | 21-Jun-2016 |
| 292674 | Schütte, Wolfgang | EK des Landes Sachsen-Anhalt, Kühnauer Str. 70, 06846, Dessau-Roßlau, GERMANY | 24-Nov-2016 |
| 292728 | Bischoff, Helge | Ethikkommission der Medizinischen Fakultät Heidelberg, Alte Glockengießerei 11/1, 69115, Heidelberg, GERMANY | 24-Nov-2016 |
| 292731 | Reinmuth, Niels | Ethik-Kommission der Bayerischen Landesärztekammer, Mühlbaurstr. 16, Sekretariat, 81677, München, GERMANY | 24-Nov-2016 |
| 292732 | Rittmeyer, Achim | EK Hessen LÄK, Im Vogelsang 3, 60488, Frankfurt, GERMANY | 24-Nov-2016 |
| 293046 | GERVAIS, RADJ | CPP Nord Ouest IV | 05-Jul-2016 |
| 293047 | Greillier, Laurent | CPP Nord Ouest IV | 05-Jul-2016 |
| 293054 | SCHERPEREEL, ARNAUD | CPP Nord Ouest IV | 05-Jul-2016 |
| 293055 | Cousin, Sophie | CPP Nord Ouest IV, CHU de Lille - Service de Pharmacologie, 1 Place de Verdun, 59045, LILLE, FRANCE | 05-Jul-2016 |
| 293097 | Toy, Elizabeth | NRES Committee East Midlands - Leicester, The Old Chapel, Royal Standard Place,, Nottingham, NG1 6FS, UNITED KINGDOM | 05-Jul-2016 |
| 293395 | HUGHES, BRETT | Melbourne Health Human Research Ethics Committee, Grattan Street, 3050, PARKVILLE, Victoria, AUSTRALIA | 19-Jul-2016 |
| 294091 | TAKAHASHI, TOSHIAKI | Shizuoka Cancer Center Ethical Review Board for Business Clinical Studies, 1007 Shimonagakubo Nagaizumi-cho, Suntou-gun, 411-8777, Shizuoka, JAPAN | 21-Sep-2016 |
| 294092 | Yokoyama, Toshihide | Kurashiki Central Hospital Institutional Review Board, 1-1-1 Miwa, Kurashiki-shi, 710-8602, Okayama, JAPAN | 21-Sep-2016 |

| Site # | Investigator | EC/IRB Name and Address | IRB/EC Approval |
|---|---|---|---|
| 294093 | Hayashi, Hidetoshi | Kindai University Hospital Institutional Review Board, 377-2 Ohnohigashi, Osaka-Sayama-shi, 589-8511, Osaka, JAPAN | 26-Jul-2016 |
| 294094 | Sugawara, Shunichi | Sendai Kousei Hospital Institutional Review Board, 4-15 Hirose-Machi, Aoba-Ku, Sendai-shi, 980-0873, Miyagi, JAPAN | 20-Jul-2016 |
| 294095 | Akamatsu, Hiroaki | Wakayama Medical University Institutional Review Board, 811-1 Kimiidera, Wakayama-shi, 641-8510, Wakayama, JAPAN | 01-Aug-2016 |
| 294096 | Nishio, Makoto | The Cancer Institute Hospital of JFCR Institutional Review Board, 3-8-31 Ariake Koto-Ku, 135-8550, Tokyo, JAPAN | 03-Aug-2016 |
| 294097 | Sakai, Hiroshi | Saitama Cancer Center Institutional Review Board, 780 Komuro Inamachi, Kitaadachi-gun, 362-0806, Saitama, JAPAN | 26-Jul-2016 |
| 294098 | Kato, Terufumi | Kanagawa Cancer Center IRB, 1-1-2, Nakao, Asahi-ku, Yokohama-shi, 241-8515, Kanagawa, JAPAN | 28-Jul-2016 |
| 294099 | Okamoto, Isamu | Kyushu University Hospital IRB, 3-1-1 Maidashi, Higashi-Ku, Fukuoka-Shi, 812-8582, Fukuoka, JAPAN | 28-Jul-2016 |
| 294100 | Atagi, Shinji | National Hospital Organization Kinki-chuo Chest Medical Center Institutional Review Board, 1180 Nagasone-cho, Kita-ku, Sakai, 591-8555, Osaka, JAPAN | 22-Jul-2016 |
| 294101 | Okuma, Yusuke | Tokyo Metropolitan Komagome Hospital Ethics Committee, 3-18-22 HONKOMAGOME, BUNKYO-KU, 113-8677, TOKYO, JAPAN | 30-Sep-2016 |
| 294103 | Nakahara, Yasuharu | Institutional Review Board of National Hospital Organization Himeji Medical Center, 68 Honmachi, Himeji-shi, 670-8520, Hyogo, JAPAN | 04-Aug-2016 |
| 294199 | Takayama, Koichi | University Hospital, Kyoto Prefectural University of Medicine Institutional Review Board, 465 Kajii-cho, Kawaramachi-dori Hirokoji Agaru, Kamigyo-ku, 602-8566, Kyoto, JAPAN | 07-Sep-2016 |
| 295424 | Alatorre Alexander, Jorge Arturo | CEI de Clinica Bajio CLINBA; Comite de Etica en Investigacion, Valenciana 7, Col. Paxtitlán. Guanajuato, Gto., 36090, Guanajuato, MEXICO | 23-Aug-2016 |
| 297156 | Cheng, Ying | Ethic Committee of Jilin Cancer Hospital, 1018 HUGUANG ROAD, CHAOYANG DISTRICT, 130012, CHANGCHUN, CHINA | 18-Oct-2016 |
| 297159 | Hu, Jie | EC of Zhongshan Hospital Fudan University, 180 FENGLIN ROAD, 1474 WEST YANAN ROAD, 200032, SHANGHAI, CHINA | 20-Feb-2017 |
| 297160 | Chang, Jianhua | EC of Fudan University Shanghai Cancer Center, 5th Floor, Building 2, No.270, Dong'an Road, 200032, Shanghai, CHINA | 20-Jan-2017 |
| 297161 | He, Jianxing | EC Of The First Affiliated Hospital Of Guangzhou Medical University, No.151,Yanjiang Road, 510120, Guangzhou, CHINA | 28-Feb-2017 |
| 297217 | zhao, yanqiu | Henan Tumor Hospital; Ethics committee of Henan Tumor Hospital, 127# Dongming Rd, Jinshui District, 450008, Zhengzhou, CHINA | 13-Mar-2017 |
| 297706 | wang, ziping | The Ethics Committee of Beijing Cancer Hospital, No.52 Fucheng Road,, Haidian District,, 100036, Beijing, CHINA | 27-Dec-2016 |
| 297707 | Chen, Gongyan | Harbin Medical University Cancer Hospital; Ethics Committee, No.150, Haping Road, Nangang District, 150081, Harbin, CHINA | 11-Jan-2017 |
| 297780 | Fan, Yun | Zhejiang Cancer Hospital; Ethics Committee/IRB, second floor, building of administration, No.38, Guangji Road, 310022, Hangzhou City, CHINA | 23-Feb-2017 |
| 301344 | Rao, Suman | Chesapeake Research Review; IRB, 7063 Columbia Gateway Drive, Suite 110, Columbia, MD, 21046, UNITED STATES | 08-Mar-2017 |
| 303074 | FENG, JIFENG | EC of Jiangsu Cancer Hospital, No.42, Baiziting, 210009, Nanjing, CHINA | 29-Jun-2017 |
