## Supplementary material for "Allelic Variation in *HLA-DRB1* is Associated with Development of Anti-Drug Antibodies in Cancer Patients Treated with Atezolizumab that are Neutralizing *in Vitro*": Ethics Committee approvals: IMpower150 EC IRB.pdf

### Names and Addresses of Institutional Review Boards / Ethics Committees
### Protocol: GO29436

| Site # | Investigator | IRB/EC Name and Address | Approval Date |
|---|---|---|---|
| 278207 | Telivala, Bijoy | Copernicus Group Independent Review Board; 1 Triangle Drive Suite 100 PO Box 110605 Research Triangle Park North Carolina 27709 United States | 8-Apr-15 |
| 278208 | Hamm, John | Western Institutional Review Board; 1019 39th Avenue Southeast Suite 120 Puyallup Washington 98374 United States | 3-Jun-15 |
| 278209 | Shtivelband, Mikhael | Copernicus Group Independent Review Board; 1 Triangle Drive Suite 100 PO Box 110605 Research Triangle Park North Carolina 27709 United States | 8-Jun-15 |
| 278210 | Erickson, Brian | Copernicus Group Independent Review Board; 1 Triangle Drive Suite 100 PO Box 110605 Research Triangle Park North Carolina 27709 United States | 25-Feb-15 |
| 278223 | John, Thomas | Melbourne Health Human Research Ethics Committee | 4-Dec-15 |
| 278225 | Hughes, Brett | Melbourne Health Human Research Ethics Committee | 21-Apr-15 |
| 278265 | Crombie, Catherine | Melbourne Health Human Research Ethics Committee | 21-Apr-15 |
| 278283 | Hooberman, Arthur | Copernicus Group Independent Review Board; 1 Triangle Drive Suite 100 PO Box 110605 Research Triangle Park North Carolina 27709 United States | 4-Mar-15 |
| 278738 | Blinman, Prunella | Melbourne Health Human Research Ethics Committee | 21-Apr-15 |
| 278740 | Gauden, Stan | Tasmania Health and Medical Human Research Ethics Committee | 30-Mar-15 |
| 278746 | Potasz, Nicole | Melbourne Health Human Research Ethics Committee | 21-Apr-15 |
| 278748 | Singhal, Nimit | Melbourne Health Human Research Ethics Committee | 21-Apr-15 |
| 278754 | O'Byrne, Kenneth | Melbourne Health Human Research Ethics Committee | 21-Apr-15 |
| 278824 | Aleman Polanco, Diana Sofía | Asociacion Benefica Prisma | 23-Apr-15 |
| 278825 | Mas Lopez, Luis | Comité Institucional de ética en Investigación Instituto Nacional de Enfermedades Neoplasicas | 4-May-15 |
| 278827 | Lim, Darren | Singhealth Centralised Institutional Review Board | 10-Mar-15 |
| 278837 | Hung, Jen-Yu | Institutional Review Board Kaohsiung Medical University Chung-Ho Memorial Hospital | 30-May-15 |
| 278841 | Chen, Yuh-Min | Institutional Review Board Taipei Veterans General Hospital | 30-Mar-16 |
| 278844 | Li, Chien-Te | Institutional Review Board, Changhua Christian Hospital | 9-Jun-15 |
| 278847 | Chen, Chao-Hsun | Institutional Review Board of the Chi Mei Medical Center | 2-Jun-15 |
| 278849 | Chang, Gee-Chen | The Institutional Review Board of Taichung Veterans General Hospital | 8-Jun-15 |
| 278850 | Ho, Ching-Liang | Institutional Review Board of Tri-Service General Hospital | 13-Jun-15 |
| 279286 | Chen, Wei-Teing | Cheng-Hsin General Hospital Institutional Review Board | 9-Jul-15 |
| 279291 | Magri, Ignacio | Consejo de Evaluación Ética de Investigación en Salud - CoEIS | 14-Jul-16 |
| 279293 | Kowalyszyn, Rubén | Ministerio de Salud - Provincia de Río Negro | 12-Nov-15 |
| 279293 | Kowalyszyn, Rubén | Comité Independiente de Etica en investigación clínica "Dr. Carlos A. Barclay | 12-Nov-15 |
| 279295 | Lerzo, Guillermo | Comité Independiente de Etica en investigación clínica "Dr. Carlos A. Barclay | 12-Feb-15 |
| 279300 | Jarchum, Gustavo | Consejo de Evaluación Ética de Investigación en Salud - CoEIS | 1-Dec-15 |
| 279302 | Kaen, Diego | Comité Independiente de Etica en investigación clínica "Dr. Carlos A. Barclay | 19-Feb-15 |
| 279305 | Kahl, Susana | Comité de ética en investigación Fundación Oncosalud | 20-Feb-16 |
| 279305 | Kahl, Susana | Comisión Conjunta de Investigación en Salud (CCIS) | 20-Feb-16 |

| Site # | Investigator | IRB/EC Name and Address | Approval Date |
|---|---|---|---|
| 279308 | Varela, Mirta | Comité de Ética Centro de Oncología e Investigación Buenos Aires | 6-May-15 |
| 279308 | Varela, Mirta | Comisión Conjunta de Investigación en Salud (CCIS) | 6-May-15 |
| 279311 | Kotliar, Mauricio | Comité de Ética Independiente - Fundación Sanatorio | 28-Apr-15 |
| 279317 | Lin, Yu-Ching | Institutional Review Board of Chang Gung Medical Foundation | 11-Jun-15 |
| 279318 | Eckmayr, Josef | Ethikkommission für das Bundesland Salzburg; Sebastian-Stief-Gasse 2 Salzburg Salzburg 5010 Austria | 16-Oct-15 |
| 279322 | Zvirbule, Zanete | The Ethics Committee for Clinical Trials of Medicinal Products | 24-Apr-15 |
| 279787 | Drew, David | Copernicus Group Independent Review Board; 1 Triangle Drive Suite 100 PO Box 110605 Research Triangle Park North Carolina 27709 United States | 1-Apr-15 |
| 279788 | Almubarak, Mohammed | Chesapeake Institutional Review Board; 6940 Columbia Gateway Drive Suite 110 Columbia Maryland 21046 United States | 13-ago-15 |
| 279789 | Assikis, Vasileios | Copernicus Group Independent Review Board; 1 Triangle Drive Suite 100 PO Box 110605 Research Triangle Park North Carolina 27709 United States | 9-Apr-15 |
| 279819 | Belman, Neil | St. Luke’s Hospital & Health Network IRB; 801 Ostrum Street East Wing 4 Bethlehem Pennsylvania 18015 United States | 25-Mar-15 |
| 279820 | Chaudhry, Arvind | Copernicus Group Independent Review Board; 1 Triangle Drive Suite 100 PO Box 110605 Research Triangle Park North Carolina 27709 United States | 3-Feb-15 |
| 279821 | Chitneni, Shobha | Copernicus Group Independent Review Board; 1 Triangle Drive Suite 100 PO Box 110605 Research Triangle Park North Carolina 27709 United States | 29-Jan-15 |
| 279822 | Cohenuram, Michael | Copernicus Group Independent Review Board; 1 Triangle Drive Suite 100 PO Box 110605 Research Triangle Park North Carolina 27709 United States | 13-Mar-15 |
| 279823 | De Vore, Russell | Copernicus Group Independent Review Board; 1 Triangle Drive Suite 100 PO Box 110605 Research Triangle Park North Carolina 27709 United States | 3-Jun-15 |
| 279824 | Goldschmidt, Jerome | Copernicus Group Independent Review Board; 1 Triangle Drive Suite 100 PO Box 110605 Research Triangle Park North Carolina 27709 United States | 4-Feb-15 |
| 279825 | Halibey, Bohdan | Copernicus Group Independent Review Board; 1 Triangle Drive Suite 100 PO Box 110605 Research Triangle Park North Carolina 27709 United States | 10-Apr-15 |
| 279826 | Kirshner, Eli | Western Institutional Review Board; 1019 39th Avenue Southeast Suite 120 Puyallup Washington 98374 United States | 31-mar-15 |
| 279827 | Martin, William | St Charles Medical Center; 2500 Northeast Neff Road Bend Oregon 97701 United States | 25-mar-15 |
| 279828 | Mitchell, Reed | Copernicus Group Independent Review Board; 1 Triangle Drive Suite 100 PO Box 110605 Research Triangle Park North Carolina 27709 United States | 26-Jan-15 |
| 279829 | Subramanian, Janakiraman | Saint Luke's Hospital Institutional Review Board; 915 East First Street Duluth Minnesota 55805 United States | 8-Jun-15 |
| 279830 | Swanson, Paul | Copernicus Group Independent Review Board; 1 Triangle Drive Suite 100 PO Box 110605 Research Triangle Park North Carolina 27709 United States | 20-Feb-15 |
| 279831 | Tsai, Frank Yung-Chin | Western Institutional Review Board; 1019 39th Avenue Southeast Suite 120 Puyallup Washington 98374 United States | 8-Jul-15 |
| 279832 | Abdel Karim, Nagla | Western Institutional Review Board; 1019 39th Avenue Southeast Suite 120 Puyallup Washington 98374 United States | 19-feb-16 |
| 279833 | Früh, Martin | Kantonale Ethikkommission Bern (KEK) | 3-Mar-16 |
| 279836 | Orlandi Jorquera, Francisco | Comite de Etica Cientifico del Servicio de Salud Metropolitano Norte | 26-May-15 |
| 279838 | Allegrini, Giacomo | Comitato Etico Area Vasta Nord Ovest presso Azienda Ospedaliero Universitaria Pisana di Pisa | 9-Jul-15 |

| Site # | Investigator | IRB/EC Name and Address | Approval Date |
|---|---|---|---|
| 279839 | Cartenì, Giacomo | Comitato Etico Cardarelli-Santobono | 7-Oct-15 |
| 279840 | Falcone, Alfredo | Comitato Etico Area Vasta Nord Ovest presso Azienda Ospedaliero Universitaria Pisana di Pisa | 9-Jul-15 |
| 279841 | Mencoboni, Manlio | Comitato Etico San Martino IST 2 | 16-Jul-15 |
| 279843 | Migliorino, Maria | Comitato Etico Lazio 1 | 30-Jul-15 |
| 279844 | Tonini, Giuseppe | COMITATO ETICO DELL´UNIVERSITA´ CAMPUS BIO-MEDICO DI ROMA | 18-Jun-15 |
| 279845 | Amoroso, Domenico | Comitato Etico Area Vasta Nord Ovest presso Azienda Ospedaliero Universitaria Pisana di Pisa | 9-Jul-15 |
| 279846 | Biesma, Bonne | Medical Research Ethics Committees United | 29-Oct-15 |
| 279847 | Wilschut, Frank | Medical Research Ethics Committees United | 29-Oct-15 |
| 279848 | Moro-Sibilot, Denis | CPP Sud-Méditerranée 2; 270 Boulevard Sainte Marguerite Hôpital Sainte Margeurite Pavillon 9 Cedex 9 Marseille Bouches-du-Rhône 13274 France | 5-Jun-15 |
| 279849 | Kosmider, Suzanne | Melbourne Health Human Research Ethics Committee | 21-Apr-15 |
| 279850 | Millward, Michael | Sir Charles Gairdner Hospital HREC | 28-Jul-15 |
| 279851 | Parnis, Francis | Bellberry Human Research Ethics Committee | 5-May-15 |
| 279852 | Lewis, Craig | Melbourne Health Human Research Ethics Committee | 21-Apr-15 |
| 280022 | Richardson, Gary | Cabrini Human Research Ethics Committee | 29-Jul-15 |
| 280023 | Gill, Sanjeev | Melbourne Health Human Research Ethics Committee | 20-May-15 |
| 280024 | Soto-Parra, Hector | Comitato Etico Catania 1 presso A.O. Universitaria Policlinico Vittorio Emanuele di Catania | 9-Jun-15 |
| 280029 | Almodovar, Maria Teresa | Comissão de Ética para a Investigação Clínica - CEIC | 10-Jul-15 |
| 280031 | Araújo, António | Comissão de Ética para a Investigação Clínica - CEIC | 10-Jul-15 |
| 280032 | Barata, Fernando | Comissão de Ética para a Investigação Clínica - CEIC | 10-Jul-15 |
| 280033 | Queiroga, Henrique | Comissão de Ética para a Investigação Clínica - CEIC | 10-Jul-15 |
| 280034 | Azevedo, Isabel | Comissão de Ética para a Investigação Clínica - CEIC | 10-Jul-15 |
| 280035 | Wesseler, Claas | Ethikkommission an der Universität Regensburg; Landshuter Straße 4 Raum 134, 1.OG. Regensburg 93047 Germany | 3-Nov-15 |
| 280037 | Wermke, Martin | Ethikkommission an der Universität Regensburg | 3-Nov-15 |
| 280038 | Wehler, Thomas | Ethikkommission an der Universität Regensburg; Landshuter Straße 4 Raum 134, 1.OG. Regensburg 93047 Germany | 3-Nov-15 |
| 280039 | Loges, Sonja | Ethikkommission an der Universität Regensburg | 3-Nov-15 |
| 280040 | Serke, Monika | Ethikkommission an der Universität Regensburg; Landshuter Straße 4 Raum 134, 1.OG. Regensburg 93047 Germany | 3-Nov-15 |
| 280043 | Kokowski, Konrad | Ethikkommission an der Universität Regensburg; Landshuter Straße 4 Raum 134, 1.OG. Regensburg 93047 Germany | 3-Nov-15 |
| 280044 | Schulz, Christian | Ethikkommission an der Universität Regensburg; Landshuter Straße 4 Raum 134, 1.OG. Regensburg 93047 Germany | 3-Nov-15 |
| 280046 | Fischer, Jürgen | Ethikkommission an der Universität Regensburg; Landshuter Straße 4 Raum 134, 1.OG. Regensburg 93047 Germany | 3-Nov-15 |
| 280048 | Behringer, Dirk | Ethikkommission an der Universität Regensburg; Landshuter Straße 4 Raum 134, 1.OG. Regensburg 93047 Germany | 3-Nov-15 |
| 280049 | Bargon, Joachim | Ethikkommission an der Universität Regensburg; Landshuter Straße 4 Raum 134, 1.OG. Regensburg 93047 Germany | 3-Nov-15 |
| 280052 | Rothenstein, Jeffrey | Lakeridge Health REB; 1 Hospital Court Oshawa Ontario L1G 2B9 Canada | 15-Jul-15 |

| Site # | Investigator | IRB/EC Name and Address | Approval Date |
|---|---|---|---|
| 280255 | Dobrescu, Andrei | Copernicus Group Independent Review Board; 1 Triangle Drive Suite 100 PO Box 110605 Research Triangle Park North Carolina 27709 United States | 8-May-15 |
| 280256 | Stella, Philip | St. Joseph Mercy Health System Institutional Review Board #2 - Oncology Central IRB; 5301 East Huron River Drive Clinical Research Department, RHB 6017 Ann Arbor Michigan 48106 United States | 16-jul-15 |
| 280258 | Alnsour, Mohammad | Mercy Saint Vincent Medical Center Institutional Review Board; 2213 Cherry Street Toledo Ohio 43608 United States | 13-Apr-15 |
| 280259 | Lukas, Jason | Western Institutional Review Board; 1019 39th Avenue Southeast Suite 120 Puyallup Washington 98374 United States | 3-Jun-15 |
| 280260 | Jotte, Robert | US Oncology Inc. Institutional Review Board; 10101 Woodloch Forest The Woodlands Texas 77380 United States | 18-Jun-15 |
| 280262 | Yanagihara, Ronald | Western Institutional Review Board (WIRB); 1019 39th Avenue Southeast Suite 120 Puyallup Washington 98374 United States | 26-Jan-16 |
| 280263 | Finley, Gene | Copernicus Group Independent Review Board; 1 Triangle Drive Suite 100 PO Box 110605 Research Triangle Park North Carolina 27709 United States | 16-Jun-15 |
| 280264 | Thomas, Christian | Copernicus Group Independent Review Board; 1 Triangle Drive Suite 100 PO Box 110605 Research Triangle Park North Carolina 27709 United States | 11-May-15 |
| 280265 | Silberberg, Jeffrey | Copernicus Group Independent Review Board; 1 Triangle Drive Suite 100 PO Box 110605 Research Triangle Park North Carolina 27709 United States | 8-May-15 |
| 280267 | Garcia, Yolanda | CEIC de la Corporacion Sanitaria del Parc Tauli | 9-Jun-15 |
| 280268 | Lopez Brea, Marta | CEIC de Cantabria | 9-Jun-15 |
| 280269 | Barneto-Aranda, Isidoro | CEIC de Andalucia (CCEIBA) | 9-Jun-15 |
| 280270 | De Castro Carpeño, Javier | CEIC Hospital Universitario La Paz | 9-Jun-15 |
| 280271 | Domine Gomez, Manuel | CEIC Fundación Jiménez Díaz | 9-Jun-15 |
| 280272 | Insa Molla, Amelia | CEIC Hospital Clínico Universitario de Valencia | 9-Jun-15 |
| 280273 | Alvarez, Rosa | CEIC Hospital General Universitario Gregorio Marañon | 9-Jun-15 |
| 280274 | Gonzalez Larriba, Jose Luis | CEIC Hospital Clinico San Carlos | 9-Jun-15 |
| 280275 | Jiménez Munarriz, Beatriz | CEIC Grupo Hospital de Madrid | 9-Jun-15 |
| 280300 | Taus, Alvaro | CEIC Parc de Salut Mar | 9-Jun-15 |
| 280301 | Terrasa Pons, Josefa | CEIC Islas Baleares (CEIC-IB) | 9-Jun-15 |
| 280302 | Vazquez Estevez, Sergio | CEIC de Galicia (CAEI) | 9-Jun-15 |
| 280303 | Viñolas, Nuria | CEIC Hospital Clinic de Barcelona | 9-Jun-15 |
| 280506 | Ochsenbein, Adrian | Kantonale Ethikkommission Bern (KEK) | 3-Mar-16 |
| 280507 | Schütte, Wolfgang | Ethikkommission an der Universität Regensburg; Landshuter Straße 4 Raum 134, 1.OG. Regensburg 93047 Germany | 3-Nov-15 |
| 280508 | Stauder, Heribert | Ethikkommission an der Universität Regensburg; Landshuter Straße 4 Raum 134, 1.OG. Regensburg 93047 Germany | 3-Nov-15 |
| 280509 | Weißinger, Florian | Ethikkommission an der Universität Regensburg; Landshuter Straße 4 Raum 134, 1.OG. Regensburg 93047 Germany | 3-Nov-15 |
| 280510 | Engel-Riedel, Walburga | Ethikkommission an der Universität Regensburg; Landshuter Straße 4 Raum 134, 1.OG. Regensburg 93047 Germany | 3-Nov-15 |
| 280913 | Beatty, Patrick | Copernicus Group Independent Review Board; 1 Triangle Drive Suite 100 PO Box 110605 Research Triangle Park North Carolina 27709 United States | 17-Mar-15 |
| 280915 | Goueli, Basem | Saint Luke's Hospital Institutional Review Board; 915 East | 1-Sep-15 |

| Site # | Investigator | IRB/EC Name and Address | Approval Date |
|---|---|---|---|
| | | First Street Duluth Minnesota 55805 United States | |
| 280916 | Koh, Han | Kaiser Permanente Southern California Institutional Review Board; 393 East Walnut Street, 2nd Floor Pasadena California 91188 United States | 19-May-15 |
| 280917 | Schnell, Frederick | Copernicus Group Independent Review Board; 1 Triangle Drive Suite 100 PO Box 110605 Research Triangle Park North Carolina 27709 United States | 13-Jul-15 |
| 280919 | Nott, Louise | Tasmania Health and Medical Human Research Ethics Committee | 2-Jun-15 |
| 280920 | Yu, Chong-Jen | Research Ethics Committee of National Taiwan University Hospital | 16-Jun-15 |
| 280921 | Liu, Chien-Ying | Institutional Review Board of Chung Shan Medical University Hospital | 11-Jun-15 |
| 280922 | Greil, Richard | Ethikkommission für das Bundesland Salzburg; Sebastian-Stief-Gasse 2 Salzburg Salzburg 5010 Austria | 16-Oct-15 |
| 280925 | Reck, Martin | Ethikkommission an der Universität Regensburg; Landshuter Straße 4 Raum 134, 1.OG. Regensburg 93047 Germany | 3-Nov-15 |
| 280927 | Bourhaba, Maryam | CHU de Liège - Comité d'Ethique; Domaine Universitaire du Sart Tilman B35 Liege 4000 Belgium | 29-May-15 |
| 280928 | Goeminne, Jean-Charles | CHU de Liège - Comité d'Ethique; Domaine Universitaire du Sart Tilman B35 Liege 4000 Belgium | 29-May-15 |
| 280929 | Purkalne, Gunta | The Ethics Committee for Clinical Trials of Medicinal Products | 24-Apr-15 |
| 280930 | Lohri, Andreas | Kantonale Ethikkommission Bern (KEK) | 8-Mar-16 |
| 280932 | Hashemi, Sayed | Medical Research Ethics Committees United | 29-Oct-15 |
| 280983 | Masood, Nehal | Copernicus Group Independent Review Board; 1 Triangle Drive Suite 100 PO Box 110605 Research Triangle Park North Carolina 27709 United States | 4-Jun-15 |
| 280985 | Teixeira, Encarnação | Comissão de Ética para a Investigação Clínica - CEIC | 10-Jul-15 |
| 280986 | Adamchuk, Hryhoriy | CEQ of MI Kryvyi Rih Oncology Dispensary of Dnipropetrovsk Regional Council | 27-Feb-15 |
| 280987 | Andrusenko, Orest | CEQ of Treatment and Prevention Institution Volyn Regional Oncology Dispensary | 15-May-15 |
| 280988 | Bondarenko, Igor | CEQ of MI Dnipropetrovsk City Multifield Clinical Hospital #4 of Dnipropetrovsk Regional Council | 19-Mar-15 |
| 280989 | Chornobai, Anatolii | CEQ of Poltava Regional Clinical Oncology Dispensary of Poltava Regional Council | 26-Feb-15 |
| 280990 | Hotko, Yevhen | Commission on Ethics Questions of Uzhgorod Central City Clinical Hospital | 3-Mar-15 |
| 280991 | Ivashchuk, Oleksandr | Commission of Ethics Questions on the basis of the Chernivtsi Regional Clinical Oncology Dispensary | 8-Jul-15 |
| 280992 | Kolesnik, Oleksii | CEQ of MI of Zaporizhzhia Regional Council Zaporizhzhia Regional Clinical Oncology Dispensary | 11-Jun-15 |
| 280994 | Rusyn, Andriy | CEQ of Transcarpathian Regional Clinical Oncology Dispensary | 10-Mar-15 |
| 280995 | Vasylyev, Leonid | CEQ of SI Institute of Medical Radiology n.a. S.P. Hryhoriev of NAMS of Ukraine | 17-Mar-15 |
| 280996 | Shapovalov, Dmytro | CEQ of Municipal Noncommercial Institution Regional Center of Oncology | 20-Mar-15 |
| 280998 | Vynnychenko, Ihor | CEQ of Regional Municipal Institution Sumy Regional Clinical Oncology Dispensary | 2-Mar-15 |
| 280999 | Borra, Gloria | Comitato Etico Azienda Ospedaliera Universitaria Maggiore della Carità | 27-Jul-15 |
| 281001 | Barone, Carlo | Comitato Etico Dell Universita Cattolica del Sacro Cuore Policlinico Universitario Agostino Gemelli | 24-Sep-15 |
| 281002 | Girotto, Gustavo | Comitê de Ética em Pesquisa em Seres Humanos da Faculdade de Medicina de São José do Rio Preto | 11-May-15 |
| 281003 | Aragao, Bruno | Comitê de Ética em Pesquisa em Seres Humanos do Hospital Socor | 9-Mar-16 |
| 281005 | Faccio, Adilson | Comitê de Ética em Pesquisa em Seres Humanos da Universidade de Ribeirão Preto (UNAERP) | 13-Apr-16 |
| 281009 | Campos, Clodoaldo | Comitê de Ética em Pesquisa em Seres Humanos da | 8-May-16 |

| Site # | Investigator | IRB/EC Name and Address | Approval Date |
|---|---|---|---|
| | | Irmandade da Santa Casa de Londrina | |
| 281010 | Matias, Danielli | Comitê de Ética em Pesquisa em Seres Humanos da Liga Norte Riograndense Contra o Câncer | 2-Feb-16 |
| 281011 | da Silva, Carlos | Comitê de Ética em Pesquisa Fundação Pio XII Hospital de Câncer de Barretos | 18-Feb-16 |
| 281013 | Lopez, Yamil | Comité de Ética en Investigación de la Facultad de Medicina y Hospital Universitario | 12-Aug-15 |
| 281857 | Faller, Bryan | Missouri Baptist Medical Center Institutional Review Board; 3015 North Ballas Road St. Louis Missouri 63131 United States | 13-Jul-15 |
| 281858 | Paschold, John | US Oncology Inc. Institutional Review Board; 10101 Woodloch Forest The Woodlands Texas 77380 United States | 18-Jun-15 |
| 281860 | Ou, Sai-Hong Ignatius | University of California Irvine Institutional Review Board; 5171 Calfornia Avenue, Office Of Research Suite 150 Irvine California 92697 United States | 22-Jul-15 |
| 281904 | Carr, Laurie | Western Institutional Review Board; 1019 39th Avenue Southeast Suite 120 Puyallup Washington 98374 United States | 16-Oct-15 |
| 281905 | Daniels, Gregory | University of California, San Diego Human Research Protections Program; 3350 La Jolla Village Drive San Diego California 92161 United States | 18-Feb-16 |
| 281906 | Eskander, Elhamy | Frederick Memorial Hospital Institutional Review Board; 400 West 7th Street Frederick Maryland 21701 United States | 27-Jun-16 |
| 281907 | Kotiah, Sandy | Mercy Medical Center IRB; 345 St. Paul Place Bunting Center, 7th Floor Baltimore Maryland 21202 United states | 20-Apr-15 |
| 281908 | Kuzma, Charles | Copernicus Group Independent Review Board; 1 Triangle Drive Suite 100 PO Box 110605 Research Triangle Park North Carolina 27709 United States | 19-May-15 |
| 281909 | Hoffman, Philip | University of Chicago Hospitals Institutional Review Board; 5751 South Woodlawn Avenue McGiffert Hall Chicago Illinois 60637 United States | 7-Aug-15 |
| 281910 | Rodriguez, Estelamari | Mount Sinai Medical Center IRB; 4300 Alton Road Miami Beach Florida 33140 United States | 19-Oct-15 |
| 281944 | Pastor, Andrea | Comité Provincial de Bioética - Ministerio de Salud de la Provincia de Santa Fé | 28-Oct-15 |
| 281945 | Streich, Guillermo | Comité Independiente de Etica en investigación clínica "Dr. Carlos A. Barclay | 30-Jun-15 |
| 281946 | Aerts, Joachim | Medical Research Ethics Committees United | 29-Oct-15 |
| 281947 | Schramel, Franz | Medical Research Ethics Committees United | 29-Oct-15 |
| 281948 | Snijders, Dominic | Medical Research Ethics Committees United | 29-Oct-15 |
| 281950 | Aerts, Joachim | Medical Research Ethics Committees United | 29-Oct-15 |
| 281954 | Dickgreber, Nicolas | Ethikkommission an der Universität Regensburg; Landshuter Straße 4 Raum 134, 1.OG. Regensburg 93047 Germany | 3-Nov-15 |
| 281957 | Kollmeier, Jens | Ethikkommission an der Universität Regensburg; Landshuter Straße 4 Raum 134, 1.OG. Regensburg 93047 Germany | 3-Nov-15 |
| 281964 | Gomez-Villanueva, Angel | Comite de Etica Investigacion de la Clinica Bajío | 5-Jun-15 |
| 281966 | Dominguez Andrade, Adriana | Comite de Etica en Investigacion de Mexico Centre for Clinical Research SA de CV | 15-May-15 |
| 281966 | Dominguez Andrade, Adriana | Comite de Etica en Investigacion de Mexico Centre for Clinical Research SA de CV | 15-May-15 |
| 281983 | Costamilan, Rita de Cassia | Comitê de Ética em Pesquisa da Universidade de Caxias do Sul | 29-Mar-16 |
| 282032 | Schwartsmann, Gilberto | Comitê de Ética em Pesquisa do Hospital de Clínicas de Porto Alegre | 6-Apr-16 |
| 282033 | Tadokoro, Hakaru | Comite de Etica em Pesquisa da Universidade Federal de Sao Paulo - Hospital Sao Paulo | 17-Mar-16 |
| 282034 | Bruno, Luiz | Comitê de Ética em Pesquisa - Hospital Mãe de Deus | 10-Mar-16 |
| 282035 | Abbade Dettino, Aldo | Comitê de Ética em Pesquisa da Fundação Antônio Prudente – Hospital do Câncer A. C. Camargo | 6-May-16 |

| Site # | Investigator | IRB/EC Name and Address | Approval Date |
|---|---|---|---|
| 282036 | Dimitrov, Borislav | Ethics Committee for Multi-Centre Trials; 5 Sveta Nedelya Square Sofia Sofia-Grad 1000 Bulgaria | 3-Jun-15 |
| 282037 | Koynov, Krassimir | Ethics Committee for Multi-Centre Trials; 5 Sveta Nedelya Square Sofia Sofia-Grad 1000 Bulgaria | 3-Jun-15 |
| 282038 | Mihaylova, Zhasmina | Ethics Committee for Multi-Centre Trials; 5 Sveta Nedelya Square Sofia Sofia-Grad 1000 Bulgaria | 20-Jan-16 |
| 282039 | Ilieva, Rumyana | Ethics Committee for Multi-Centre Trials; 5 Sveta Nedelya Square Sofia Sofia-Grad 1000 Bulgaria | 3-Jun-15 |
| 282043 | Galiulin, Rinat | Ethics Committee at Clinical Oncology Dispensary | 22-Jun-15 |
| 282044 | Karaseva, Nina | Ethics Committee at City Clinical oncologic dispensary | 7-Jul-15 |
| 282047 | Gorbunova, Vera | Ethics Committee at Russian Oncology Research Center n.a. N.N.Blokhin | 30-Jun-15 |
| 282048 | Stroyakovskii, Daniil | Ethics Committee at Moscow City Oncology Hospital #62 of Moscow Healthcare Department | 26-Jul-15 |
| 282049 | Kovalenko, Nadezhda | Ethics Committee at Volzhskiy regional clinical oncology dispensary #3 | 27-Jun-16 |
| 282051 | Berard, Henri | CPP Sud-Méditerranée 2; 270 Boulevard Sainte Marguerite Hôpital Sainte Margeurite Pavillon 9 Cedex 9 Marseille Bouches-du-Rhône 13274 France | 11-Jun-15 |
| 282053 | Coudert, Bruno | CPP Sud-Méditerranée 2; 270 Boulevard Sainte Marguerite Hôpital Sainte Margeurite Pavillon 9 Cedex 9 Marseille Bouches-du-Rhône 13274 France | 5-Jun-15 |
| 282054 | Denis, Fabrice | CPP Sud-Méditerranée 2; 270 Boulevard Sainte Marguerite Hôpital Sainte Margeurite Pavillon 9 Cedex 9 Marseille Bouches-du-Rhône 13274 France | 5-Jun-15 |
| 282056 | Fabre, Elizabeth | CPP Sud-Méditerranée 2; 270 Boulevard Sainte Marguerite Hôpital Sainte Margeurite Pavillon 9 Cedex 9 Marseille Bouches-du-Rhône 13274 France | 5-Jun-15 |
| 282163 | Barlesi, Fabrice | CPP Sud-Méditerranée 2; 270 Boulevard Sainte Marguerite Hôpital Sainte Margeurite Pavillon 9 Cedex 9 Marseille Bouches-du-Rhône 13274 France | 5-Jun-15 |
| 282164 | Dayen, Charles | CPP Sud-Méditerranée 2; 270 Boulevard Sainte Marguerite Hôpital Sainte Margeurite Pavillon 9 Cedex 9 Marseille Bouches-du-Rhône 13274 France | 5-Jun-15 |
| 282165 | Foa, Cyril | CPP Sud-Méditerranée 2; 270 Boulevard Sainte Marguerite Hôpital Sainte Margeurite Pavillon 9 Cedex 9 Marseille Bouches-du-Rhône 13274 France | 5-Jun-15 |
| 282166 | Gazaille, Virgile | CPP Sud-Méditerranée 2; 270 Boulevard Sainte Marguerite Hôpital Sainte Margeurite Pavillon 9 Cedex 9 Marseille Bouches-du-Rhône 13274 France | 5-Jun-15 |
| 282167 | Veillon, Remi | CPP Sud-Méditerranée 2; 270 Boulevard Sainte Marguerite Hôpital Sainte Margeurite Pavillon 9 Cedex 9 Marseille Bouches-du-Rhône 13274 France | 5-Jun-15 |
| 282168 | Morere, Jean François | CPP Sud-Méditerranée 2; 270 Boulevard Sainte Marguerite Hôpital Sainte Margeurite Pavillon 9 Cedex 9 Marseille Bouches-du-Rhône 13274 France | 5-Jun-15 |
| 282169 | Audigier-Valette, Clarisse | CPP Sud-Méditerranée 2; 270 Boulevard Sainte Marguerite Hôpital Sainte Margeurite Pavillon 9 Cedex 9 Marseille Bouches-du-Rhône 13274 France | 5-Jun-15 |
| 282213 | Kryzhanivska, Anna | CEQ of Ivano-Frankivsk Regional Oncology Dispensary | 12-May-16 |
| 282442 | Rodriguez-Abreu, Delvys | CEIC Hospital Universitario Insular Materno-Infantil de Las Palmas | 9-Jun-15 |
| 282444 | Felip Font, Enriqueta | CEIC Hospital Universitari Vall d'Hebron | 9-Jun-15 |
| 282445 | Garrido Lopez, Pilar | CEIC Hospital Universitario Ramon y Cajal | 9-Jun-15 |
| 282447 | Ponce Aix, Santiago | CEIC Hospital Universitario 12 de Octubre | 9-Jun-15 |
| 282448 | Beniak, Juraj | Eticka komisia Presovskeho samospravneho kraja | 29-Jun-15 |
| 282449 | Kasan, Peter | Eticka komisia Univerzitna nemocnica Bratislava | 29-Jun-15 |
| 282452 | Cicenas, Saulius | Lithuanian Bioethics Committee | 20-May-15 |
| 282454 | Filipauskiene, Judita | Lithuanian Bioethics Committee | 20-May-15 |

| Site # | Investigator | IRB/EC Name and Address | Approval Date |
|---|---|---|---|
| 282455 | Nadal, Ernest | CEIC Hospital Universitari de Bellvitge | 9-Jun-15 |
| 282880 | Godal, Robert | Eticka komisia pri Narodnom onkologickom ustave | 29-Jun-15 |
| 282882 | Moron Escobar, Hernan | Asociacion Benefica Prisma | 28-May-15 |
| 282884 | Gurubhagavatula, Sarada | Copernicus Group Independent Review Board; 1 Triangle Drive Suite 100 PO Box 110605 Research Triangle Park North Carolina 27709 United States | 20-May-15 |
| 282885 | Nissenblatt, Michael | Copernicus Group Independent Review Board; 1 Triangle Drive Suite 100 PO Box 110605 Research Triangle Park North Carolina 27709 United States | 19-May-15 |
| 282886 | Herman, James | WIRB Copernicus Group; 1 Triangle Drive Suite 100 PO Box 110605 Research Triangle Park North Carolina 27709 United States | 29-Jan-16 |
| 282887 | Kono, Scott | Kaiser Permanente of Colorado Institutional Review Board; 10065 East Harvard Avenue Suite 300 Denver Colorado 80231 United States | 27-Jan-16 |
| 282970 | Zhiltsova, Elena | Ethics Committee at Russian Medical Military Academy n.a. S.M.Kirov | 30-Jun-15 |
| 283383 | Cisneros Tipismana, Rocio | Comite de Etica en Investigacion del Instituto Regional de Enfermedades Neoplasicas | 13-Nov-15 |
| 284076 | Su, Wu-Chou | National Cheng Kung University Hospital Human Experiment and Ethic Committee | 12-Jun-15 |
| 284111 | VanderWalde, Ari | Western Institutional Review Board (WIRB); 1019 39th Avenue Southeast Suite 120 Puyallup Washington 98374 United States | 16-Nov-15 |
| 284113 | Bernicker, Eric | Houston Methodist Research Institute IRB; 6670 Bertner Suite 6-351 Houston Texas 77030 United States | 15-Dec-15 |
| 284120 | Socoteanu, Matei | US Oncology Inc. Institutional Review Board; 10101 Woodloch Forest The Woodlands Texas 77380 United States | 18-jun-15 |
| 284122 | Fiorillo, Joseph | US Oncology Inc. Institutional Review Board; 10101 Woodloch Forest The Woodlands Texas 77380 United States | 18-jun-15 |
| 284123 | Kozloff, Mark | Ingalls Memorial Hospital IRB; 1 Ingalls Drive Ingalls Memorial Hospital Harvey Illinois 60426 United States | 22-Jul-15 |
| 284126 | Grossi, Francesco | Comitato Etico San Martino IST 2 | 11-Mar-16 |
| 284128 | Dingemans, Anne-Marie | Medical Research Ethics Committees United | 29-Oct-15 |
| 284130 | Mazieres, Julien | CPP Sud-Méditerranée 2; 270 Boulevard Sainte Marguerite Hôpital Sainte Margeurite Pavillon 9 Cedex 9 Marseille Bouches-du-Rhône 13274 France | 5-Jun-15 |
| 284672 | Braiteh, Fadi | Copernicus Group Independent Review Board; 1 Triangle Drive Suite 100 PO Box 110605 Research Triangle Park North Carolina 27709 United States | 4-Aug-15 |
| 284673 | Narang, Mohit | US Oncology Inc. Institutional Review Board; 10101 Woodloch Forest The Woodlands Texas 77380 United States | 18-jun-15 |
| 284674 | Spira, Alexander | US Oncology Inc. Institutional Review Board; 10101 Woodloch Forest The Woodlands Texas 77380 United States | 18-Jun-15 |
| 284675 | Batus, Marta | Rush University Medical Center Institutional Review Board; 1653 West Congress Parkway Chicago Illinois 60612 United States | 15-Jan-16 |
| 285270 | Tummala, Mohan | Mercy Health Springfield Communities Institutional Review Board; 1235 East Cherokee Street Springfield Missouri 65804 United States | 21-Aug-16 |
| 285271 | Jhangiani, Haresh | Copernicus Group Independent Review Board; 1 Triangle Drive Suite 100 PO Box 110605 Research Triangle Park North Carolina 27709 United States | 16-Jul-15 |
| 285273 | Menefee, Michael | Mayo Clinic Institutional Review Board; 200 First Street Southwest Rochester Minnesota 55905 United States | 10-Aug-16 |
| 285274 | Rafiyath, Shamudheen | Salus IRB; 2111 West Braker Lane Suite 400 Austin Texas 78758 United States | 11-Dec-15 |
| 286362 | Hsieh, Ruey-Kuen | Mackay Memorial Hospital Institutional Review Board | 17-Aug-15 |

| Site # | Investigator | IRB/EC Name and Address | Approval Date |
|---|---|---|---|
| 288218 | Shamrai, Volodymyr | Commission on Ethics Questions of Vinnytsya Regional Clinical Oncology Dispensary | 22-Jul-15 |
| 288407 | Orellana Ulunque, Eric | Comite Etico Cientifico Clinica Santa Maria | 19-Oct-15 |
| 288408 | Cheng, Haiying | BRANY IRB; 225 Community Drive Suite 100 Great Neck New York 11021 United States | 1-Dec-15 |
| 288409 | Eisenberg, Peter | Copernicus Group Independent Review Board; 1 Triangle Drive Suite 100 PO Box 110605 Research Triangle Park North Carolina 27709 United States | 14-Aug-15 |
| 288410 | Sabbath, Kert | Yale University Human Research Protection Program; 55 College Street New Haven Connecticut 6510 United States | 2-Mar-16 |
| 288811 | Becker, Kevin | Maimonides Med Ctr Institutional Review Board; 4802 10th Avenue Brooklyn New York 11219 United States | 1-Sep-15 |
| 288812 | Kosty, Michael | Scripps Health Institutional Review Board; 11025 North Torrey Pines Road Suite 200 La Jolla California 92037 United States | 7-Mar-16 |
| 288813 | Fabregas, Jesus | Copernicus Group Independent Review Board; 1 Triangle Drive Suite 100 PO Box 110605 Research Triangle Park North Carolina 27709 United States | 17-Nov-15 |
| 288814 | Karnad, Anand | University of Texas Health Science Center San Antonio Institutional Review Board; 7703 Floyd Curl Drive Greyhound North Campus Research Administration Room 2.104 San Antonio Texas 78229 United States | 20-Oct-15 |
| 288815 | Joshi, Abhishek | Melbourne Health Human Research Ethics Committee | 18-Nov-15 |
| 291888 | Zylla, Dylan | Park Nicollet Institute Institutional Review Board; 3800 Park Nicollet Boulevard Minneapolis Minnesota 55416 United States | 18-mar-16 |
| 292837 | Chovanec, Jozef | Eticka komisia NsP Sv. Jakuba, n.o., Bardejov | 2-Jun-16 |
| 292838 | Steppert, Claus | Ethikkommission der Bayerischen Landesärztekammer | 15-Jun-16 |
| 292994 | Ogorodnikova, Nina | CEQ of Kyiv City Clinical Oncological Center | 18-Jul-16 |
| 293115 | Hsia, Te-Chun | The Institutional Review Board of China Medical University Hospital | 3-Jun-16 |
| 293119 | Penkov, Konstantin | Ethics Committee at Private Medical Institution "Evromedservis" | 27-May-16 |
| 293120 | Areses Manrique, Maria del Carmen | CEIC de la Corporacion Sanitaria del Parc Tauli | 26-Apr-16 |
| 293124 | Hackanson, Björn | Ethikkommission der Bayerischen Landesärztekammer | 15-Jun-16 |
| 293125 | Gironés, Regina | CEIC de la Corporacion Sanitaria del Parc Tauli | 26-Apr-16 |
| 293130 | Gautschi, Oliver | Kantonale Ethikkommission Bern (KEK) | 2-Aug-16 |
| 293132 | Lammers, Ernst | Medical Research Ethics Committees United | 13-Jun-16 |
| 293294 | Le Moulec, Sylvestre | CPP Sud-Méditerranée 2; 270 Boulevard Sainte Marguerite Hôpital Sainte Margeurite Pavillon 9 Cedex 9 Marseille Bouches-du-Rhône 13274 France | 13-May-16 |
| 293295 | Barlo, Nicole | Medical Research Ethics Committees United | 13-Jun-16 |
| 293296 | Keizer - van 't Westeinde, Susan | Medical Research Ethics Committees United | 13-Jun-16 |
| 293301 | van Haarst, Jan Maarten | Medical Research Ethics Committees United | 13-Jun-16 |
| 293338 | Pazzola, Antonio | Comitato di Bioetica dell'AUSL 1 di Sassari | 20-Jun-16 |
| 293747 | Niederman, Thomas | Copernicus Group Independent Review Board; 1 Triangle Drive Suite 100 PO Box 110605 Research Triangle Park North Carolina 27709 United States | 3-May-16 |
| 293748 | Raez, Luis | Western Institutional Review Board; 1019 39th Avenue Southeast Suite 120 Puyallup Washington 98374 United States | 30-Sep-16 |
| 293749 | Porubska, Miriam | Eticka komisia Onkologicky ustav sv. Alzbety | 2-Jun-16 |
| 293750 | Vasiliev, Aleksandr | Ethics Committee at Railway Clinical Hospital JSC RZhD | 27-May-16 |
| 293751 | Koleva, Marchela | Ethics Committee for Multi-Centre Trials; 5 Sveta Nedelya Square Sofia Sofia-Grad 1000 Bulgaria | 29-Aug-2016 |

| Site # | Investigator | IRB/EC Name and Address | Approval Date |
|---|---|---|---|
| 293752 | van Lindert, Anne | Medical Research Ethics Committees United | 13-Jun-16 |
| 293753 | Mansour, Khaled | Medical Research Ethics Committees United | 13-Jun-16 |
| 294212 | Rosales, Joseph | Western Institutional Review Board; 1019 39th Avenue Southeast Suite 120 Puyallup Washington 98374 United States | 20-Jul-16 |
| 294298 | Spadafora, Silvana | Sault Area Hospital Research Ethics Board; 750 Great Northern Road Sault Ste. Marie Ontario P6B 0A8 Canada | 23-Jun-16 |
| 294828 | Naidu, Sashi | Copernicus Group Independent Review Board; 1 Triangle Drive Suite 100 PO Box 110605 Research Triangle Park North Carolina 27709 United States | 8-Jul-16 |
| 294829 | Lee, Arielle | Copernicus Group Independent Review Board; 1 Triangle Drive Suite 100 PO Box 110605 Research Triangle Park North Carolina 27709 United States | 20-Jun-16 |
| 294830 | Matthews-Smith, Velmalia | Copernicus Group Independent Review Board; 1 Triangle Drive Suite 100 PO Box 110605 Research Triangle Park North Carolina 27709 United States | 1-Jul-16 |
| 294833 | Saturnino Duarte de Brito, Ulisses | Comissão de Ética para a Investigação Clínica - CEIC | 27-Sep-16 |
| 295101 | Keogh, George | Copernicus Group Independent Review Board; 1 Triangle Drive Suite 100 PO Box 110605 Research Triangle Park North Carolina 27709 United States | 6-Sep-16 |
| 285227 | Kishi, Kazuma | Toranomon Hospital and Toranomon Hospital Kajigaya IRB, 2-2-2 Toranomon, Minato-ku, 105-8470, Tokyo, JAPAN | 14-Jul-2015 |
| 285229 | Otani, Sakiko | Kitasato University Sagamihara IRB, 1-15-1 Kitasato, Minami-ku, Sagamihara-shi, 252-0375, Kanagawa, JAPAN | 15-Jul-2015 |
| 285230 | Tanaka, Hiroshi | Niigata Cancer Center Hospital IRB, 2-15-3 Kawagishi-cho, Chuo-ku, Niigata-shi, 951-8566, Niigata, JAPAN | 13-Jul-2015 |
| 285231 | Kim, Young Hak | Kyoto University Hospital IRB, 54 Kawahara-cho Shogoin Sakyo-ku, Kyoto-shi, 606-8507, Kyoto, JAPAN | 15-Jul-2015 |
| 285232 | Kawaguchi, Tomoya | Osaka City University Hospital IRB, 1-5-7,Asahimachi, Abeno-ku, Osaka-shi, 545-8586, Osaka, JAPAN | 22-Jul-2015 |
| 285233 | Yokota, Soichiro | National Hosptial Organization Toneyama National Hospital IRB, 5-1-1 Toneyama, Toyonaka-shi, 560-8552, Osaka, JAPAN | 31-Jul-2015 |
| 285234 | Yamamoto, Nobuyuki | Wakayama Medical University IRB, 811-1 Kimiidera, Wakayama-shi, 641-8509, Wakayama, JAPAN | 21-Jul-2015 |
| 285235 | Azuma, Koichi | Kurume University IRB, 67 Asahimachi, Kurume-shi, Fukuoka, 830-0011, JAPAN | 21-Jul-2015 |
| 285236 | Seto, Takashi | National Hospital Organization Kyushu Cancer Center; IRB, 3-1-1 Notame, Minami-ku, Fukuoka-shi, 811-1395, Fukuoka, JAPAN | 1-Jul-2015 |
| 285300 | Ikeda, Satoshi | Kanagawa Cardiovascular and Respiratory Center IRB, 6-16-1 Tomiokahigashi, Kanazawa-ku, Yokohama-shi, 236-0051, Kanagawa, JAPAN | 14-Jul-2015 |
| 285301 | Ichiki, Masao | National Hospital Organization Kyushu Medical Center IRB, 1-8-1 Jigyohama, Chuo-Ku, Fukuoka-shi, 810-8563, Fukuoka, JAPAN | 22-Jul-2015 |
| 285302 | Nogami, Naoyuki | National Hospital Organization Shikoku Cancer Center IRB, 160 Minamiumemotomachi-Kou, Matsuyama-shi, 791-0280, Ehime, JAPAN | 23-Jul-2015 |
| 286439 | Fukuhara, Tatsuro | Miyagi Cancer Center IRB, 47-1 Nodayama, Medeshima-Shiote, Natori-shi, 981-1293, Miyagi, JAPAN | 15-Sep-2015 |
| 286754 | Yokoyama, Takuma | Kyorin University Hospital IRB, 6-20-2 Shinkawa, Mitaka-shi, 181-8611, Tokyo, JAPAN | 12-Aug-2015 |
| 286755 | Takeda, Yuichiro | Center Hospital of the National Center for Global Health and Medicine IRB, 1-21-1 Toyama, Shinjuku-ku, 162-8655, Tokyo, Japan | 16-Jul-2015 |
