## Supplementary material for "Allelic Variation in *HLA-DRB1* is Associated with Development of Anti-Drug Antibodies in Cancer Patients Treated with Atezolizumab that are Neutralizing *in Vitro*": Ethics Committee approvals: IMvigor211 EC IRB.pdf

| Site # | Investigator | EC/IRB Name and Address | IRB/EC Approval |
|---|---|---|---|
| 271273 | Bahl, Amit | NRES Committee London - West London and GTAC, The Old Chapel, Royal Standard Place, Nottingham, NG1 6FS, UNITED KINGDOM | 03-Jun-2015 |
| 271291 | Lee, Jae Lyun | Asan Medical Center Ethics Committee, 88, Olympic-ro 43-gil, Songpa-gu, 05505, Seoul, KOREA, REPUBLIC OF | 10-Nov-2014 |
| 271292 | Park, Se Hoon | Samsung Medical Center EC, 81, Irwon-ro, Gangnam-gu, 06351, Seoul, KOREA, REPUBLIC OF | 25-Nov-2014 |
| 271293 | Kim, Miso | Seoul National University Hospital; IRB, 101, Daehak-ro, Jongno-gu, 03080, Seoul, KOREA, REPUBLIC OF | 12-Nov-2014 |
| 271314 | Mazhar, Danish | NRES Committee London - West London and GTAC, The Old Chapel, Royal Standard Place, Nottingham, NG1 6FS, UNITED KINGDOM | 30-Apr-2015 |
| 271332 | Weickhardt, Andrew | Austin Health HREC, Research Ethics Unit, 145 Studley Road, 3084, Heidelberg, Victoria, AUSTRALIA | 18-Dec-2014 |
| 271334 | Goh, Jeffrey | Austin Health; Austin Health Human Research Ethics Committee, 145 Studley Rd, Heidelberg, 3084, Melbourne, Victoria, AUSTRALIA | 18-Dec-2014 |
| 271335 | Tan, Hsiang | Austin Health; Austin Health Human Research Ethics Committee, 145 Studley Rd, Heidelberg, 3084, Melbourne, Victoria, AUSTRALIA | 18-Dec-2014 |
| 271336 | Pook, David | Austin Health HREC, Research Ethics Unit, 145 Studley Road, 3084, Heidelberg, Victoria, AUSTRALIA | 18-Dec-2014 |
| 271349 | Syndikus, Isabel | NRES Committee London - West London and GTAC, The Old Chapel, Royal Standard Place, Nottingham, NG1 6FS, UNITED KINGDOM | 17-Mar-2015 |
| 271350 | Powles, Thomas | NRES Committee London - West London and GTAC, The Old Chapel, Royal Standard Place, Nottingham, NG1 6FS, UNITED KINGDOM | 19-Feb-2015 |
| 271456 | Crabb, Simon | NRES Committee London - West London and GTAC, The Old Chapel, Royal Standard Place, Nottingham, NG1 6FS, UNITED KINGDOM | 30-Apr-2015 |
| 271851 | Sacco, Cosimo | COMITATO ETICO UNICO REGIONALE VIA POZZUOLO 330, UDINE | 27-May-2015 |
| 271852 | Necchi, Andrea | COMITATO ETICO DELLA FONDAZIONE IRCCS "ISTITUTO NAZIONALE DEI TUMORI"- VIA G. VENEZIAN 1, MILANO | 16-Dec-2014 |
| 271853 | CARTENI, GIACOMO | COMITATO ETICO CARDARELLI-SANTOBONO VIA A. CARDARELLI, 9 NAPOLI | 28-May-2015 |
| 271854 | AGLIETTA, MASSIMO | COMITATO ETICO IRCCS DI CANDIOLO STRADA PROVINCIALE 142 CANDIOLO (TO) | 30-Apr-2015 |
| 271855 | Sabbatini, Roberto | Comitato Etico Provinciale Modena | 03-Feb-2015 |
| 271856 | De Giorgi, Ugo | Comitato Etico Di Area Vasta Romagna E Irst, Via Piero Maroncelli 40, 47014, Meldola, Emilia-Romagna, ITALY | 19-Feb-2015 |
| 271860 | STERNBERG, CORA N. | COMITATO ETICO LAZIO 1 CIRCONVALLAZIONE GIANICOLENSE 87 - ROMA | 01-Apr-2015 |
| 271861 | BRACARDA, SERGIO | COMITATO ETICO REGIONE TOSCANA - AREA VASTA SUD EST VIALE BRACCI 16 - SIENA | 16-Mar-2015 |
| 271862 | Milesi, Laura | COMITATO ETICO DELLA PROVINCIA DI BERGAMO | 26-Mar-2015 |
| 271979 | Huddart, Robert | NRES Committee London - West London and GTAC, The Old Chapel, Royal Standard Place, Nottingham, NG1 6FS, UNITED KINGDOM | 25-Mar-2015 |
| 272041 | CATHOMAS, RICHARD | Kantonale Ethikkommission Zürich (KEK) | 31-Mar-2015 |
| 272043 | Beyer, Joerg | Kantonale Ethikkommission Zürich (KEK) | 31-Mar-2015 |
| 272180 | Erman, Mustafa | Ege Universitesi Klinik Arastirmalar Etik Kurulu | 10-Apr-2015 |
| 272181 | OZGUROGLU, MUSTAFA | Ege Universitesi Klinik Arastirmalar Etik Kurulu | 10-Apr-2015 |
| 272183 | EVRENSEL, TURKKAN | Ege Universitesi Klinik Arastirmalar Etik Kurulu | 30-Apr-2015 |
| 272185 | Karabulut, Bulent | Ege Universitesi Klinik Arastirmalar Etik Kurulu | 30-Apr-2015 |
| 272228 | Birtle, Alison | NRES Committee London - West London and GTAC, The Old Chapel, Royal Standard Place, Nottingham, NG1 6FS, UNITED KINGDOM | 01-May-2015 |
| 272263 | McMenemin, Rhona | NRES Committee London - West London and GTAC, The Old Chapel, Royal Standard Place, Nottingham, NG1 6FS, UNITED KINGDOM | 19-Jun-2015 |
| 272264 | James, Nicholas | NRES Committee London - West London and GTAC, The Old Chapel, Royal Standard Place, Nottingham, NG1 6FS, UNITED KINGDOM | 03-Aug-2015 |

| | | | |
|---|---|---|---|
| 272627 | DITTRICH, CHRISTIAN | Ethikkommission der Stadt Wien gemäß KAG, AMG und MPG | 17-Apr-2015 |
| 272629 | Castellano, Daniel | Hospital Ramon y Cajal ;Comité Etico de Investigación Clínica | 11-Feb-2015 |
| 272630 | Arranz Arija, Jose Angel | Hospital Ramon y Cajal ;Comité Etico de Investigación Clínica | 11-Feb-2015 |
| 272631 | Perez Gracia, Jose Luis | Hospital Ramon y Cajal ;Comité Etico de Investigación Clínica | 11-Feb-2015 |
| 272632 | Gajate Borau, Pablo | Hospital Ramon y Cajal ;Comité Etico de Investigación Clínica Ctra. Colmenar Viejo, km 9,1 28034 MADRID | 11-Feb-2015 |
| 272636 | Morales Barrera, Rafael | Hospital Ramon y Cajal ;Comité Etico de Investigación Clínica Ctra. Colmenar Viejo, km 9,1 28034 MADRID | 11-Feb-2015 |
| 272637 | Garcia del Muro, Xavier | UZ Gent, Commissie voor Medische Ethiek, C. Heysmanslaan 10, 9000 Gent - BELGIUM | 11-Feb-2015 |
| 272638 | Perez Valderrama, Begoña | UZ Gent, Commissie voor Medische Ethiek, C. Heysmanslaan 10, 9000 Gent - BELGIUM | 11-Feb-2015 |
| 272639 | Gil, Thierry | UZ Gent, Commissie voor Medische Ethiek, C. Heysmanslaan 10, 9000 Gent - BELGIUM | 13-Apr-2015 |
| 272640 | Schrijvers, Dirk | UZ Gent, Commissie voor Medische Ethiek, C. Heysmanslaan 10, 9000 Gent - BELGIUM | 13-Apr-2015 |
| 272641 | Dumez, Herlinde | UZ Gent, Commissie voor Medische Ethiek, C. Heysmanslaan 10, 9000 Gent - BELGIUM | 13-Apr-2015 |
| 272642 | Debruyne, Philip | UZ Gent, Commissie voor Medische Ethiek, C. Heysmanslaan 10, 9000 Gent - BELGIUM | 13-Apr-2015 |
| 272643 | Rottey, Sylvie | UZ Gent, Commissie voor Medische Ethiek, C. Heysmanslaan 10, 9000 Gent - BELGIUM | 13-Apr-2015 |
| 272787 | ROLLAND, FREDERIC | CPP Sud-Ouest et Outre-mer IV- ( Docteur Claire Demiot)Centre hospitalier Esquirol Cabanis Haut- 15 rue du Docteur Marcland- | 13-Mar-2015 |
| 272805 | Flechon, Aude | CPP Sud-Ouest et Outre-mer IV- ( Docteur Claire Demiot)Centre hospitalier Esquirol Cabanis Haut- 15 rue du Docteur Marcland- 87025 LIMOGES Cedex | 13-Mar-2015 |
| 272806 | Roubaud, Guilhem | CPP Sud Ouest Et Outre Mer IV | 13-Mar-2015 |
| 272807 | Houede, Nadine | CPP Sud Ouest Et Outre Mer IV, Hôpital Jean Rebeyrol, avenue du Buisson, 87042, Limoges, FRANCE | 13-Mar-2015 |
| 272808 | Huillard, Olivier | CPP Sud Ouest Et Outre Mer IV, Hôpital Jean Rebeyrol, avenue du Buisson, 87042, Limoges, FRANCE | 13-Mar-2015 |
| 272809 | BEUZEBOC, PHILIPPE | CPP Sud-Ouest et Outre-mer IV- ( Docteur Claire Demiot)Centre hospitalier Esquirol Cabanis Haut- 15 rue du Docteur Marcland- 87025 LIMOGES Cedex | 13-Mar-2015 |
| 272810 | Hilgers, Werner | CPP Sud-Ouest et Outre-mer IV- ( Docteur Claire Demiot)Centre hospitalier Esquirol Cabanis Haut- 15 rue du Docteur Marcland- 87025 LIMOGES Cedex | 13-Mar-2015 |
| 272811 | DELVA, REMY | CPP Sud-Ouest et Outre-mer IV- ( Docteur Claire Demiot)Centre hospitalier Esquirol Cabanis Haut- 15 rue du Docteur Marcland- 87025 LIMOGES Cedex | 13-Mar-2015 |
| 272812 | OUDARD, STEPHANE | CPP Sud Ouest Et Outre Mer IV, Hôpital Jean Rebeyrol, avenue du Buisson, 87042, Limoges, FRANCE | 13-Mar-2015 |
| 272813 | SPAETH, DOMINIQUE | CPP Sud-Ouest et Outre-mer IV- ( Docteur Claire Demiot)Centre hospitalier Esquirol Cabanis Haut- 15 rue du Docteur Marcland- 87025 LIMOGES Cedex | 13-Mar-2015 |
| 272815 | Genet, Dominique | CPP Sud Ouest Et Outre Mer IV, Hôpital Jean Rebeyrol, avenue du Buisson, 87042, Limoges, FRANCE | 13-Mar-2015 |
| 272817 | THEODORE, CHRISTINE | CPP Sud Ouest Et Outre Mer IV | 13-Mar-2015 |
| 272835 | ALANKO, TUOMO | Varsinais-Suomen shp Eettinen toimikunta | 18-Nov-2014 |
| 272836 | Bamias, Aristotelis | National Ethics Committee, Ministry of Health and Social Wellfare, 284, Messogion Avenue, 15562, Cholargos, GREECE | 27-Feb-2015 |
| 272837 | Mavroudis, Dimitris | National Ethics Committee, Ministry of Health and Social Wellfare, 284, Messogion Avenue, 15562, Cholargos, GREECE | 07-Sep-2015 |
| 272838 | KALOFONOS, HARALABOS | National Ethics Committee, Ministry of Health and Social Wellfare, 284, Messogion Avenue, 15562, Cholargos, GREECE | 27-Feb-2015 |
| 272862 | GRAVIS MESCAM, GWENAELLE | CPP Sud-Ouest et Outre-mer IV- ( Docteur Claire Demiot)Centre hospitalier Esquirol Cabanis Haut- 15 rue du Docteur Marcland- 87025 LIMOGES Cedex | 13-Mar-2015 |
| 272863 | JOLY LOBBEDEZ, FLORENCE | CPP Sud Ouest Et Outre Mer IV | 13-Mar-2015 |
| 272875 | Boström, Peter | Varsinais-Suomen shp Eettinen toimikunta | 18-Nov-2014 |
| 272885 | Eigl, Bernhard | UBC BCCA Research Ethics Board (BCCA REB), 902-750 West Broadway, Fairmont Medical Building, V5Z 1H8, Vancouver, British Columbia, CANADA | 06-Mar-2015 |

| | | | |
|---|---|---|---|
| 272886 | Finch, Daygen | UBC BCCA Research Ethics Board (BCCA REB), 902-750 West Broadway, Fairmont Medical Building, V5Z 1H8, Vancouver, British Columbia, CANADA | 06-Mar-2015 |
| 272887 | Winquist, Eric | Ontario Cancer Research Ethics Board, MaRS Centre, South Tower, 101 College Stree, Suite 800, M5G 1L7, Toronto, Ontario, CANADA | 06-Feb-2015 |
| 272888 | CHENG, SUSANNA | Ontario Cancer Research Ethics Board, MaRS Centre, South Tower, 101 College Stree, Suite 800, M5G 1L7, Toronto, Ontario, CANADA | 29-Jan-2015 |
| 272889 | Ong, Michael | Ontario Cancer Research Ethics Board, MaRS Centre, South Tower, 101 College Stree, Suite 800, M5G 1L7, Toronto, Ontario, CANADA | 29-Jan-2015 |
| 272890 | Ferrario, Cristiano | McGill University; McGill University; Ethics Board, 3655 Promenade Sir William Osler - 6th Floor, H3G 1Y6, Montreal, Quebec, CANADA | 22-Jan-2015 |
| 272891 | Zalewski, Pawel | Lakeridge Health Research Ethics Board, 1 HOSPITAL COURT, L1G 2B9, OSHAWA, Ontario, CANADA | 03-Nov-2014 |
| 272893 | Heng, Daniel | HREBA - Health Research Ethics Board of Alberta - Cancer Committee, c/o Alberta Innovates - Health Solutions, Suite 1500 - 10104, 103 Avenue NW, T5J 4A7, Edmonton, Alberta, CANADA | 23-Jan-2015 |
| 272948 | Ullén, Anders | Regionala Etikprövningsnämnden i Stockholm, Box 289, Karolinska Institute, Nobelsväg 12A, 171 77, Stockholm, SWEDEN | 25-Feb-2015 |
| 272949 | Öfverholm, Elisabeth | Regionala Etikprövningsnämnden i Stockholm, Box 289, Karolinska Institute, Nobelsväg 12A, 171 77, Stockholm, SWEDEN | 25-Feb-2015 |
| 272950 | Thellenberg, Camilla | Regionala Etikprövningsnämnden i Stockholm, Box 289, Karolinska Institute, Nobelsväg 12A, 171 77, Stockholm, SWEDEN | 25-Feb-2015 |
| 272973 | WHEATLEY, DUNCAN | NRES Committee London - West London and GTAC, The Old Chapel, Royal Standard Place, Nottingham, NG1 6FS, UNITED KINGDOM | 23-Feb-2015 |
| 272983 | Vergidis, Joanna | UBC BCCA Research Ethics Board (BCCA REB), 902-750 West Broadway, Fairmont Medical Building, V5Z 1H8, Vancouver, British Columbia, CANADA | 06-Mar-2015 |
| 273022 | Puente Vazquez, Javier | Hospital Ramon y Cajal ;Comité Etico de Investigación Clínica Ctra. Colmenar Viejo, km 9,1 28034 MADRID | 11-Feb-2015 |
| 273023 | Mellado Gonzalez, Begoña | Hospital Ramon y Cajal ;Comité Etico de Investigación Clínica Ctra. Colmenar Viejo, km 9,1 28034 MADRID | 11-Feb-2015 |
| 273024 | Gallardo Diaz, Enrique | Hospital Ramon y Cajal ;Comité Etico de Investigación Clínica Ctra. Colmenar Viejo, km 9,1 28034 MADRID | 11-Feb-2015 |
| 273025 | Maroto Rey, Pablo | Hospital Ramon y Cajal ;Comité Etico de Investigación Clínica Ctra. Colmenar Viejo, km 9,1 28034 MADRID | 11-Feb-2015 |
| 273026 | MENDEZ, Mª JOSE | Hospital Ramon y Cajal ;Comité Etico de Investigación Clínica Ctra. Colmenar Viejo, km 9,1 28034 MADRID | 11-Feb-2015 |
| 273027 | Lainez Milagro, Nuria | CEIC de Navarra; Departamento de Salud. Pabellón de Docencia. | 11-Feb-2015 |
| 273028 | Gonzalez Del Alba Baamonde, Aranzazu | Hospital Ramon y Cajal ;Comité Etico de Investigación Clínica Ctra. Colmenar Viejo, km 9,1 28034 MADRID | 11-Feb-2015 |
| 273030 | Garcia Gonzalez, Jorge | Hospital Ramon y Cajal ;Comité Etico de Investigación Clínica Ctra. Colmenar Viejo, km 9,1 28034 MADRID | 11-Feb-2015 |
| 273064 | PASSALACQUA, RODOLFO | COMITATO ETICO AREA CREMONA MANTOVA E LODI, Viale Concordia 1, Servizio di Farmacia, 26100, Cremona, Lombardia, ITALY | 30-Mar-2015 |
| 273065 | Di Costanzo, Francesco | COMITATO ETICO REGIONE TOSCANA - AREA VASTA CENTRO LARGO BRAMBILLA, 3 FIRENZE | 23-Mar-2015 |
| 273066 | Morelli, Franco | SEZ DEL CE IRCCS IST TUMORI G PAOLO II BA C/O FONDAZIONE CASA SOLLIEVO DELLA SOFFERENZA SG ROTONDO - VIALE CAPPUCCINI - SAN GIOVANNI ROTONDO (FG) | 30-Mar-2015 |
| 273090 | PROTHEROE, ANDREW | NRES Committee London - West London and GTAC, The Old Chapel, Royal Standard Place, Nottingham, NG1 6FS, UNITED KINGDOM | 11-Mar-2015 |
| 273093 | Jenkins, Peter | NRES Committee London - West London and GTAC, The Old Chapel, Royal Standard Place, Nottingham, NG1 6FS, UNITED KINGDOM | 07-Apr-2015 |
| 273095 | Russell, Kent | Ontario Cancer Research Ethics Board, MaRS Centre, South Tower, 101 College Stree, Suite 800, M5G 1L7, Toronto, Ontario, CANADA | 11-Feb-2015 |
| 273237 | Tsai, Yu-Chieh | Research Ethics Committee, Nat. Taiwan Univ. Hosp., No.1, Changde Street, Zhongzheng District, 100, TAIPEI, TAIWAN | 07-Jan-2015 |
| 273238 | Chang, Yen-Hwa | TVGH Institutional Review Board, No.201, Shih-Pai Road, Sec.2, 112, Taipei, TAIWAN | 12-Feb-2015 |
| 273249 | Sundar, Santhanam | NRES Committee London - West London and GTAC, The Old Chapel, Royal Standard Place, Nottingham, NG1 6FS, UNITED KINGDOM | 16-Feb-2015 |

| | | | |
|---|---|---|---|
| 273254 | Beeker, Aart | MEC-U \| Medical Research Ethics Committees United | 15-May-2015 |
| 273256 | van der Heijden, Michiel | MEC-U \| Medical Research Ethics Committees United | 13-May-2015 |
| 273257 | Los, Maartje | MEC-U \| Medical Research Ethics Committees United | 20-Apr-2015 |
| 273258 | Coenen, J.L.L.M. | MEC-U \| Medical Research Ethics Committees United | 01-May-2015 |
| 273299 | Alyasova, Anna | EC of FSBI Privolzhsky Federal Medical Research Centre, 18-1 Verhnevolzhskaya embankment, 603155, Nizhny Novgorod, RUSSIAN FEDERATION | 05-Jun-2015 |
| 273368 | MAURICIO, JOAQUINA | CEIC - Comissão de Ética para Investigação Clínica, CEIC - Comissão de Ética para Investigação Clínica, Av. do Brasil, 53 - Pav 17-A ,Parque da Saúde de Lisboa, 1749-004, Lisboa, PORTUGAL | 27-Feb-2015 |
| 273369 | Lopes, Fábio | CEIC - Comissão de Ética para Investigação Clínica | 20-Apr-2015 |
| 273370 | Fernandes, Isabel | CEIC - Comissão de Ética para Investigação Clínica, CEIC - Comissão de Ética para Investigação Clínica, Av. do Brasil, 53 - Pav 17-A ,Parque da Saúde de Lisboa, 1749-004, Lisboa, PORTUGAL | 15-Apr-2015 |
| 273384 | Pfister, Christian | CPP Sud Ouest Et Outre Mer IV, Hôpital Jean Rebeyrol, avenue du Buisson, 87042, Limoges, FRANCE | 13-Mar-2015 |
| 273386 | Chevreau, Christine | CPP Sud-Ouest et Outre-mer IV- ( Docteur Claire Demiot)Centre hospitalier Esquirol Cabanis Haut- 15 rue du Docteur Marcland-87025 LIMOGES Cedex | 13-Mar-2015 |
| 273388 | TOURNIGAND, CHRISTOPHE | CPP Sud Ouest Et Outre Mer IV, Hôpital Jean Rebeyrol, avenue du Buisson, 87042, Limoges, FRANCE | 13-Mar-2015 |
| 273389 | CULINE, STEPHANE | CPP Sud Ouest Et Outre Mer IV, Hôpital Jean Rebeyrol, avenue du Buisson, 87042, Limoges, FRANCE | 13-Mar-2015 |
| 273466 | TOMCZAK, PIOTR | Niezalezna Komisja Bioetyczna ds Badan Naukowych, Debinki 7, budynek nr 1, III pietro, 80-211, Gdansk, POLAND | 19-Mar-2015 |
| 273467 | Senkus-Konefka, Elzbieta | Niezalezna Komisja Bioetyczna ds Badan Naukowych, Debinki 7, budynek nr 1, III pietro, 80-211, Gdansk, POLAND | 08-Jan-2015 |
| 273471 | Cernea, Dana Michaela | Comisia Nationala de Bioetica a Medicamentului si a Dispozitivelor Medicale- Sos , Sos. Stefan cel Mare nr. 19-21, Pavilion K, sector 2, 020125, Bucuresti, ROMANIA | 17-Jun-2015 |
| 273473 | Stanculeanu, Dana Lucia | Comisia Nationala de Bioetica a Medicamentului si a Dispozitivelor Medicale- Sos , Sos. Stefan cel Mare nr. 19-21, Pavilion K, sector 2, 020125, Bucuresti, ROMANIA | 12-Mar-2015 |
| 273475 | Volovat, Constantin | Comisia Nationala de Bioetica a Medicamentului si a Dispozitivelor Medicale- Sos , Sos. Stefan cel Mare nr. 19-21, Pavilion K, sector 2, 020125, Bucuresti, ROMANIA | 17-Jun-2015 |
| 273476 | Oprean, Cristina Marinela | Comisia Nationala de Bioetica a Medicamentului si a Dispozitivelor Medicale- Sos , Sos. Stefan cel Mare nr. 19-21, Pavilion K, sector 2, 020125, Bucuresti, ROMANIA | 12-Mar-2015 |
| 273478 | Wang, Shian-Shiang | The IRB, Taichung Veterans General Hospital, No. 1650, Taiwan Boulevard, Sect. 4, 407, Taichung, TAIWAN | 13-Feb-2015 |
| 273488 | Varlamov, Sergey | E.C. of Altai Oncological Center, Nikitina street, 77, 656049, Barnaul, RUSSIAN FEDERATION | 25-May-2015 |
| 273490 | Geczi, Lajos | Medical Research Council, Ethics Committee for Clinical Pharmacology, Arany J. u. 6-8., 1051, Budapest, HUNGARY | 17-Mar-2015 |
| 273493 | Csoszi, Tibor | Medical Research Council, Ethics Committee for Clinical Pharmacology, Arany J. u. 6-8., 1051, Budapest, HUNGARY | 17-Mar-2015 |
| 273500 | MELICHAR, BOHUSLAV | Eticka Komise Fakultni Nemocnice Olomouc, I.P. PAVLOVA 6, 775 20, OLOMOUC, CZECH REPUBLIC | 19-Jan-2015 |
| 273501 | Wiechno, Pawel | Niezalezna Komisja Bioetyczna ds Badan Naukowych, Debinki 7, budynek nr 1, III pietro, 80-211, Gdansk, POLAND | 08-Jan-2015 |
| 273505 | Lakomy, Radek | Eticka komise Masarykova onkologickeho ustavu, Zluty Kopec 7, 656 53, Brno, CZECH REPUBLIC | 19-Jan-2015 |
| 273512 | PETRUZELKA, LUBOS | Eticka Komise Vseobecne fakultni nemocnice, Na Bojisti 1, 128 08, Praha 2, CZECH REPUBLIC | 22-Jan-2015 |
| 273517 | Šeruga, Boštjan | Komisija Republike Slovenije za Medicinsko Etiko, Ministrstvo za zdravje, Stefanova 5, 1000, Ljubljana, SLOVENIA | 24-Nov-2014 |
| 273523 | Nyirady, Peter | Medical Research Council, Ethics Committee for Clinical Pharmacology, Arany J. u. 6-8., 1051, Budapest, HUNGARY | 17-Mar-2015 |
| 273548 | Yeh, Su-Peng | IRB, ChinaMedicalUniversityHospital, No. 2 Yuh Der Road, 404, Taichung, TAIWAN | 12-Dec-2014 |

| | | | |
|---|---|---|---|
| 273561 | Retz, Margitta | Ethikkommission Technische Universität München, Fakultät für Medizin | 23-Mar-2015 |
| 273564 | Strauß, Arne | Ethikkommission Technische Universität München, Fakultät für Medizin Ismaninger Str. 22 81675 München | 23-Mar-2015 |
| 273565 | Wirth, Manfred | Ethikkommission Technische Universität München, Fakultät für Medizin Ismaninger Str. 22 81675 München | 23-Mar-2015 |
| 273567 | Grüllich, Carsten | Ethikkommission Technische Universität München, Fakultät für Medizin Ismaninger Str. 22 81675 München | 23-Mar-2015 |
| 273568 | Miller, Kurt | Ethikkommission Technische Universität München, Fakultät für Medizin Ismaninger Str. 22 81675 München | 23-Mar-2015 |
| 273570 | Meidenbauer, Norbert | Ethikkommission Technische Universität München, Fakultät für Medizin Ismaninger Str. 22 81675 München | 23-Mar-2015 |
| 273574 | Tartas, Sophie | CPP Sud-Ouest et Outre-mer IV- ( Docteur Claire Demiot)Centre hospitalier Esquirol Cabanis Haut- 15 rue du Docteur Marcland- 87025 LIMOGES Cedex | 13-Mar-2015 |
| 273575 | RAVAUD, ALAIN | CPP Sud-Ouest et Outre-mer IV- ( Docteur Claire Demiot)Centre hospitalier Esquirol Cabanis Haut- 15 rue du Docteur Marcland- 87025 LIMOGES Cedex | 13-Mar-2015 |
| 273579 | MOLINAS, NIL | Ege Universitesi Klinik Arastirmalar Etik Kurulu | 10-Apr-2015 |
| 273580 | Cicin, Irfan | Ege Universitesi Klinik Arastirmalar Etik Kurulu | 10-Apr-2015 |
| 273637 | Schostak, Martin | Ethikkommission Technische Universität München, Fakultät für Medizin Ismaninger Str. 22 81675 München | 23-Mar-2015 |
| 273639 | Perst, Volker | Ethikkommission Technische Universität München, Fakultät für Medizin Ismaninger Str. 22 81675 München | 23-Mar-2015 |
| 273651 | Turk, Hacı Mehmet | Ege Universitesi Klinik Arastirmalar Etik Kurulu | 30-Apr-2015 |
| 273716 | Pappot, Helle | De Videnskabsetiske Komitéer for Region Hovedstaden, Regionsgården, Kongens Vænge 2, 3400 Hillerød; Denmark | 09-Feb-2015 |
| 273717 | Sengeløv, Lisa | De Videnskabsetiske Komitéer for Region Hovedstaden, Regionsgården, Kongens Vænge 2, 3400 Hillerød, Denmark | 09-Feb-2015 |
| 273744 | Køstner, Anne Helene | REK Sør-Øst, P.b. 1130 Blindern, 0318 Oslo, NORWAY | 10-Mar-2015 |
| 273845 | Kukielka-Budny, Bozena | Niezalezna Komisja Bioetyczna ds Badan Naukowych, Debinki 7, budynek nr 1, III pietro, 80-211, Gdansk, POLAND | 17-Mar-2015 |
| 274011 | Popovic, Lazar | Ethics Committee Institute for Oncology of Vojvodina, Put doktora Goldmana 4, 21204, Sremska Kamenica, SERBIA | 29-Dec-2014 |
| 274072 | AZRIA, DAVID | CPP Sud Ouest Et Outre Mer IV, Hôpital Jean Rebeyrol, avenue du Buisson, 87042, Limoges, FRANCE | 13-Mar-2015 |
| 274112 | Babovic, Nada | Ethics Committee of Institute of Oncol.& Radiology, PASTEROVA 14, 11000, BELGRADE, SERBIA | 21-Oct-2014 |
| 274134 | WOJTUKIEWICZ, MAREK | Niezalezna Komisja Bioetyczna ds Badan Naukowych, Debinki 7, budynek nr 1, III pietro, 80-211, Gdansk, POLAND | 08-Jan-2015 |
| 274191 | Mach, Nicolas | Commission centrale d'éthique, Hôpital Universitaire Genève | 02-Jun-2015 |
| 274192 | Schardt, Julian | Kantonale Ethikkommission Bern KEK | 31-Mar-2015 |
| 274193 | Omlin, Aurelius | Ethikkommission Ostschweiz (EKOS) | 31-Mar-2015 |
| 274265 | Thiery Vuillemin, Antoine | CPP Sud Ouest Et Outre Mer IV, Hôpital Jean Rebeyrol, avenue du Buisson, 87042, Limoges, FRANCE | 13-Mar-2015 |
| 274334 | Zdrojowy, Romuald | Niezalezna Komisja Bioetyczna ds Badan Naukowych, Debinki 7, budynek nr 1, III pietro, 80-211, Gdansk, POLAND | 21-May-2015 |
| 274498 | Harrison, Michael | Duke University Health System; Institutional Review Board for Clinical Investigations | 12-Jan-2015 |
| 274501 | Carthon, Bradley | WESTERN INTERNATIONAL REVIEW BOARD | 03-Dec-2014 |
| 274538 | Rathmell, Kimryn | Vanderbilt University Institutional Review Board | 26-Jan-2015 |
| 274599 | Dyar, Stephen | Western Institutional Review Board | 23-Nov-2014 |
| 274872 | Loriot, Yohann | CPP Sud-Ouest et Outre-mer IV- ( Docteur Claire Demiot)Centre hospitalier Esquirol Cabanis Haut- 15 rue du Docteur Marcland- 87025 LIMOGES Cedex | 13-Mar-2015 |
| 274880 | Ralph, Christy | NRES Committee London - West London and GTAC, The Old Chapel, Royal Standard Place, Nottingham, NG1 6FS, UNITED KINGDOM | 02-Apr-2015 |
| 275121 | Schenker, Michael | Comisia Nationala de Bioetica a Medicamentului si a Dispozitivelor Medicale- Sos , Sos. Stefan cel Mare nr. 19-21, Pavilion K, sector 2, 020125, Bucuresti, ROMANIA | 12-Mar-2015 |
| 275129 | Herzal, Amalia Alina | Comisia Nationala de Bioetica a Medicamentului si a Dispozitivelor Medicale- Sos , Sos. Stefan cel Mare nr. 19-21, Pavilion K, sector 2, 020125, Bucuresti, ROMANIA | 12-Mar-2015 |
| 275924 | von Amsberg, Gunhild | Ethikkommission Technische Universität München, Fakultät für Medizin Ismaninger Str. 22 81675 München | 23-Mar-2015 |

| | | | |
|---|---|---|---|
| 275925 | Niegisch, Günter | Ethikkommission Technische Universität München, Fakultät für Medizin Ismaninger Str. 22 81675 München | 23-Mar-2015 |
| 275933 | Barthelemy, Philippe | CPP Sud Ouest Et Outre Mer IV, Hôpital Jean Rebeyrol, avenue du Buisson, 87042, Limoges, FRANCE | 13-Mar-2015 |
| 276295 | Bedke, Jens | Ethikkommission Technische Universität München, Fakultät für Medizin<br>Ismaninger Str. 22<br>81675 München | 23-Mar-2015 |
| 276365 | Vogelzang, Nicholas | Lehigh Valley Health Network IRB | 20-Dec-2014 |
| 276366 | Philips, George | MedStar Health Research Institute-Georgetown Univ. Oncology IRB | 16-Dec-2014 |
| 276382 | Schultze-Seemann, Wolfgang | Ethikkommission Technische Universität München, Fakultät für Medizin<br>Ismaninger Str. 22<br>81675 München | 23-Mar-2015 |
| 276619 | Obara, Wataru | Iwate Medical University Institutional Review Board, 19-1, Uchimaru, Morioka-shi, 020-8505, Iwate, JAPAN | 19-Dec-2014 |
| 276620 | Osawa, Takahiro | Hokkaido University Hospital Institutional Review Board, Kita14-jo,Nishi5-chome,Kita-ku,Sapporo, 060-8648, Hokkaido, JAPAN | 16-Dec-2014 |
| 276621 | Nishimura, Kazuo | Institutional Review Board of Osaka International Cancer Institute, 3-3 Nakamichi 1-Chome, Higashinari-ku, 537-8511, Osaka, JAPAN | 18-Dec-2014 |
| 276622 | Hashine, Katsuyoshi | SHIKOKU CANER CENTER INSTITUTIONAL REVIEW BOARD, 160 Minamiumemoto-Machi-Kou; Matsuyama-Shi, 791-0280, Ehime, JAPAN | 21-Jan-2015 |
| 276623 | Kawai, Koji | University of Tsukuba Hospital Institutional Review Board, 2-1-1 Amakubo, Tsukuba-shi, 305-8576, Ibaraki, JAPAN | 25-Dec-2014 |
| 276624 | Suzuki, Kazuhiro | Gunma University Hospital Institutional Review Board, 3-39-15 Showa-machi,Maebashi-shi, 371-8511, Gunma, JAPAN | 24-Dec-2014 |
| 276625 | Yonese, Junji | The Cancer Institute Hospital of JFCR Institutional Review Board, 3-8-31 Ariake Koto-Ku, 135-8550, Tokyo, JAPAN | 07-Jan-2015 |
| 276627 | Uemura, Motohide | Osaka University Hospital Institutional Review Board, 2-15, Yamadaoka, Suita-shi, 565-0871, Osaka, JAPAN | 16-Dec-2014 |
| 276628 | Nakaigawa, Noboru | Yokohama City University Hospital Institutional Review Board, 3-9 Fukuura,Kanazawa-ku,Yokohama-shi, 236-0004, Kanagawa, JAPAN | 03-Feb-2015 |
| 276629 | Yoshimura, Kazuhiro | Kindai University Hospital Institutional Review Board, 377-2 Ohnohigashi, Osaka-Sayama-shi, 589-8511, Osaka, JAPAN | 20-Jan-2015 |
| 276630 | Fukasawa, Satoshi | Chiba Cancer Center Institutional Review Board, 666-2 Nitona-cho Chuo-ku, Chiba-shi, 260-8717, Chiba, JAPAN | 19-Dec-2014 |
| 276706 | Tandstad, Torgrim | REK Sør-Øst, P.b. 1130 Blindern, 0318 Oslo, NORWAY | 10-Mar-2015 |
| 276733 | Takano, Toshimi | Toranomon Hospital and Toranomon Hospital Kajigaya Institutional Review Board, 2-2-2 Toranomon, Minato-ku, 105-8470, Tokyo, JAPAN | 19-Feb-2015 |
| 276735 | Kanayama, Hiro-omi | Tokushima University Hospital Institutional Review Board | 24-Dec-2014 |
| 276736 | Yamashita, Ryo | Shizuoka Cancer Center Ethical Review Board for Business Clinical Studies, 1007 Shimonagakubo Nagaizumi-cho, Suntou-gun, 411-8777, Shizuoka, JAPAN | 26-Jan-2015 |
| 276755 | Yatsuda, Junji | The Institutional Review Board of Kumamoto University Hospital, 1-1-1 Honjo, Chuo-ku, Kumamoto-shi, 860-8556, Kumamoto, JAPAN | 27-Jan-2015 |
| 276756 | Fujimoto, Hiroyuki | National Cancer Center Institutional Review Board, 5-1-1 Tsukiji Chuo-Ku, 104-0045, Tokyo, JAPAN | 21-Jan-2015 |
| 276965 | Saito, Toshihiro | Niigata Cancer Center Hospital Institutional Review Board, 2-15-3 Kawagishi-cho, Chuo-ku, Niigata-shi, 951-8566, Niigata, JAPAN | 05-Mar-2015 |
| 276966 | Sassa, Naoto | Nagoya university Hospital IRB, 65 tsurumai-cho, showa-ku, nagoya-shi, 466-8560, Aichi, JAPAN | 02-Mar-2015 |
| 277103 | Faust, Guy | NRES Committee London - West London and GTAC, The Old Chapel, Royal Standard Place, Nottingham, NG1 6FS, UNITED KINGDOM | 04-Mar-2015 |
| 277110 | Worlding, Jane | NRES Committee London - West London and GTAC, The Old Chapel, Royal Standard Place, Nottingham, NG1 6FS, UNITED KINGDOM | 06-May-2015 |
| 277111 | Srinivasan, Rajaguru | NRES Committee London - West London and GTAC, The Old Chapel, Royal Standard Place, Nottingham, NG1 6FS, UNITED KINGDOM | 08-Jun-2015 |
| 277286 | Kondo, Yukihiro | Nippon Medical School Hospital Institutional Review Board, 1-1-5 Sendagi, Bunkyo-ku, 113-8603, Tokyo, JAPAN | 23-Jan-2015 |
| 277287 | Matsubara, Nobuaki | NATIONAL CANCER CENTER EAST INST. REVIEW BOARD, 6-5-1 KASHIWANOHA, KASHIWA, 277-8577, CHIBA, JAPAN | 15-Apr-2015 |
| 277375 | Koie, Takuya | Institutional Review Board of Hirosaki University School of Medicine and Hospital, 53 Honcho, Hirosaki-shi, 036-8563, Aomori, JAPAN | 30-Jan-2015 |

| | | | |
|---|---|---|---|
| 278381 | Yamaguchi, Akito | Harasanshin Hospital Institutional Review Board, 1-8 Taihaku-machi, Hakata-ku, Fukuoka-shi, 812-0033, Fukuoka, JAPAN | 22-Jan-2015 |
| 281144 | Borchiellini, Delphine | CPP Sud Ouest Et Outre Mer IV | 13-Mar-2015 |
| 281551 | Ritter, Manuel | Ethikkommission Technische Universität München, Fakultät für Medizin<br>Ismaninger Str. 22<br>81675 München | 22-May-2015 |
| 283590 | Shkodenko, Oxana | EC of State Institution of Healthcare Stavropol regional clinical oncology dispensary, 182a, Octyabrskaya Str., 355047, Stavropol, RUSSIAN FEDERATION | 10-Jun-2015 |
| 284362 | Chirivella Gonzalez, Isabel | CEIC Hospital Clinico Universitario de Valencia, Avda. Menendez Pelayo 4, accesorio, 46010, Valencia, VALENCIA, SPAIN | 20-Jul-2015 |
| 284362 | Chirivella Gonzalez, Isabel | Comite Ético de Investigación Clínica Agencia de Ensayos-HOSPITAL RAMÓN Y CAJAL, CTRA. DE COLMENAR VIEJO, KM 9,1, 28034, MADRID, MADRID, SPAIN | 20-Jul-2015 |
| 284363 | Caballero Diaz, Cristina | CEIC Hospital General Universitario de Valencia, Avda. Tres Cruces s/n, 46014, Valencia, VALENCIA, SPAIN | 20-Jul-2015 |
| 284363 | Caballero Diaz, Cristina | Comite Ético de Investigación Clínica Agencia de Ensayos-HOSPITAL RAMÓN Y CAJAL, CTRA. DE COLMENAR VIEJO, KM 9,1, 28034, MADRID, MADRID, SPAIN | 20-Jul-2015 |
| 284772 | Montesa Pino, Alvaro Gaizka | CEIC Hospital Clinico Universitario Virgen de la Victoria, Campus de Teatinos s/n, Garantia de Calidad, 29010, Malaga, MALAGA, SPAIN | 20-Jul-2015 |
| 284772 | Montesa Pino, Alvaro Gaizka | Comite Ético de Investigación Clínica Agencia de Ensayos-HOSPITAL RAMÓN Y CAJAL, CTRA. DE COLMENAR VIEJO, KM 9,1, 28034, MADRID, MADRID, SPAIN | 20-Jul-2015 |
| 284802 | Borrega, Pablo | Hospital Ramon y Cajal ;Comité Etico de Investigación Clínica | 20-Jul-2015 |
| 285284 | Aarts, Maureen | MEC-U \| Medical Research Ethics Committees United | 14-Jul-2015 |
| 288516 | Bolenz, Christian | Ethikkommission Technische Universität München, Fakultät für Medizin Ismaninger Str. 22 81675 München | 06-Nov-2015 |
